# Intravenous iron therapy with ferric carboxymaltose results in a rapid and sustained rise in myocardial iron content through a non-canonical pathway: a translational study

**DOI:** 10.1101/2024.01.18.24301496

**Authors:** M Vera-Aviles, S Kabir, A Shah, P Polzella, Y Lim, P Buckley, C Ball, D Swinkels, H Matlung, C Blans, P Holdship, J Nugent, E Andreson, M Desborough, S Piechnik, V Ferreira, S Lakhal-Littleton

## Abstract

**Background and Aims:** Intravenous iron therapies contain iron-carbohydrate complexes, designed to ensure iron becomes bioavailable via the intermediary of spleen and liver reticuloendothelial macrophages. How other tissues obtain and handle this iron remains unknown. This study addresses this question in the context of the heart.

**Methods:** A prospective observational study was conducted in 12 patients receiving ferric carboxymaltose (FCM) for iron deficiency. Myocardial, spleen and liver magnetic resonance relaxation times, and plasma iron markers were collected longitudinally. To examine the handling of iron taken up by the myocardium, intracellular labile iron pool (LIP) was imaged in FCM-treated mice and cells.

**Results:** In patients, myocardial relaxation time T1 dropped maximally 3hrs post FCM, remaining low 42 days later, while splenic T1 dropped maximally at 14 days, recovering by 42 days. In plasma, non-transferrin bound iron (NTBI) peaked at 3hrs, while ferritin peaked at 14 days. Changes in liver T1 diverged amongst patients. In mice, myocardial LIP rose 1h and remained elevated 42 days after FCM. In cardiomyocytes, FCM exposure raised LIP rapidly. This was prevented by inhibitors of NTBI transporters T-type and L-Type calcium channels and divalent metal transporter 1.

**Conclusions:** Intravenous iron therapy with FCM delivers iron to the myocardium rapidly through NTBI transporters, independently of reticuloendothelial macrophages. This iron remains labile for weeks, reflecting the myocardium’s limited iron storage capacity. These findings challenge current notions of how the heart obtains iron from these therapies and highlight the potential for long-term dosing to cause cumulative iron build-up in the heart.

**TRANSLATIONAL PERSPECTIVE:**

- Many patients now receive long-term IV iron therapy. The finding that a single standard dose of IV iron causes a sustained rise in myocardial iron underscores the risk for cumulative build-up to occur with multiple doses. Magnetic resonance monitoring of myocardial iron may be required to safeguard against progression towards pathological myocardial iron overload in these patients.
- Plasma ferritin levels reflect the iron content of reticuloendothelial macrophages. The finding that myocardial iron elevation following IV iron therapy is independent from reticuloendothelial macrophages highlights the limitations of using plasma ferritin cut-offs to safeguard against the risk of tissue iron overload.

**GRAPHICAL ABSTRACT:** 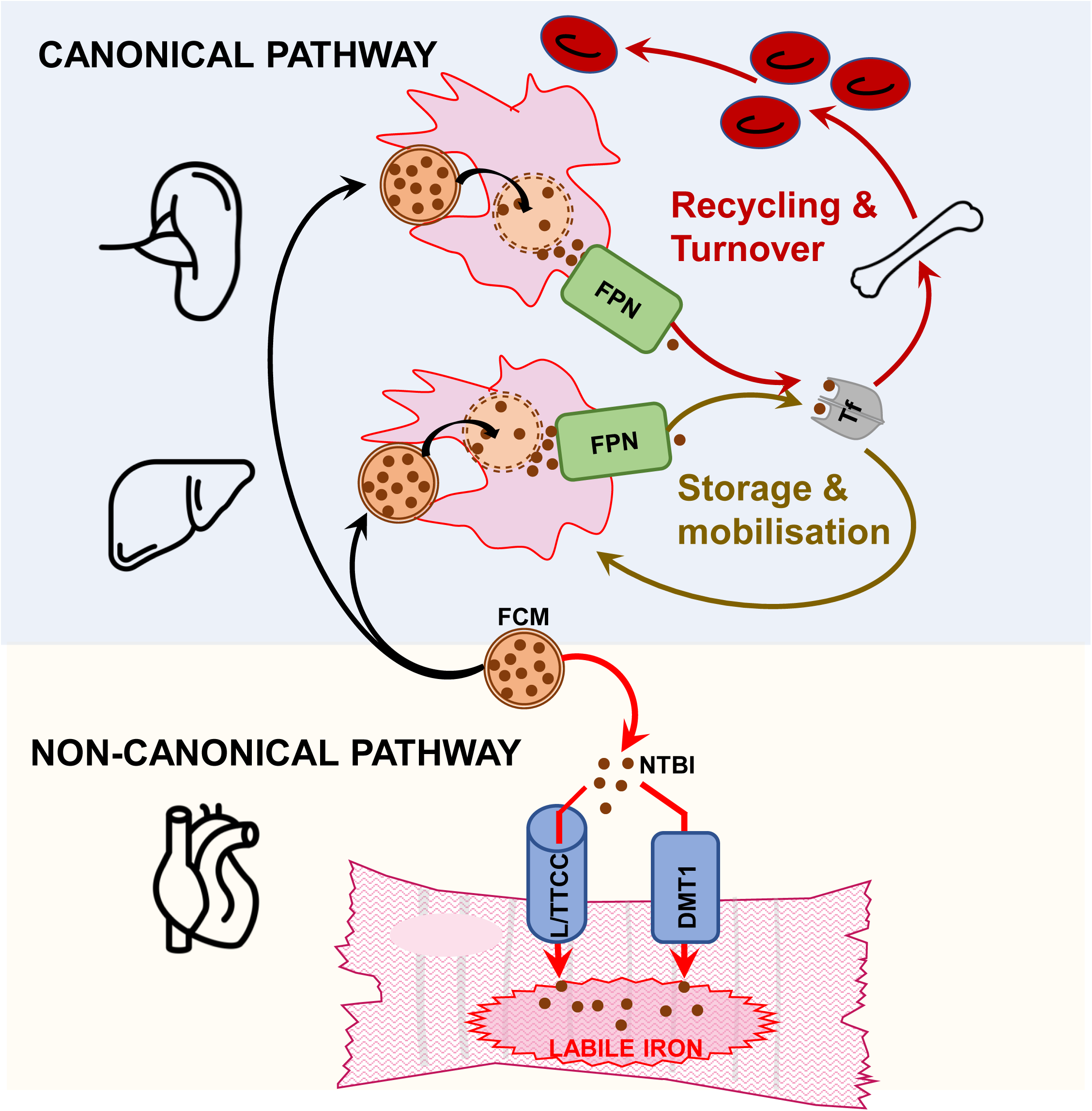

## INTRODUCTION

Iron deficiency ID is the commonest nutritional deficiency worldwide and is associated with adverse outcomes in a broad range of settings^1–12^. Thus, tackling ID efficiently and safely holds the potential to deliver cross-cutting benefits to large numbers of patients. ID has been traditionally treated with oral iron supplements, which replenish iron incrementally over weeks by delivering ∼100mg doses into the gut lumen, of which typically 10 to 22% is absorbed into the circulation ^13^. Oral iron’s gastrointestinal side effects, poor compliance and limited absorption in inflammatory settings have fuelled a gradual shift towards intravenous iron therapies^14–18^. These can deliver up to 2000mg of iron into the circulation at once. To minimise the otherwise toxic effects of free iron, the iron in IV iron therapies is contained within carbohydrate complexes^18^. These are designed for targeted uptake by macrophages of the reticuloendothelial system, in the spleen and to a lesser extent in the liver^18–20^. According to the consensus canonical pathway of IV iron metabolism, the iron-carbohydrate complex is degraded within the lysosome, iron freed, exported into the circulation and loaded onto the plasma iron chaperone transferrin ^18–20^. The iron made bioavailable through this pathway is considered safe, because the transferrin-bound iron is non-reactive and only taken up by tissues according to their needs. Indeed, surface expression of transferrin receptor (TfR) is coupled to cellular iron need, because its translation is controlled by the iron homeostatic proteins Iron Regulatory Proteins (IRPs)^21^. Inside cells, labile iron is quickly made non-reactive through storage into ferritin, the translation of which is also controlled by IRPs^21^. Because plasma ferritin derives primarily from macrophages, uptake of iron-carbohydrate complexes by reticuloendothelial macrophages is closely followed by a rise in plasma ferritin ^22–23^.

Pharmacokinetic studies have reported a rapid rise in plasma non-transferrin bound iron (NTBI) following treatment with intravenous iron, indicating that some iron is released from the carbohydrate complex directly into the circulation ^24^. What remains unknown is whether this NTBI is taken up by tissues, and the extent to which such uptake contributes to the ultimate rise in tissue iron content resulting from intravenous iron therapy. This is an important question because, unlike transferrin-bound iron, NTBI enters cells via non-canonical pathways that are not coupled to cellular iron need^25^. Because NTBI transporters are not regulated by IRPs, they continue to take up iron despite excess cellular labile iron^25^. The heart is particularly prone to NTBI uptake due to abundant expression of certain NTBI transporters including L-type and T-type calcium channels (LTCCs, TTCCs) and divalent metal transporter 1 (DMT1)^26,27^. This, together with the heart’s relatively limited capacity to store labile iron into ferritin explain why iron overload cardiomyopathy is a frequent and fatal complication in hemochromatosis and β-thalassaemia^28–30^.

Tissue iron uptake can be assessed non-invasively by magnetic resonance (MR) because the ferromagnetic properties of iron influence certain MR tissue characteristics, in particular relaxation times T1, T2, and T2*^31^. Furthermore, the handling of iron taken up into tissues can be assessed through reporter systems that rely on labile iron reactivity^32^. Using these approaches, the objective of the current study is to investigate myocardial uptake and handling of iron following intravenous iron therapy with the most widely used formulation ferric farboxymaltose (FCM, Ferinject®).

## METHODS

This report was prepared according to STROBE and ARRIVE guidelines.

### Clinical Study Design

<u>S</u>tudy of <u>T</u>issue Iron <u>U</u>ptake in Iron-<u>d</u>eficient patients following intravenous iron therap<u>y</u> (STUDY) is an investigator-initiated, prospective, observational study conducted at one UK site, sponsored by the University of Oxford. The full study protocol is available in Supplemental File 3. The trial protocol and amendments were approved by a NHS ethics committee in the UK (North West - Liverpool Central Research Ethics Committee Ref: 22/NW/0172), and the Health Research Authority. The study was registered prospectively on the ISRCTN registry (ISRCTN15770553) and clinicaltrials.gov (NCT05609318). The sample size of 12 participants was not arrived at statistically, due to lack of previous information on the magnitude of acute effects of FCM on T1/T2/T2* values. Instead, the sample size of 12 was selected based on a previous exploratory study of MRI imaging following intravenous infusion with ultrasmall superparamagnetic particles of iron oxide (USPIO), which used sample sizes of 5-12 participants per group^33^.

### Patients

Patients were recruited through the Iron Deficiency Management Service, part of the Oxford University Hospitals NHS Foundation Trust (OUHFT). Patients, aged 18 years or above, scheduled to receive intravenous iron therapy as per standard clinical care for correction of iron deficiency (ferritin less than 100mcg/L and/or transferrin saturation less than 20%) with or without anaemia (haemoglobin less than 120g/L for women and less than 130g/L for men) were invited to participate. All eligible patients identified during the screening period were contacted. Following written informed consent, patients were screened for exclusion criteria, which were any of the following: any MRI incompatible implants, pregnant or lactating participants, acute decompensated heart failure, unstable clinical status, any other medical conditions which would influence the reliability of the study results as determined by the investigators, any other contraindication to MRI. The full list of inclusion and exclusion criteria is provided within the study protocol in Supplemental File.

### Study Procedures

A flow chart outlining study procedures is shown in Appendix A of study protocol in Supplemental File.

The full CMR protocol (including the standard Siemens printout) and mapping methods are provided in supplemental File.

FCM solution (Ferinject® [FCM], Vifor Pharma, Glattbrugg, Switzerland) was given as a 20-mL intravenous infusion (equivalent to 1000 mg of iron) diluted in a sterile saline solution (0.9% wt/vol NaCl) and administered over 15 minutes. Participants were closely monitored for signs of hypersensitivity during the infusion and for at least 30 min after the treatment, as per standard clinical care.

### Study Outcomes

Primary outcome-Changes from baseline in multi-organ magnetic resonance relaxation times T1, T2 and T2* for each participant.

Secondary outcome**-** Change from baseline in plasma iron indices: iron, ferritin, transferrin saturation Tsat, non-transferrin bound iron NTBI, and serum levels of lipid peroxidation marker malondialdehyde MDA

### Mice

All animal procedures were compliant with the UK Home Office Animals (Scientific Procedures) Act 1986 (licence# P84F13B1B) and approved by the University of Oxford Medical Sciences Division Ethical Review Committee. Bioluminescence studies used B6;FVB-Ptprca Tg(CAG-luc,-GFP)L2G85Chco Thy1a/J mice (JAX strain #025854), which harbour the firefly luciferase transgene under control of CAG promoter; composed of human cytomegalovirus immediate early promoter enhancer with chicken beta-actin/rabbit beta-globin hybrid promoter. These mice have previously been characterised as having bioluminescence in the heart, spleen, muscle, pancreas, skin, thymus and bone marrow, but not in mature erythrocytes^36^. Mice were housed under standard conditions in individually-ventilated cages with enrichment at a density of 2-4 animals per cage. A priori humane endpoints were weight loss ≥15%, or signs of ill health (hunched posture and reduced activity) that did not resolve within 24 hours. Each animal represents an experimental unit. All animals were given an identifier code, to allow blinding during the conduct and analysis of experiments. Power calculations were not used due to lack of previous data on myocardial LIP.

### Manipulation of iron status in mice

Mice heterozygous for luciferase transgene were randomly assigned to a standard iron-replete diet containing 200 ppm iron (n=22), or an iron-deficient diet containing 5 ppm iron (n=25) (Teklad TD.99397), which were provided for 6 weeks.

Within each dietary group, mice were then randomly assigned to receive a 100ul injection via the tail vein of either saline (n=11 for iron-replete diet, n=12 for iron-deficient diet) or iron as ferric carboxymaltose (Ferinject® [FCM], Vifor Pharma, Glattbrugg, Switzerland) at 15mg/kg (n=11 for iron-replete group, n=13 for iron-deficient group). Mice were used for bioluminescence imaging either 1 hour or 42 days post infusion. To minimise any potential confounding effects of age and sex, all animals entered the diary protocol at 6 weeks of age, and block randomisation was used to ensure balanced numbers of males and females in each group. Randomisation into diets and treatments was carried out using the Rand() function in excel.

### Labile iron imaging in murine hearts

Bioluminescence studies used the iron-caged luciferin described previously^32^.

Mice were anaesthetised using 2% isoflurane in O_2_, and injected via the tail vein with 25nmoles of iron-caged luciferin (ICL-1) dissolved in 100ul saline. One minute later, the chest cavity was opened, and the heart immediately surgically excised, washed in ice-cold PBS to remove any blood clots, and placed on a petri dish within the IVIS Lumina system. Bioluminescence imaging was performed in an IVIS LUMINA II system, with small binning and F stop 1 settings. Bioluminescence signal was collected for at least 2 minutes, with auto settings. Luminescence data were extracted using Living Image Software4.7.3. First, a non-luminescence photograph was used to generate ROIs, by manually drawing around the outline of each heart (excluding atria). ROIs corresponding to individual hearts were automatically propagated onto the luminescence sequence, and average radiance extracted from each ROI as p/s/cm^2^/sr (photons, per second, per square cm, per steradian). The mean of the average radiance values acquired over in the first two minutes were calculated for each heart.

Following removal of the heart, blood was immediately collected from the chest cavity for Hb measurement and preparation of serum. Spleens and livers were also removed, washed in ice-cold PBS before snap-freezing for quantitation of total iron content.

No animals were lost to humane endpoints. However, some animals did not undergo myocardial LIP imaging due to failure to infuse ICL-1 intravenously prior to imaging. Their spleens and livers were still harvested for direct iron quantitation.

### Iron quantitation in murine tissues

Snap-frozen livers and spleens were crushed on liquid nitrogen, and a minimum of 5mg digested in nitric acid using a CEM microwave system. Elemental component analysis was carried out using induced couple plasma mass spectrometry ICP-MS as per previous studies ^37^. Values are normalised to tissue weight.

### Cardiac myocytes

For in-vitro bioluminescence studies, the rat cardiac myocyte cell line H9C2 (ATCC CRL1446, cardiac myoblasts from rat) was transfected with Firefly Luiferase-eGFP Lentivirus (Tetubio, product reference #14979980) and positive selection was maintained by inclusion of Geneticin in growth media. Transfected cells were maintained in complete Dulbecco′s Modified Eagle′s Medium (DMEM) growth medium, supplemented with 10% fetal bovine serum (FBS) in standard tissue culture conditions.

### Labile iron imaging in cultured cardiac myocytes

Cells were plated in complete media in opaque 96 well plates at 50000cells/well overnight. To eliminate any confounding effects of existing iron in FBS, growth media were changed to FBS-free DMEM media two hours before treatment with sterile saline or FCM, still in FBS-free media. The amount of FCM added to the FBS-free media were determined as follows; as the standard human FCM dose is 15mg iron/Kg, and the average plasma volume in humans is 60ml/Kg, the concentration of iron delivered to human plasma following a standard FCM dose is 0.25mg/mL. Thus FCM was added to the FBS-free growth media at a concentration of 0.25mg iron/mL to mimic the standard human dose.

Prior to imaging, saline and FCM-containing media were removed, cells washed with PBS, then ICL-1 added at 100uM to each well, and plates imaged immediately in an IVIS LUMINA II system, using the sequence settings described above. Luminescence data were extracted using Living Image Software, using the standard 12X8 grid ROI setting.

### Haematological and iron parameters

Peripheral venous blood from participants was collected into lithium heparin tubes. Some blood was subject to haematological analysis for a full blood count using an ABX Pentra 60 system and the remainder was used to extract plasma by spinning at 2500g for 10min at 4C. Plasma samples were immediately stored at −80C for further analysis.

Haemoglobin in mice was recorded from fresh blood using the HemoCue Hb 201+ system. To extract serum, blood was allowed to clot at room temperature for 2 hours, before spinning at 4000g for 10 min. Serum was stored at −80C for further analysis.

Iron, serum ferritin, transferrin levels were determined in human plasma and mouse serum using the ABX-Pentra C400 system (Horiba). Transferrin saturation was calculated using iron and transferrin concentrations. NTBI measurements were carried out by Sanquin Diagnostic Services (Amesterdam) as described previously^38^. It was not possible to obtain data on serum iron and NTBI from some animals due to the amount of serum collected not meeting the minimum volume requirements for the respective assays.

MDA levels were measured in plasma or serum using Lipid Peroxidation kit Abcam (ab233471) according to the manufacturer’s instructions.

### Statistical analysis

Data in figures are shown as mean ±standard error of the mean ±S.E.M. In-text values are reported as mean ±STDEV.

Normality and equal variance were tested using the Shapiro-Wilk test and Levene test respectively. Pairwise comparisons were drawn using 2-tailed, unpaired t test (for sample size n≥5), or Mann-Whitney test (for sample size<5). Multiple group comparisons were drawn using a 1-way or 2-way ANOVA followed by Dunnett’s post hoc test. Longitudinal repeated measure data in patients were analysed using mixed effects analysis to account for missing data, with Dunnett’s multiple comparison test. P value of less than 0.05 was considered statistically significant. Analysis was carried out using GraphPad PRISM.10

## RESULTS

### Ferric carboxymaltose treatment in patients results in rapid and sustained rise in myocardial iron

Between October 18^th^ 2022 and July 31^st^ 2023, 54 patients were screened for eligibility, of whom 13 were recruited, and 12 completed the study (Figure 1). Patient demographics and baseline characteristics are shown in Table 1.

**Figure 1.**
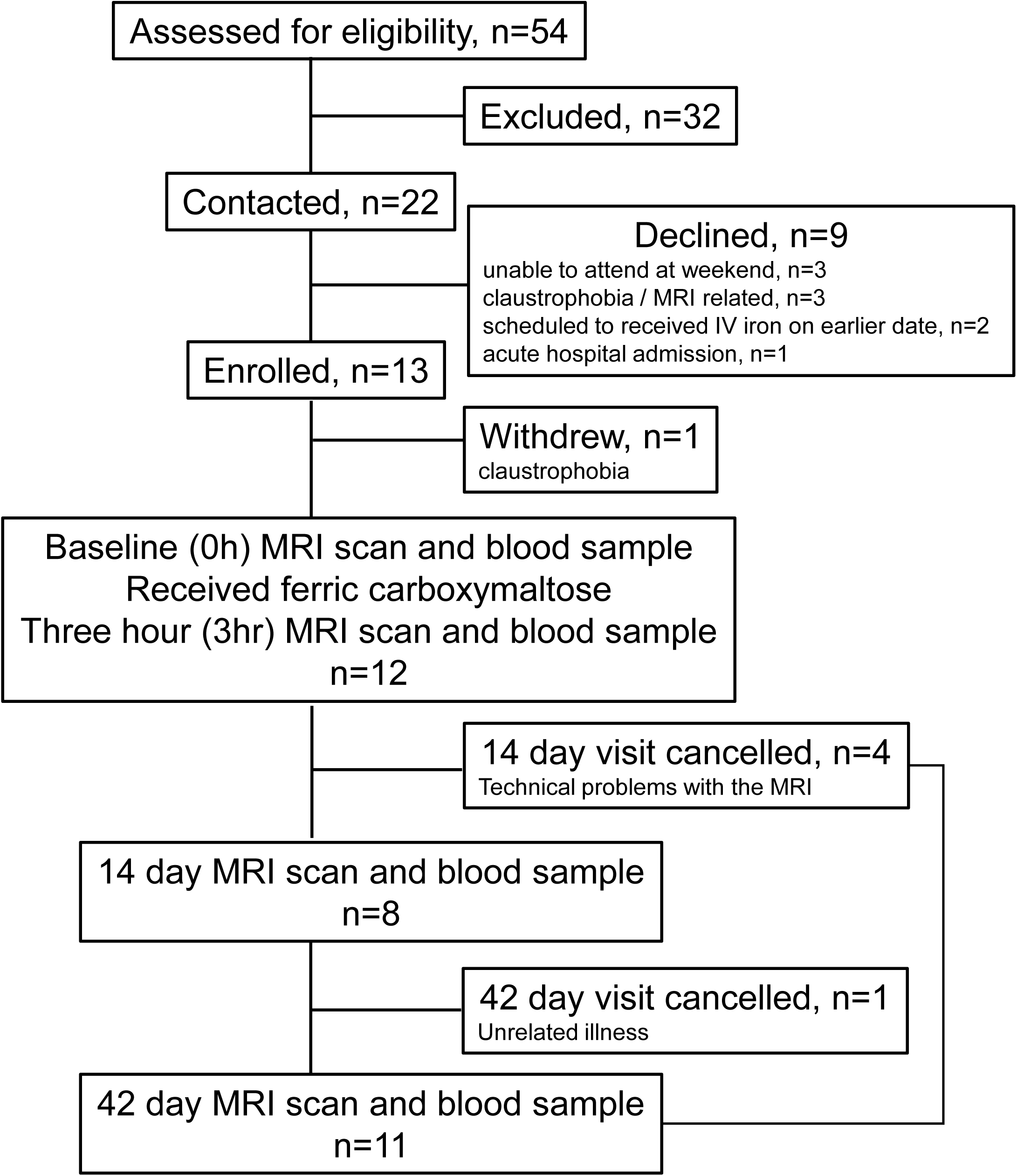
Trial profile.

**Table 1.** Patient demographics and baseline characteristics (n=12)

| Characteristic | Value |
| --- | --- |
| **Age (years), median (IQR), [Range]** | 44 (12) [27-69] |
| **Sex, n (%)** |  |
| Female / Male | 11 (91)/ 1 (9) |
| **Weight (kg), mean (SD)** | 71.2 (13.7) |
| **BMI (kg/m<sup>2</sup>), median (IQR)** | 27.2 (5.7) |
| **Ethnicity, n (%)** |  |
| White | 6 (50) |
| South Asian | 4 (33) |
| Black | 1 (17) |
| Mixed | 0 (0) |
| Other | 1 (17) |
| **Reason for referral n, (%)** |  |
| Heavy menstrual bleeding | 8 (66.8%) |
| Abnormal uterine bleeding (e.g. fibroids) | 1 (8.3%) |
| Rectal bleeding | 1 (8.3%) |
| IDA of unclear aetiology | 2 (16.6%) |
| **Previous IV iron infusions, n (%)** |  |
| None | 9 (75) |
| One prior infusion | 3 (25) |
| **Relevant comorbidities, n (%)** |  |
| Ischaemic heart disease | 0 (0) |
| Chronic respiratory disease (asthma, COPD) | 0 (0) |
| **Laboratory iron parameters at referral (Normal range), median (IQR)** |  |
| Haemoglobin (120- 170), g/l | 106 (12) |
| Ferritin (10 – 300), mcg/l | 7.4 (10.1) |
| Tsat (16 – 50), % | 7 (3) |

BMI, Body Mass Index; COPD, Chronic Obstructive Pulmonary Disease; IDA, Iron deficiency anaemia; IQR, Interquartile range; IV, intravenous; SD, Standard deviation; Tsat, Transferrin saturation

To assess the short and long-term impact of FCM on myocardial iron, participants were scanned immediately prior to infusion (0h), then three hours (3h), 14 days and 42 days post infusion (Figure 2A). At 3h post infusion, myocardial T1 dropped in 11/12 participants, with a mean change (ΔT1) of −35.35±26.88ms(p=0.0008). At 14 days and 42days post infusion, myocardial T1 was still lower than 0h values, with a change of −26.76±32.93ms (p=0.0255) and −28.45±20.33ms (p=0.008) respectively. Importantly, ΔT1 values at days 14 and 42 were not significantly different from those at 3h post infusion (p=0.709 and 0.775 respectively) (Figure 2B). Thus, a single standard dose of FCM in patients with iron deficiency raises myocardial iron rapidly and maximally within 3 hours, and this rise is sustained for at least 42 days post infusion.

**Figure 2.**
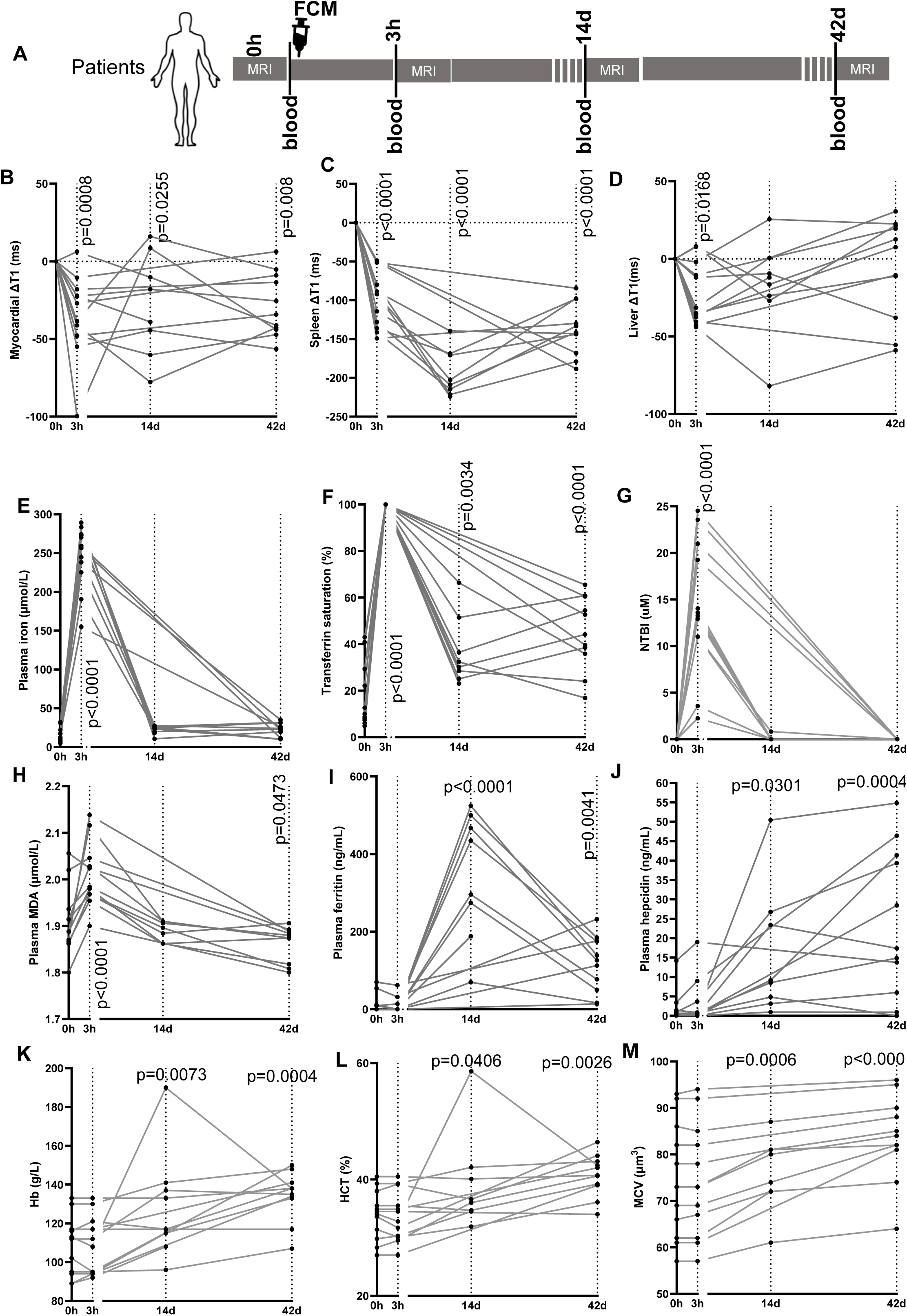
Ferric carboxymaltose treatment in patients results in rapid and sustained rise in myocardial iron. A) Schematic of study design. B) Longitudinal changes from baseline (0h) in myocardial T1 (ΔT1) for each participant. C) Longitudinal changes from baseline in splenic T1 (ΔT1) for each participant, D) Longitudinal changes from baseline in liver T1 (ΔT1) for each participant. Per-participant longitudinal assessments of plasma iron (E), transferrin saturation Tsat (F), plasma non-transferrin-bound iron NTBI (G), plasma lipid peroxidation marker MDA (H), plasma ferritin (I), plasma hepcidin (J), haemoglobin Hb (K), haematocrit HCT (L) and mean corpuscular volume MCV (M).

To understand the pathways underlying the rise in myocardial iron, we also assessed changes in spleen and liver iron, where reticuloendothelial macrophages reside. Splenic T1 dropped in all participants at 3hrs post infusion, with a mean drop (ΔT1) of −39.7±36.2ms (p<0.0001). However maximal drop of −193.9±30.59ms was only seen at 14 days (p<0.0001). By 42 days post infusion, the mean drop ΔT1 was −137±33.68ms (p<0.0001). Between days 14 and 42, there was a statistically significant recovery in ΔT1 values (p=0.0004) (Figure 2C). Thus, after a single standard dose of FCM in patients with iron deficiency, spleen iron begins to rise within 3 hours, but rises maximally at 14 days, and is declining by 42 days..

Next, we assessed T1 changes in the liver. At 3h post infusion, liver T1 dropped in 11/12 participants. The mean ΔT1 was −23±19.9ms (p=0.0168). By 14 days and 42days post infusion, liver ΔT1values had diverged markedly amongst participants, and were no longer statistically significantly different from 0h values (p=0.093 and 0.829 respectively) (Figure 2D).

In terms of circulating iron, plasma iron concentration rose sharply in all participants from a mean concentration of 14.05±2.52uM at 0h to 246.64±10.60uM at 3 hours (p<0.0001), then declined to 22.01±1.14uM at 14 days and to 22.23±2.31uM at 42 days such that plasma iron levels at these timepoints were no longer significantly higher than what they were at 0h (p=0.78 and 0.722 respectively) (Figure 2E).

Transferrin saturation (Tsat) rose sharply in all participants from 16.74±3.65% at 0h to full saturation at 3 hours (p<0.0001), then declined to 36.68±3.78% at 14 days and 44.82±4.12% at 42 days, though it remained significantly higher than it was at 0h (p=0.0034 and p<0.0001 respectively) (Figure 2F).

Plasma NTBI levels rose sharply in all participants from being below detection (assay LLOD is 0.607uM) at 0h to 14.16±1.87uM at 3 hours post FCM infusion(p<0.0001), then declined at day 14 to below the LLOD in all participants but one, such that the mean concentration at this timepoint was 0.102±0.073μM at 14 days (p=0.999 compared to 0h). At 42 days post infusion, NTBI levels were below the detection limit for all participants (>0.999 compared to 0h) (Figure 2G).

The presence of labile iron entities such as NTBI generates peroxides which in turn react with susceptible molecules, including certain lipids. Plasma malondialdehyde (MDA), a biomarker of lipid peroxidation rose in all participants from a mean of 1.908±0.0185uM at 0h to 2.007±0.0180uM at 3 hours (p<0.0001). They declined to 1.886±0.005uM at day 14 such that they were no longer different from 0h (p=0.7816), and then declined further to 1.858±0.010uM at day 42 (p=0.0473 relative to 0h) (Figure 2H)

In terms of plasma ferritin, mean levels were 12.77±6.27ug/mL at 0h and were not significantly different at 3hours post infusion (8.925±5.08ug/mL, p=0.9983). However, they rose to 344±41.59 ug/mL at 14 days (p<0.0001 relative 0h) and 65.8±18.99 ug/mL at 42 days (p=0.0041 relative to 0h) The decline in ferritin levels from day 14 to day 42 was significant (p=0.0029) (Figure 2I).

We also assessed the levels of the iron homeostatic hormone hepcidin, which were at 2.12±ng1.139ng/mL at 0h, remaining unchanged at 3h (3.16±1.55ng/ml, p=0.9923) but rising significantly to 15.9±4.27ng/mL at 14 days (p=0.0301) and further still to 23.93±5.05ng/mL at 42 days (p=0.0004) (Figure 2J).

Haematological parameters, including haemoglobin, haematocrit and mean corpuscular volume were all significantly improved at days 14 and 42 compared to 0h (Figure 2k-m).

These data demonstrate that following infusion of a standard dose of FCM in iron-deficient patients, the myocardium takes up iron quickly, within 3 hours, coinciding with maximal rises in serum iron availability, both transferrin and non-transferrin bound. The iron taken up within the first 3 hours is retained in the myocardium for at least 42 days, despite declining serum iron availability. This is in contrast to the spleen, where iron uptake is maximal at 14 days, subsequently declining by 42days,. Additionally, changes in plasma ferritin levels followed the pattern of changes in splenic iron.

### Ferric carboxymaltose treatment in mice results in rapid and sustained rise in myocardial labile iron pool (LIP)

Having observed an increase in myocardial iron following FCM infusion in patients, we sought to determine how this iron is handled in the myocardium. To that effect, we used iron-caged luciferin (ICL-1), which is only converted into the substrate of the cytoplasmic enzyme luciferase in the presence of cytoplasmic labile iron. This approach was used to quantitatively assess the impact on myocardial LIP of a single intravenous infusion of FCM (15mg/kg iron) into iron-replete or iron-deficient mice 1 hour or 42 days after infusion (Figure 3A). The iron status of mice was confirmed (supplemental Figure 2A, B).

**Figure 3.**
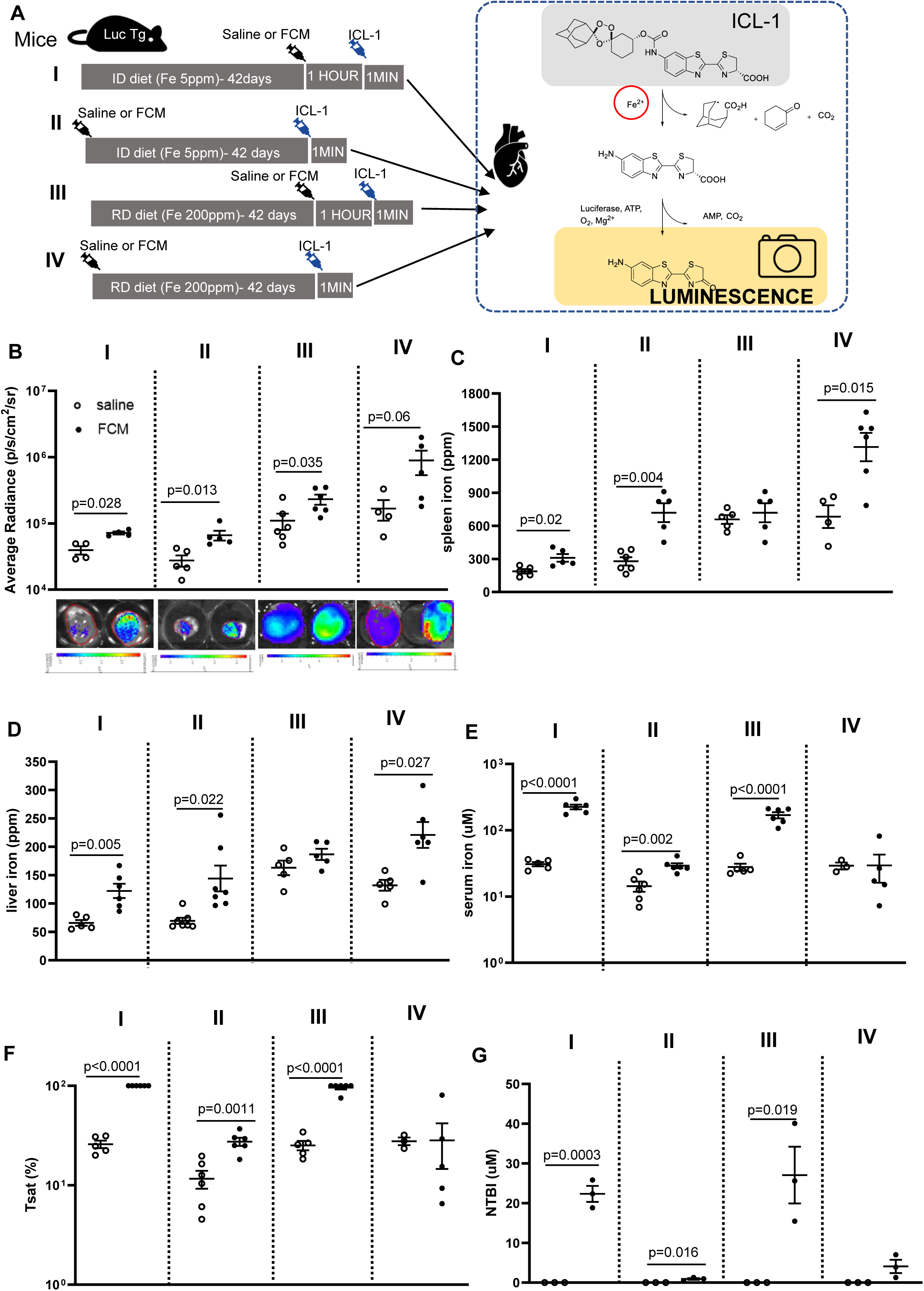
Ferric carboxymaltose treatment in mice results in rapid and sustained rise in myocardial labile iron pool LIP. A) Schematic of study design. ID=iron-deficient diet containing 5 parts per million (ppm) iron. RD=replete diet containing 200ppm iron. ICL-1=iron-caged luciferin. Luc Tg mice are transgenic for the luciferase gene. **B)** Average radiance in hearts of iron-deficient or iron replete mice 1 hour or 42 days after intravenous infusion of saline or FCM (15mg/kg iron). Representative luminescence images of hearts from each experimental group are also shown. **C**) Total elemental iron concentration in the spleen. D) Total elemental iron concentration in the liver. E) Concentrations of iron in serum, F) Transferrin saturation Tsat in serum, G) Concentrations of non-transferrin bound iron NTBI in serum.

In FCM-treated iron-deficient mice, myocardial LIP was raised both 1 hour after infusion (p=0.028 vs saline) and 42 days after infusion (p=0.013 vs saline) (Figure 3B). In FCM-treated iron-replete mice, myocardial LIP was raised 1 hour post infusion (p=0.035 vs saline) and there was also a trend for the increase to be sustained 42 days after infusion (p=0.06 saline vs FCM) (Figure 3B). Of note, myocardial LIP was lower in iron-deficient mice than in iron-replete mice (p=0.0047) (Supplemental figure 2C)

As with patients, we sought to understand the pathways underpinning the rise in myocardial iron. FCM infusion into iron-deficient mice raised spleen iron within 1 hour (p=0.02 relative to saline), through the rise was much more pronounced 42 days post infusion (p=0.004 relative to saline). In iron-replete mice, FCM infusion did not raise spleen iron after 1 hour, but did so after 42 days (p=0.015 relative to saline) (Figure 3C). Changes in liver iron followed a similar trend as changes in spleen iron (Figure 3D). In terms of circulating iron availability, serum iron levels were raised by FCM infusion in comparison to saline in iron-deficient mice 1 hour after infusion (p<0.0001 relative to saline), and were still raised 42 days post infusion (p=0.0002 relative to saline). In iron-replete mice, serum iron levels were raised 1 hour after FCM infusion (p<0.0001 relative to saline) but not significantly so 42 days after infusion (p=0.392 relative to saline) (Figure 3E). Changes in transferrin saturation (Tsat) reflected changes in serum iron levels, with full or near full saturation seen 1 hour after FCM infusion (Figure 3F). Consistent with this, NTBI was below the detection limit in saline-treated mice, but rose markedly 1 hour after FCM infusion in both iron-deficient mice (p=0.0003 relative to saline) and iron-replete mice (p=0.019 relative to saline). There was some residual NTBI 42 days post FCM infusion in iron-deficient mice (p=0.016 relative to saline) (Figure 3G).

FCM did not have an acute effect on serum ferritin in either iron-deficient of iron-replete mice, but raised serum ferritin in iron-deficient mice after 42 days (p=0.0155 relative to saline) (supplemental figure 2D).

These preclinical data confirm the previous clinical observation that FCM results in a rapid and sustained increase in myocardial iron and further demonstrate that the iron taken up by the myocardium is in labile (reactive) form, and remains so for at least 42 days post treatment.

### Exposure to ferric carboxymaltose raises cellular LIP in cardiac myocytes via NTBI transporters

The rapid rise in myocardial iron following FCM infusion indicates direct myocardial uptake of iron from the circulation through a pathway that bypasses reticuloendothelial macrophages. To ascertain direct uptake and determine its route(s), we used the rat cardiac myocyte cell line H2C9, transfected via lentivirus, to express the luciferase transgene (Supplemental Figure 3). FCM was added to the growth media at a concentration of 0.25mg iron/mL to reflect a standard patient dose. To eliminate any confounding effects of iron from the growth media additive fetal bovine serum (FBS), cardiac myocytes were switched to FBS-free growth media two hours before and throughout exposure to FCM or saline (Figure 4A).

**Figure 4.**
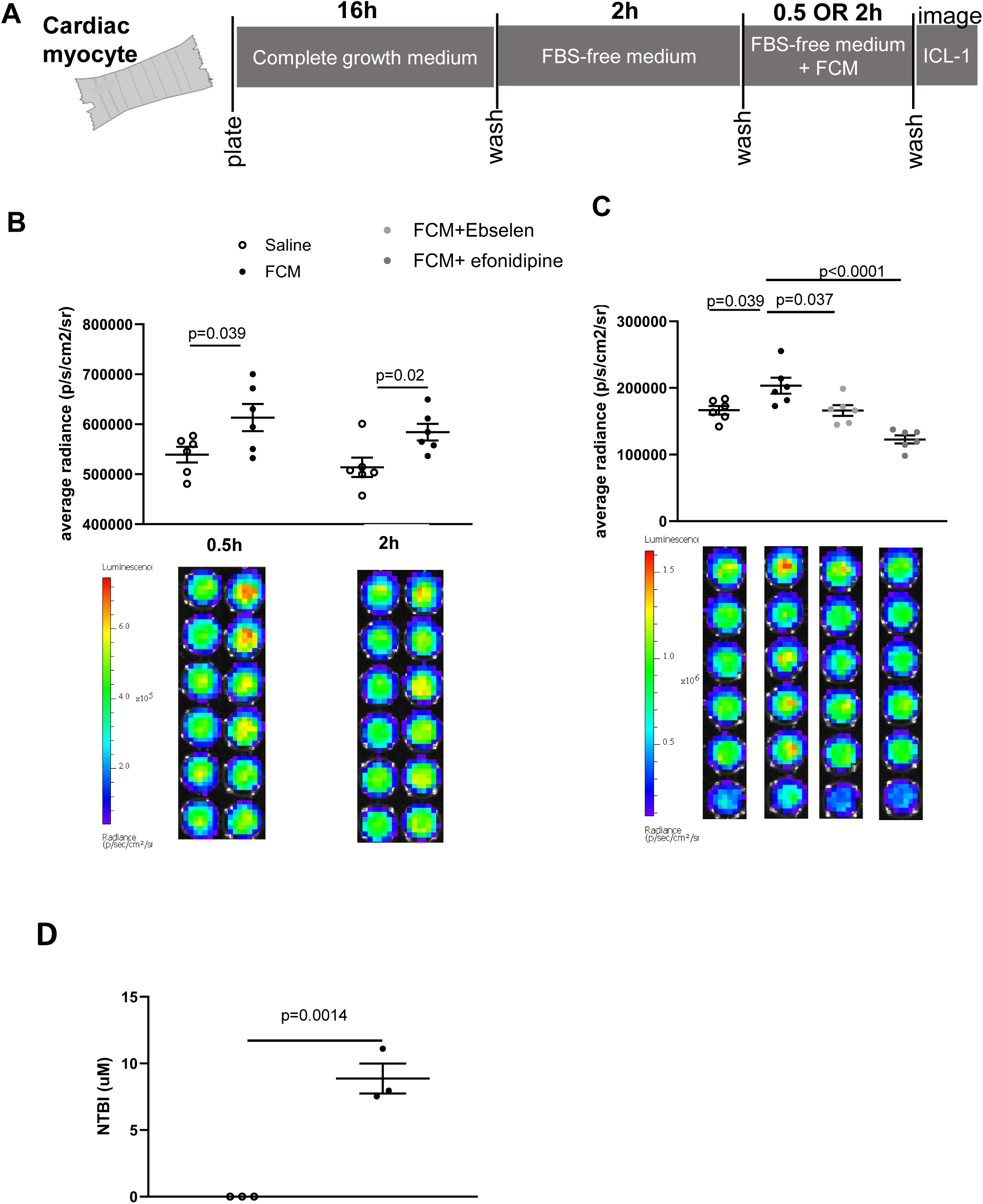
Exposure to Ferric carboxymaltose raises cellular LIP in cardiac myocytes via NTBI transporters. A) Schematic of study design using rat cardiac myocytes, expressing the luciferase transgene. B) Average radiance in cardiac myocytes after 0.5h or 2h exposure to saline or FCM (0.25mg/ml iron) in growth media. Luminescence of individual wells are shown in bottom panel. C) Average radiance in cardiac myocytes after 1h exposure to saline, FCM (0.25mg/ml iron) alone, or in combination with ebselen (DMT1 inhibitor) or efonidipine (L-type and T-type calcium channel inhibitor). Luminescence of individual wells are shown in bottom panel D) NTBI concentration in supernatants of cells exposed to saline or FCM (0.25mg/ml).

Compared to saline treatment, FCM in growth media raised cellular LIP in cardiac myocytes at 0.5h (p=0.039 relative to saline) and 2h (p=0.0204 relative to saline) (Figure 4B). Given that this rise occurred in the absence of FBS (the source of transferrin in growth media), we postulated that it was the result of NTBI uptake. To test this hypothesis, we determined the impact of efonidipine, an inhibitor of L-type and T-type calcium channels (LTCC and TTCC), and Ebselen, an inhibitor of the ferrous iron transporter divalent metal transporter DMT1. We found that Ebselen reduced (p=0.0372 compared to FCM alone) and efonidipine completely prevented (p<0.0001 compared to FCM alone) the rise in cardiac myocyte LIP (Figure 4C).

Consistent with this, NTBI was present in the supernatants of FCM-treated cells, being below the detection limit in the supernatants of saline-treated cells (p=0.0014) (Figure 4D).

These data confirm that FCM treatment raises LIP in cardiac myocytes and further demonstrate this rise is due to the uptake of extracellular NTBI derived from FCM.

## Discussion

While IV iron therapies have been suggested to raise the iron content of certain tissues, including the heart^39^, the mechanisms underlying this rise, and the ultimate fate of the iron that is taken up remained unknown. The present study addresses these unknowns in the context of the heart, through longitudinal assessment of changes in myocardial iron relative to changes in splenic, liver and circulating iron, and specific tracing, of cellular labile iron, the immediate product of iron uptake.

The key and novel finding of the present study is that the rise in myocardial iron following a standard dose of intravenous iron with FCM results from rapid and direct uptake of FCM-derived NTBI in the circulation. Indeed, in iron-deficient patients and in mice, myocardial iron content peaked within hours of FCM infusion, a timeframe that is too rapid to be attributed to the canonical pathway of FCM degradation by reticuloendothelial macrophages. Indeed, splenic iron in patients (closely followed by plasma ferritin levels), only peaked at 14 days post infusion, and began to decline by 42 days post infusion, consistent with the initial uptake of FCM and subsequent release of iron into the circulation. Acute rises in circulating iron levels, including the appearance of NTBI in patients and in mice, and the presence of NTBI in the supernatants of FCM-treated cells demonstrate that ferric carboxymaltose complex releases some of its iron directly into the circulation and in quantities that are sufficient to alter the iron content of the myocardium. A previous pharmacokinetic study of intravenous iron formulations in healthy volunteers also detected acute rises in NTBI and estimated that 10-40% of the iron contained within FCM is released rapidly and directly into the circulation as NTBI^24^. In cultured cardiac myocytes, exposure to clinically relevant dose of FCM raised intracellular iron content in the absence of any macrophages, and in a manner that was primarily dependent on LTCC and or TTCC, and to a lesser extent DMT1, recognised NTBI transporters that are abundantly expressed in the heart, and known to be responsible for iron-overload cardiomyopathy in thalassaemia and hemochromatosis ^25–27^. While DMT1 is regulated by IRPs according to cellular iron needs, LTCCs and TTCCs are not regulated in this manner, continuing to take up iron into cells despite excess intracellular iron levels.

The second key and novel finding of the present study is that the iron taken up into the myocardium following FCM treatment remains in labile form for weeks. Specialised organs of iron handling, such as the spleen and liver, are characterised by high iron storage capacity and rapid iron turnover owing to abundant levels of ferritin and ferroportin respectively,^28,29,40^. That iron taken up into the myocardium remains labile 42 days later, is consistent with the fact that the myocardium is not a specialised organ of iron handling, and has relatively limited capacity for iron storage and turnover^28,29^. Intriguingly, in the spleen and liver, other relaxation parameters T2 and T2* echoed changes in T1, dropping significantly following FCM infusion, with maximal drops at 14 days post infusion (supplemental figure 4A-D). In contrast, changes in myocardial T2 and T2* did not echo changes in myocardial T1 and diverged greatly between participants, with myocardial ΔT2 only trending towards a significant drop at 42 days post infusion (p=0.086 compared to 0h) (supplemental figure 4E,F). The lack of concordance between myocardial relaxation parameters T1, T2 and T2* is a well-recognised phenomenon ^41–43^. T1 has been shown to have higher sensitivity than T2* for detecting milder myocardial iron loading in thalassaemia patients, and may be better at resolving small increases following IV iron therapy^41,42^. Changes in relaxation parameters reflect microscopic inhomogeneity in the magnetic field, created by the distribution of iron inside cells. Unbound labile iron has a diffuse cytoplasmic distribution, in contrast to stored iron, where up to 4500 atoms of iron are clustered inside the 80 Å diameter cavity of the ferritin molecule^43^. Thus, it is likely that labile iron affects the magnetic field, and consequently relaxation parameters, differently from the stored iron. Our findings are consistent with the idea that T1 better reflects labile iron entities, while T2 and T2* better reflect stored iron entities.

The findings of the current study have important clinical implications. The lack of iron clearance from the myocardium over the period of 42 days highlights the potential for cumulative build up with repeated doses. Based on the observed mean drop in myocardial T1 of −28.45ms after a single IV iron dose, calculations estimate that 6 doses would cumulatively lower myocardial T1 to below 850ms, the cut-off shown to predict iron overload cardiomyopathy ^44^ (supplemental table 1). This raises pertinent questions about the safety of long-term IV iron therapy, increasingly being adopted in some patients, e.g. malabsorptive disorders, women with heavy uterine bleeding, chronic disorders^45,46^. Trials of long-term IV iron therapy in heart failure patients (where up to 9 doses were administered) have not reported an increased risk of adverse effects^1, 47–49^. However, myocardial iron content was not monitored in these studies, and any manifestation of cardiac iron toxicity could have been masked by the clinical signs of pre-existing heart failure. In these settings, minimum and maximum ferritin cut offs are often used to inform the need for further IV iron doses, and to safeguard against the risk of parenchymal iron overload respectively. However, our findings that myocardial iron uptake following IV iron therapy is independent of reticuloendothelial macrophages indicate that are also uncoupled from changes in plasma ferritin levels. Hence ferritin cut-offs may not be appropriate to safeguard against the risk of myocardial iron overload in patients on long term IV iron therapy, and MR based monitoring of myocardial iron should be considered instead. Additionally, in settings where myocardial iron repletion maybe desirable, the benefits sustained by patients would be independent of changes in plasma ferritin levels^1^.

### Study limitations

The clinical findings of the present study are derived from a relatively small number of participants. Most of these were women with iron deficiency due to heavy menstrual bleeding, and it remains to be determined if the study’s findings can be generalised to patient groups with different aetiologies of iron deficiency. Though not specific to this study, the use of MR relaxometry to estimate tissue iron content has a its limitations. For instance, relaxometry results depend on the choice of sequence and inferences on tissue iron content hinge on a gross simplification that the relaxivity coefficients for all/any iron species in blood and tissue are similar across all study visits^50^.

The pre-clinical findings of the present study rely on luminescence-based imaging of labile iron, with the main limitation that imaging is conducted on isolated hearts, to safeguard against interferences from surrounding tissues, including the epidermis and fur. Excision of the heart prior to LIP imaging has the potential to impact on luciferase enzyme activity and total luminescence signal, though this risk would have been present in all experimental groups.

## Acknowledgements

SL-L conceived project, secured funds, analysed data and wrote manuscript. PP and MD recruited patients. AS performed study procedures in participants. VF and SP designed MR imaging protocol and analysed MR data. MVA, KS, YL, CB, PB, PH, DS, HM performed experiments. All reviewed and commented on manuscript.

We are grateful to OCMR radiographers Dilliram Adhikari, Neil Fox, Joana Leal Pelado and Rebecca Mills for running MRI scans.

We are grateful to all the participants (and their families) who took part, without whom this study would not have been possible.

## Funding

S L-L is funded by the Medical Research Council (MR/ V009567/1/) and the British Heart Foundation Centre for Research Excellence HSR00031 and RE/18/3/34214. YL is funded by the British Heart Foundation Centre for Research RE/18/3/34214. CB is funded by the Wellcome Trust. AS is currently supported by a National Institute for Health and Care Research Academic Clinical Lectureship award.

## Disclosure of interest

S.L-L reports receipt of previous research funding from Vifor Pharma, personal honoraria on a lecture from Pharmacosmos and consultancy fees from Disc Medicine and ScholarRock. AS is an Editor of Anaesthesia.

## Data availability statement

Research data will be made available upon reasonable request.

## Notes

### Author Declarations

The trial protocol and amendments were approved by a NHS ethics committee in the UK (North West - Liverpool Central Research Ethics Committee Ref: 22/NW/0172), and the Health Research Authority. The study was registered prospectively on the ISRCTN registry (ISRCTN15770553) and clinicaltrials.gov (NCT05609318).

### Summary of Updates

Two authors added to the authorship list as they had previously been ommitted in error

## References

1. Lakhal-Littleton, S, Cleland JG. Iron deficiency and supplementation in heart failure. Nature Reviews Cardiology. In press

2. von Haehling S, Gremmler U, Krumm M, Mibach F, Schön N, Taggeselle J, et al. Prevalence and clinical impact of iron deficiency and anaemia among outpatients with chronic heart failure: The PrEP Registry. Clin Res Cardiol. 2017 Jun;106(6):436–43. doi:10.1007/s00392-016-1073-y

3. Jankowska EA, Rozentryt P, Witkowska A, Nowak J, Hartmann O, Ponikowska B, et al. Iron deficiency: an ominous sign in patients with systolic chronic heart failure. Eur Heart J. 2010 Aug;31(15):1872–80. doi:10.1093/eurheartj/ehq158

4. Ghio A, Hilborn ED. Indices of iron homeostasis correlate with airway obstruction in an NHANES III cohort. Int J Chron Obstruct Pulmon Dis. 2017 Jul 18;12:2075–84. doi:10.2147/copd.s138457

5. Silverberg DS, Mor R, Tia Weu M, Schwartz D, Schwartz IF, Chernin G. Anemia and iron deficiency in COPD patients: prevalence and the effects of correction of the anemia with erythropoiesis stimulating agents and intravenous iron. BMC Pulm Med. 2014 Feb 24;14(1). doi:10.1186/1471-2466-14-24

6. Eisenga MF, Nolte IM, van der Meer P, Bakker SJL, Gaillard CAJM. Association of different iron deficiency cutoffs with adverse outcomes in chronic kidney disease. BMC Nephrol. 2018 Sep 12;19(1). doi:10.1186/s12882-018-1021-3

7. Eisenga MF, Minović I, Berger SP, Kootstra-Ros JE, van den Berg E, Riphagen IJ, et al. Iron deficiency, anemia, and mortality in renal transplant recipients. Transpl Int. 2016 Nov;29(11):1176–83. doi:10.1111/tri.12821

8. Crawford J, Cella D, Cleeland CS, Cremieux P, Demetri GD, Sarokhan BJ, et al. Relationship between changes in hemoglobin level and quality of life during chemotherapy in anemic cancer patients receiving epoetin alfa therapy. Cancer. 2002 Aug 15;95(4):888–95. doi:10.1002/cncr.10763

9. Gasche C, Lomer MCE, Cavill I, Weiss G. Iron, anaemia, and inflammatory bowel diseases. Gut. 2004 Aug;53(8):1190–7. doi:10.1136/gut.2003.035758

10. Dignass AU, Gasche C, Bettenworth D, Birgegård G, Danese S, Gisbert JP, et al. European Consensus on the Diagnosis and Management of Iron Deficiency and Anaemia in Inflammatory Bowel Diseases. J Crohns Colitis. 2015 Mar;9(3):211–22. doi:10.1093/ecco-jcc/jju009

11. Vidal F, Gillibert A, Quillard M, Fardellone P, Vittecoq O, Lequerré T. Does iron deficiency contribute to fatigue in patients with rheumatoid arthritis without anemia? Joint Bone Spine. 2020 Jan;87(1):89. doi:10.1016/j.jbspin.2019.06.004

12. Fowler AJ, Ahmad T, Phull MK, Allard S, Gillies MA, Pearse RM. Meta-analysis of the association between preoperative anaemia and mortality after surgery. Br J Surg. 2015 Oct;102(11):1314–24. doi: 10.1002/bjs.9861.

13. Low MSY, Speedy J, Styles CE, De-Regil LM, Pasricha S-R. Daily Iron Supplementation for improving anaemia, iron status and health in menstruating women. Cochrane Database Syst Rev. 2016 Apr 18;2016(4). doi:10.1002/14651858.cd009747.pub2

14. American Regent, Inc. Venofer Prescribing Information. Shirley, New York: American Regent, Inc.; 2022. Available from: https://www.venofer.com/pdfs/venofer-prescribing-information.pdf

15. Sanofi Aventis US. Ferrlecit Prescribing Information. Silver Spring, Maryland: Food and Drug Administration; 2022. Available from: https://www.accessdata.fda.gov/drugsatfda_docs/label/2022/020955s020lbl.pdf

16. Pharmacosmos Therapeutics Inc. MONOFERRIC Prescribing Information. Silver Spring, Maryland: Food and Drug Administration; 2022. Available from: https://www.accessdata.fda.gov/drugsatfda_docs/label/2022/208171Orig1s002lbl.pdf

17. American Regent, Inc. Injectafer Prescribing Information. Silver Spring, Maryland: Food and Drug Administration; 2023. Available from: https://www.accessdata.fda.gov/drugsatfda_docs/label/2023/203565s024lbl.pdf

18. Funk F, Flühmann B, Barton AE. Criticality of Surface Characteristics of Intravenous Iron– Carbohydrate Nanoparticle Complexes: Implications for Pharmacokinetics and Pharmacodynamics. Int J Mol Sci. 2022 Feb 15;23(4):2140. doi:10.3390/ijms23042140

19. Alphandéry E. Iron oxide nanoparticles for therapeutic applications. Drug Discov Today. 2020 Jan;25(1):141–9. doi:10.1016/j.drudis.2019.09.020

20. Arami H, Khandhar A, Liggitt D, Krishnan KM. In vivo delivery, pharmacokinetics, biodistribution and toxicity of iron oxide nanoparticles. Chem Soc Rev. 2015 Dec 7;44(23):8576–607. doi:10.1039/c5cs00541h

21. Rouault T, Klausner R. Regulation of iron metabolism in eukaryotes. Current Topics in Cellular Regulation. 1997 Jan 1;35:1–19. doi:10.1016/s0070-2137(97)80001-5

22. Cohen LA, Gutierrez L, Weiss A, Leichtmann-Bardoogo Y, Zhang D, Crooks DR, et al. Serum ferritin is derived primarily from macrophages through a nonclassical secretory pathway. Blood. 2010 Sep 2;116(9):1574–84. doi:10.1182/blood-2009-11-253815

23. Ferring-Appel D, Hentze MW, Galy B. Cell-autonomous and systemic context-dependent functions of iron regulatory protein 2 in mammalian iron metabolism. Blood. 2009 Jan 15;113(3):679–87. doi:10.1182/blood-2008-05-155093

24. Garbowski MW, Bansal S, Porter JB, Mori C, Burckhardt S, Hider RC. Intravenous iron preparations transiently generate non-transferrin-bound iron from two proposed pathways. Haematologica. 2020 Nov 1;106(11):2885–96. doi:10.3324/haematol.2020.250803

25. Knutson MD. Non-transferrin-bound iron transporters. Free Radic Biol Med. 2019 Mar;133:101–11. doi:10.1016/j.freeradbiomed.2018.10.413

26. Oudit GY, Sun H, Trivieri MG, Koch SE, Dawood F, Ackerley C, et al. L-type Ca2+ channels provide a major pathway for iron entry into cardiomyocytes in iron-overload cardiomyopathy. Nat Med. 2003 Sep 1;9(9):1187–94. doi:10.1038/nm920

27. Kumfu S, Chattipakorn S, Chinda K, Fucharoen S, Chattipakorn N. T-type calcium channel blockade improves survival and cardiovascular function in thalassemic mice. Eur J Haematol. 2012 Jun;88(6):535–48. doi:10.1111/j.1600-0609.2012.01779.x

28. Powell LW, Alpert E, Isselbacher KJ, Drysdale JW. Human Isoferritins: Organ Specific Iron and Apoferritin Distribution. Br J Haematol. 1975 May;30(1):47–55. doi:10.1111/j.1365-2141.1975.tb00516.x

29. Srivastava AK, Reutovich AA, Hunter NJ, Arosio P, Bou-Abdallah F. Ferritin microheterogeneity, subunit composition, functional, and physiological implications. Sci Rep. 2023 Nov 14;13(1):19862. doi:10.1038/s41598-023-46880-9

30. Udani K, Chris-Olaiya A, Ohadugha C, Malik A, Sansbury J, Paari D. Cardiovascular manifestations in hospitalized patients with hemochromatosis in the United States. Int J Cardiol. 2021 Nov 1;342:117–24. doi:10.1016/j.ijcard.2021.07.060

31. Messroghli DR, Moon JC, Ferreira VM, Grosse-Wortmann L, He T, Kellman P, Mascherbauer J, Nezafat R, Salerno M, Schelbert EB, Taylor AJ, Thompson R, Ugander M, van Heeswijk RB, Friedrich MG. Clinical recommendations for cardiovascular magnetic resonance mapping of T1, T2, T2* and extracellular volume: A consensus statement by the Society for Cardiovascular Magnetic Resonance (SCMR) endorsed by the European Association for Cardiovascular Imaging (EACVI). J Cardiovasc Magn Reson. 2017 Oct 9;19(1):75. doi: 10.1186/s12968-017-0389-8. Erratum in: J Cardiovasc Magn Reson. 2018 Feb 7;20(1):9.

32. Aron AT, Heffern MC, Lonergan ZR, Vander Wal MN, Blank BR, Spangler B, et al. In vivo bioluminescence imaging of labile iron accumulation in a murine model of Acinetobacter baumannii infection. Proc Natl Acad Sci U S A. 2017 Nov 28;114(48):12669–74. doi:10.1073/pnas.1708747114

33. Lagan J, Naish JH, Simpson K, Zi M, Cartwright EJ, Foden P, Morris J, Clark D, Birchall L, Caldwell J, Trafford A, Fortune C, Cullen M, Chaudhuri N, Fildes J, Sarma J, Schelbert EB, Schmitt M, Piper Hanley K, Miller CA. Substrate for the Myocardial Inflammation-Heart Failure Hypothesis Identified Using Novel USPIO Methodology. JACC Cardiovasc Imaging. 2021 Feb;14(2):365–376. doi: 10.1016/j.jcmg.2020.02.001

34. Piechnik SK, Ferreira VM, Dall’Armellina E, Cochlin LE, Greiser A, Neubauer S, Robson MD. Shortened Modified Look-Locker Inversion recovery (ShMOLLI) for clinical myocardial T1-mapping at 1.5 and 3 T within a 9 heartbeat breathhold. J Cardiovasc Magn Reson. 2010 Nov 19;12(1):69. doi: 10.1186/1532-429X-12-69.

35. Zhang Q, Werys K, Popescu IA, Biasiolli L, Ntusi NAB, Desai M, Zimmerman SL, Shah DJ, Autry K, Kim B, Kim HW, Jenista ER, Huber S, White JA, McCann GP, Mohiddin SA, Boubertakh R, Chiribiri A, Newby D, Prasad S, Radjenovic A, Dawson D, Schulz-Menger J, Mahrholdt H, Carbone I, Rimoldi O, Colagrande S, Calistri L, Michels M, Hofman MBM, Anderson L, Broberg C, Andrew F, Sanz J, Bucciarelli-Ducci C, Chow K, Higgins D, Broadbent DA, Semple S, Hafyane T, Wormleighton J, Salerno M, He T, Plein S, Kwong RY, Jerosch-Herold M, Kramer CM, Neubauer S, Ferreira VM, Piechnik SK. Quality assurance of quantitative cardiac T1-mapping in multicenter clinical trials - A T1 phantom program from the hypertrophic cardiomyopathy registry (HCMR) study. Int J Cardiol. 2021 May 1;330:251–258. doi: 10.1016/j.ijcard.2021.01.026.

36. Cao Y-A, Wagers AJ, Beilhack A, Dusich J, Bachmann MH, Negrin RS, et al. Shifting foci of hematopoiesis during reconstitution from single stem cells. Proc Natl Acad Sci U S A. 2004 Jan 6;101(1):221–6. doi:10.1073/pnas.2637010100

37. Lakhal-Littleton S, Wolna M, Carr CA, Miller JJJ, Christian HC, Ball V, et al. Cardiac ferroportin regulates cellular iron homeostasis and is important for cardiac function. Proc Natl Acad Sci U S A. 2015 Mar 10;112(10):3164–9. doi:10.1073/pnas.1422373112

38. Jacobs EMG, Hendriks JCM, van Tits BLJH, Evans PJ, Breuer W, Liu DY, et al. Results of an international round robin for the quantification of serum non-transferrin-bound iron: Need for defining standardization and a clinically relevant isoform. Anal Biochem. 2005 Jun 15;341(2):241–50. doi:10.1016/j.ab.2005.03.008

39. Núñez J, Miñana G, Cardells I, Palau P, Llàcer P, Fácila L, Almenar L, López-Lereu MP, Monmeneu JV, Amiguet M, González J, Serrano A, Montagud V, López-Vilella R, Valero E, García-Blas S, Bodí V, de la Espriella-Juan R, Lupón J, Navarro J, Górriz JL, Sanchis J, Chorro FJ, Comín-Colet J, Bayés-Genís A; Myocardial-IRON Investigators* †. Noninvasive Imaging Estimation of Myocardial Iron Repletion Following Administration of Intravenous Iron: The Myocardial-IRON Trial. J Am Heart Assoc. 2020 Feb 18;9(4):e014254. doi: 10.1161/JAHA.119.014254.

40. Nemeth E, Ganz T. Hepcidin-Ferroportin Interaction Controls Systemic Iron Homeostasis. Int J Mol Sci. 2021 Jun 17;22(12):6493. doi:10.3390/ijms22126493

41. Meloni A, Martini N, Positano V, De Luca A, Pistoia L, Sbragi S, et al. Myocardial iron overload by cardiovascular magnetic resonance native segmental T1 mapping: a sensitive approach that correlates with cardiac complications. J Cardiovasc Magn Reson. 2021 Jun 14;23(1):70. doi:10.1186/s12968-021-00765-w

42. Torlasco C, Cassinerio E, Roghi A, Faini A, Capecchi M, Abdel-Gadir A, et al. Role of T1 mapping as a complementary tool to T2* for non-invasive cardiac iron overload assessment. PLoS One. 2018 Feb 21;13(2):e0192890. doi:10.1371/journal.pone.0192890

43. Theil EC. Ferritin: the protein nanocage and iron biomineral in health and in disease. Inorg Chem. 2013;52(21):12223–12233. doi:10.1021/ic400484n

44. Singh SP, Jagia P, Ojha V, Seth T, Naik N, Ganga KP, et al. Diagnostic Value of T1 Mapping in Detecting Iron Overload in Indian Patients with Thalassemia Major: A Comparison with T2* Mapping. Indian Journal of Radiology and Imaging. 2024;34(01):54–9. doi:10.1055/s-0043-1772467

45. Shand AW, Nassar N. Rapid increase in intravenous iron therapy for women of reproductive age in Australia. Med J Aust. 2021 Apr;214(6):285–285.e1. doi: 10.5694/mja2.50979.

46. Niepel D, Klag T, Malek NP, Wehkamp J. Practical guidance for the management of iron deficiency in patients with inflammatory bowel disease. Therap Adv Gastroenterol. 2018 Apr 26;11:1756284818769074. doi: 10.1177/1756284818769074.

47. Ponikowski P, van Veldhuisen DJ, Comin-Colet J, Ertl G, Komajda M, Mareev V, McDonagh T, Parkhomenko A, Tavazzi L, Levesque V, Mori C, Roubert B, Filippatos G, Ruschitzka F, Anker SD; CONFIRM-HF Investigators. Beneficial effects of long-term intravenous iron therapy with ferric carboxymaltose in patients with symptomatic heart failure and iron deficiency†. Eur Heart J. 2015 Mar 14;36(11):657–68. doi: 10.1093/eurheartj/ehu385.

48. Kalra PR, Cleland JGF, Petrie MC, Thomson EA, Kalra PA, Squire IB, Ahmed FZ, Al-Mohammad A, Cowburn PJ, Foley PWX, Graham FJ, Japp AG, Lane RE, Lang NN, Ludman AJ, Macdougall IC, Pellicori P, Ray R, Robertson M, Seed A, Ford I; IRONMAN Study Group. Intravenous ferric derisomaltose in patients with heart failure and iron deficiency in the UK (IRONMAN): an investigator-initiated, prospective, randomised, open-label, blinded-endpoint trial. Lancet. 2022 Dec 17;400(10369):2199–2209. doi: 10.1016/S0140-6736(22)02083-9.

49. Mentz RJ, Garg J, Rockhold FW, Butler J, De Pasquale CG, Ezekowitz JA, Lewis GD, O’Meara E, Ponikowski P, Troughton RW, Wong YW, She L, Harrington J, Adamczyk R, Blackman N, Hernandez AF; HEART-FID Investigators. Ferric Carboxymaltose in Heart Failure with Iron Deficiency. N Engl J Med. 2023 Sep 14;389(11):975–986. doi: 10.1056/NEJMoa2304968.

50. Moon JC, Messroghli DR, Kellman P, Piechnik SK, Robson MD, Ugander M, Gatehouse PD, Arai AE, Friedrich MG, Neubauer S, Schulz-Menger J, Schelbert EB; Society for Cardiovascular Magnetic Resonance Imaging; Cardiovascular Magnetic Resonance Working Group of the European Society of Cardiology. Myocardial T1 mapping and extracellular volume quantification: a Society for Cardiovascular Magnetic Resonance (SCMR) and CMR Working Group of the European Society of Cardiology consensus statement. J Cardiovasc Magn Reson. 2013 Oct 14;15(1):92. doi: 10.1186/1532-429X-15-92.

