## supplemental file for "Intravenous iron therapy with ferric carboxymaltose results in a rapid and sustained rise in myocardial iron content through a non-canonical pathway: a translational study"

### Supplemental figures

#### Supplemental Figure 1

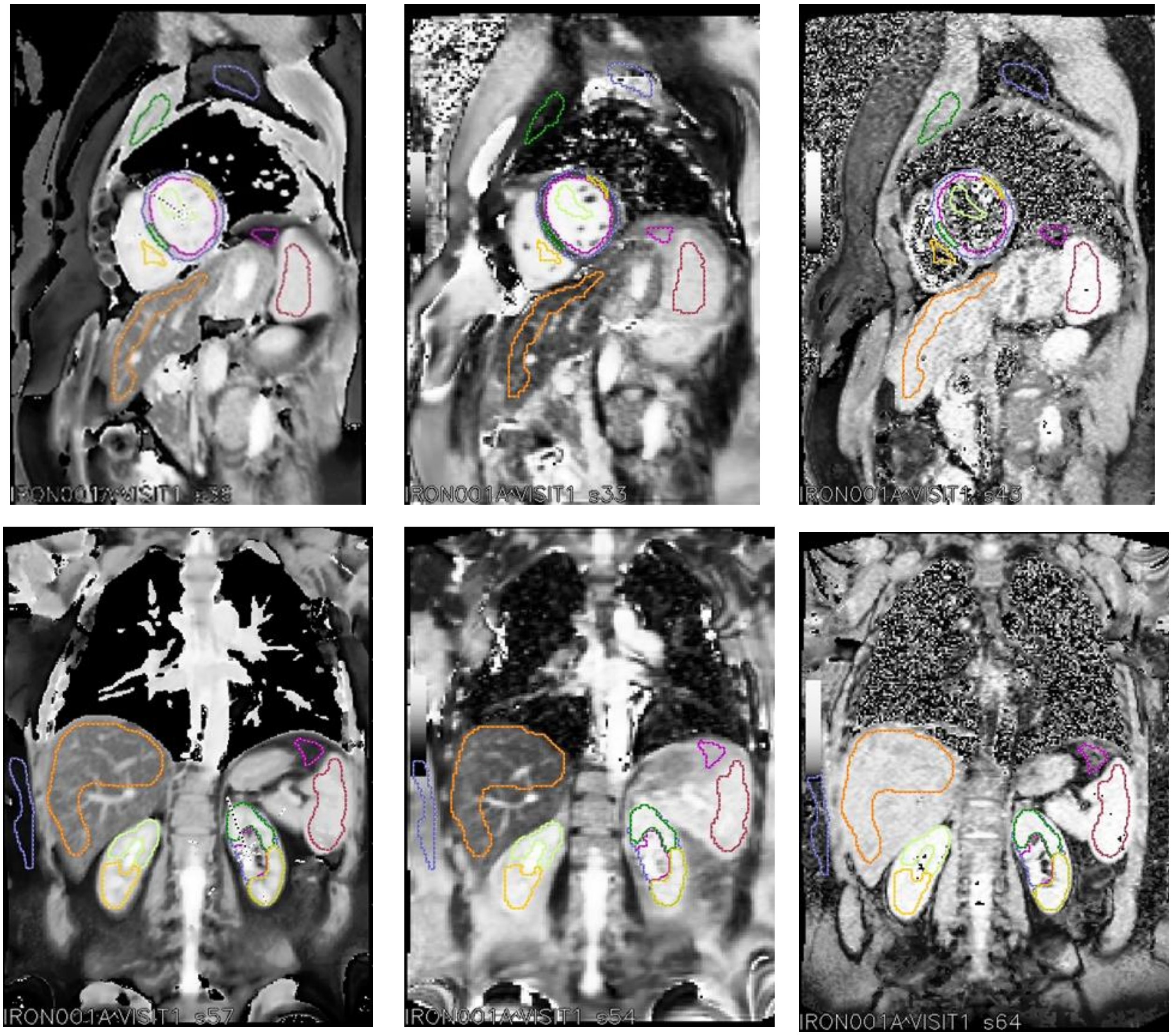

**Supplemental Figure 1**
Representative T1, T2 and T2* maps (left to right, respectively) for the cardiac and liver/spleen planes (top, bottom row, respectively). The coloured outlines indicate the edges of the regions of interest overlaid for the tissue of interest. Images have been histogram equalised to help identifying the distinctive tissue classes.

#### Supplemental Figure 2

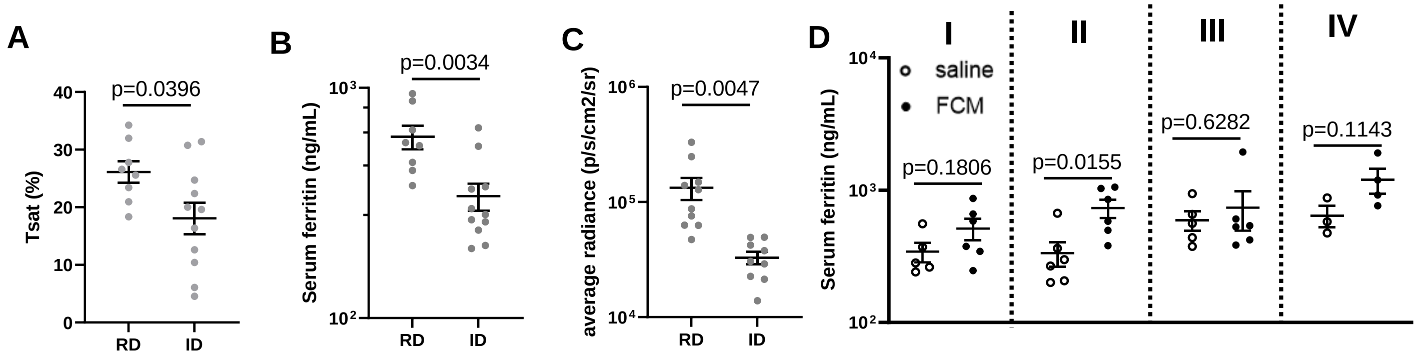

**Supplemental Figure 2- Iron status of iron-replete and iron-deficient mice**
Effects of provision of iron-deficient diet (ID) vs replete diet (RD) to mice on transferrin saturation (A), serum ferritin (B), and myocardial labile iron as assessed by bioluminescence (C). Serum ferritin concentrations in iron-deficient or iron replete mice 1 hour or 6 weeks after infusion of saline or FCM (15mg/kg iron) (D).

**Supplemental Figure 3**

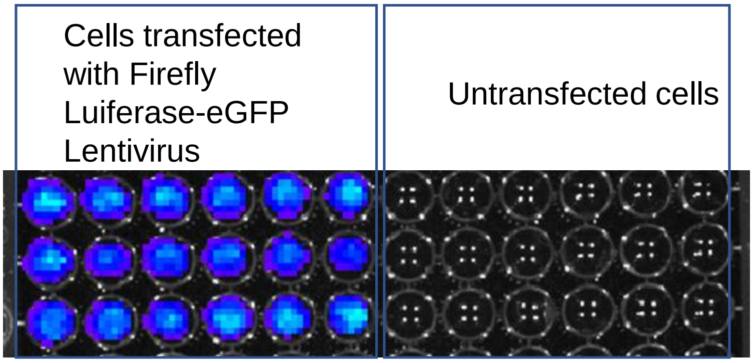

**Supplemental Figure 3-** Confirmation of successful transfection of rat cardiac myocytes with the firefly luciferase gene. D-luciferin substrate was added to transfected and non-transfected cells.

#### Supplemental Figure 4

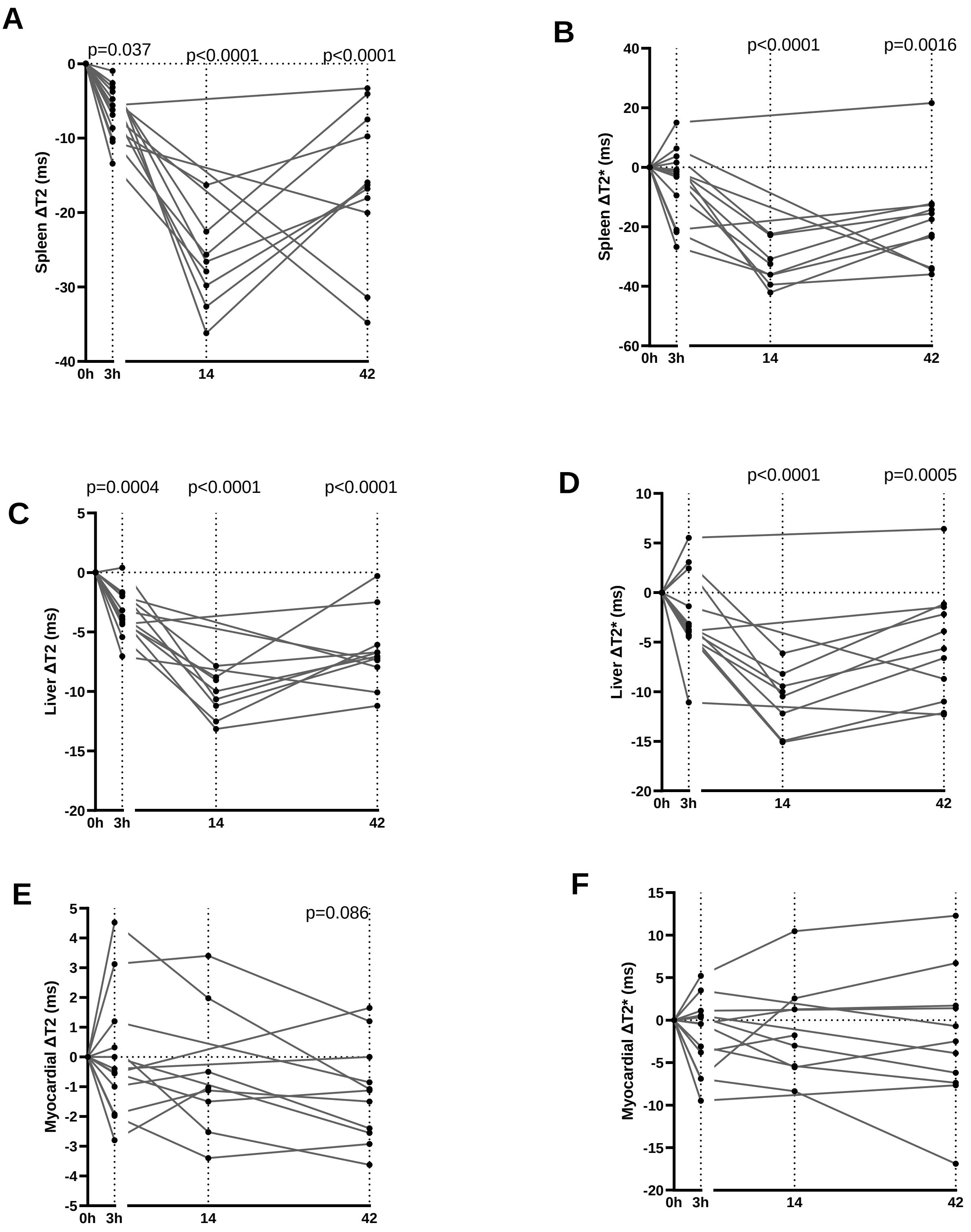

**Supplemental Figure 4- Longitudinal changes in relaxometries ΔT2 and ΔT2* in patients following infusion with 15mg/kg FCM.**

A, B) Longitudinal changes from baseline (Δ) in splenic T2 and T2*. C, D ) Longitudinal changes from baseline (Δ) in liver T2 and T2*. E, F) Longitudinal changes from baseline (Δ) in myocardial T2 and T2*.

**Supplemental table 1**

| | Predicted myocardial T1 (ms) based on 28.45ms drop per dose | | | | | |
|---|---|---|---|---|---|---|
| baseline | after 1 st dose | after 2nd dose | after 3rd dose | after 4th dose | after 5th dose | after 6th dose |
| 998.45 | 970 | 941.55 | 913.1 | 884.65 | 856.2 | 827.75 |

**Supplemental table 1- Predicted cumulative effects of multiple IV iron doses on myocardial T1 (ms).**
Calculations showing predicted cumulative effect of 6 standard doses of IV iron on myocardial T1 based on the observed 28.45ms drop in T1 with every dose.

**Study Title:** **S**tudy of **T**issue Iron **U**ptake in Iron-**D**eficient Patients Receiving Intravenous Iron Replacement Therap**y**: A prospective observational Study (**STUDY**)

Short Title: Imaging Intravenous Iron

**Internal Reference Number / Short title:** Imaging intravenous iron

**Ethics Ref:** 22/NW/0172

**IRAS Project ID:** 308355

**Date and Version No:**24/07/2023, Version 2.0

**Chief Investigator:** Prof Samira Lakhal-Littleton, University of Oxford

**Investigators:**

A/Prof Vanessa Ferreira. BHF Associate Professor of Cardiovascular Medicine, Deputy Director, Oxford Centre for Clinical Magnetic Resonance Research (OCMR), Honorary Consultant Cardiologist.

A/Prof Stefan Piechnik. Head of Advanced Cardiovascular Imaging Analysis, Cardiovascular Imaging Core Laboratory, University of Oxford Centre for Clinical Magnetic Resonance Research (OCMR).

Dr Akshay Shah. NIHR Academic Clinical Lecturer, Specalist Registrar in Anaesthesia & Intensive care, University of Oxford

Dr Michael Desborough. Consultant Haematologist, Department of Clinical Haematology, Oxford University Hospitals NHS Foundation Trust.

Dr Paolo Polzella. Consultant Haematologist, Department of Clinical Haematology, Oxford University Hospitals NHS Foundation Trust.

**Sponsor:** University of Oxford
Joint Research Office
1st floor, Boundary Brook House
Churchill Drive,
Headington,
Oxford, OX3 7GB

**Funder:** British Heart Foundation Centre for Research Excellence,

**Chief Investigator Signature:**

**Statistician Signature:** N/A

The study group declare that there are no potential conflicts of interest.

**Confidentiality Statement**

© Copyright: The University of Oxford and Oxford University Hospitals NHS Foundation Trust 2019

This document contains confidential information that must not be disclosed to anyone other than the Sponsor, the Investigator Team, HRA, host organisation, and members of the Research Ethics Committee, unless authorised to do so.

© Copyright: The University of Oxford and Oxford University Hospitals NHS Foundation Trust 2019

© Copyright: The University of Oxford and Oxford University Hospitals NHS Foundation Trust 2019

21. REFERENCES ... 24
22. APPENDIX A: STUDY FLOW CHART ... 26
23. APPENDIX B: SCHEDULE OF STUDY PROCEDURES ... 27
24. APPENDIX C: AMENDMENT HISTORY ... 28
25. APPENDIX D: STUDY PLAN ... 28

© Copyright: The University of Oxford and Oxford University Hospitals NHS Foundation Trust 2019

#### 1. KEY CONTACTS

| | |
|---|---|
| **Chief Investigator** | Prof Samira Lakhal-Littleton, Department of Physiology, Anatomy and Genetics, Parks Road OX1 3PT, University of Oxford<br><br>Samira<br><br>01865272543 |
| **Sponsor** | University of Oxford<br>Boundary Brook House<br>Churchill Drive,<br>Headington,<br>Oxford, OX3 7GB |
| **Funder(s)** | BHF Centre of Research Excellence<br><br>Division of Cardiovascular Medicine<br><br>Direct line: 01865 234348<br><br> |
| **Clinical Trials Unit** | N/A |
| **Statistician** | N/A |

#### 2. LAY SUMMARY

Iron deficiency is the most common nutritional deficiency in the world. It is now recognized that iron deficiency can affect our health in many ways, impairing brain development and physical function, and worsening co-existing conditions such as heart and kidney disease. It is normally treated with oral iron tablets, which delivers iron gradually in ~65mg doses over a number of weeks. The amount of iron absorbed from these doses is carefully balanced with preventing excess iron uptake into the body. Too much iron can be harmful. In recent years, an alternative treatment based on intravenous iron (i.e. through a 'drip') has become increasingly common. This can be helpful in patients who are unable to take oral iron, or when there is a need for rapid treatement of iron deficiency e.g. before major surgery. In contrast to oral iron, intravenous iron safely delivers a large amount of iron (up to 2g) at once. It works quickly as it is delivered directly into the blood and bypasses the gut. While it is an efficient way of quickly correcting iron deficiency (i.e. restoring blood iron markers to normal values), we do not know where in the body this large quantity of iron ends up. This is an important question, because the human body is not capable of excreting iron. As a result, some of this extra iron and may be deposited in vital organs like the liver, heart and spleen. We know from studying patients and animals with genetic iron overload disorders (e.g. hemochromatosis) that too much iron in these organs can be detrimental to health.

© Copyright: The University of Oxford and Oxford University Hospitals NHS Foundation Trust 2019

The aim of this study is to track where the iron goes in different tissues in the hours, days and weeks after an intravenous iron infusion. We can track iron in tissues by using sophisticated magnetic resonance imaging (MRI). This MRI technique is the gold standard technique for estimating tissue iron levels in patients. It collects certain relaxometry parameters R1/R2/R2* (R2 star refers to a parameter derived from raw MRI data), which are well established as accurate indicators of tissue iron content.This is safe and doesn't involve any additional invasive procedures. We will include patients who have been prescribed intravenous iron as part of standard clinical care to treat their iron deficiency.

The information gained from this study will be used to design a larger study to investigate the potential toxic effects of intravenous iron infusion in different patient populations. Ultimately, the understanding gained from these studies will help us refine current treatments of iron deficiency to maximise the benefits and minimise the harms.

#### 3. SYNOPSIS

| | | | |
|---|---|---|---|
| Study Title | **S**tudy of **Ti**ssue Iron **U**ptake in Iron-**D**eficient Patients Receiving Intravenous Iron Replacement Therap**y**: A prospective observational Study (**STUDY)** | | |
| Internal ref. no. / short title | Imaging intravenous iron | | |
| Study registration | We will register this study on ISRCTN following ethical approval and prior to recruitment of first participant. | | |
| Sponsor | University of Oxford, RGEA<br>Boundary Brook House,<br>Churchill Drive,<br>Headington,<br>Oxford, OX3 7GB | | |
| Funder | BHF Centre of Research Excellence<br>Division of Cardiovascular Medicine<br>Direct line: 01865 234348<br> | | |
| Study Design | Prospective, observational, cohort study | | |
| Study Participants | Age >18 years, scheduled to receive intravenous iron therapy for correction of iron deficiency | | |
| Sample Size | 12 | | |
| Planned Study Period | 01-August-2022 to 31-July-2024 | | |
| | Objectives | Outcome Measures | Timepoint(s) |
| Primary | To establish the kinetics of iron uptake into the heart, liver, spleen, kidney, skeletal muscle and blood | Changes from baseline in cardiac structure and function (Cine MRI) and multi-organ magnetic resonance relaxometries (delta | Baseline, prior to infusion (day 1)<br>Timepoint 1- 3hr post infusion (day 1) |

© Copyright: The University of Oxford and Oxford University Hospitals NHS Foundation Trust 2019

| | | | |
|---|---|---|---|
| | following a single intravenous iron infusion (Ferric carboxymaltose, Ferinject, 1000mg) | R1/R2/R2*) for each participant | Timepoint 2- ~2 weeks post infusion<br>Timepoint 3- ~6 weeks post infusion |
| Secondary | Determine the effects of iron infusion (Ferric carboxymaltose, Ferinject, 1000mg) on serum iron indices and serum markers of tissue iron damage. | Change from baseline in:<br>1. Serum iron indices: iron, ferritin, transferrin saturation, non-transferrin bound iron<br>2. Serum markers of tissue iron damage, including but not limited to lipid peroxidation markers malondialdehyde (MDA) and 4-Hydroxynonenal (HNE) | Baseline, prior to infusion (day 1)<br>Timepoint 1- 3hr post infusion (day 1)<br>Timepoint 2- ~2 weeks post infusion<br>Timepoint 3-~6 weeks post infusion |
| Intervention(s) | None- This is a non-interventional observational study | | |
| Comparator | None | | |

* - refers to a parameter derived from raw MRI data

#### 4. ABBREVIATIONS

| | |
|---|---|
| CI | Chief Investigator |
| CRF | Case Report Form |
| GCP | Good Clinical Practice |
| GP | General Practitioner |
| HRA | Health Research Authority |
| ICF | Informed Consent Form |
| NHS | National Health Service |
| RES | Research Ethics Service |
| PI | Principal Investigator |
| PIL | Participant/ Patient Information Leaflet |
| R&D | NHS Trust R&D Department |
| REC | Research Ethics Committee |
| RGEA | Research Governance, Ethics & Assurance, University of Oxford |
| SOP | Standard Operating Procedure |

© Copyright: The University of Oxford and Oxford University Hospitals NHS Foundation Trust 2019

| | |
|---|---|
| AHF | Acute heart failure |
| CHF | Chronic heart failure |
| I.V | intravenous |
| MRI | Magnetic Resonance Imaging |
| OCMR | Oxford Centre for Magnetic Resonance |
| IDMS | Iron Deficiency Management Service |
| FCM | Ferric carboxymaltose |
| NTBI | Non-tramsferrin bound iron |
| OUHFT | Oxford University Hospitals NHS Foundation Trust |

#### 5. BACKGROUND AND RATIONALE

**Iron deficiency**-The prevalence of non-anaemic iron deficiency is 1-2 billion worldwide[1]. It has recently become recognised as an important co-morbidity in common conditions including acute and chronic heart failure (AHF and CHF)[2,3]. Traditionally, iron deficiency has been treated through oral iron supplementation.
**New treatment for iron deficiency-** The strategy for treating iron deficiency has been changed by our growing understanding of iron homeostasis. It is now recognised that the iron-homeostatic hormone hepcidin (which is raised in inflammatory conditions such as ACF and CHF) contributes to iron deficiency by blocking the sole mammalian iron exporter ferroportin in the gut and spleen, respective sites of iron absorption and recycling[3,4]. These discoveries have driven the development, and now widespread use, of intravenous (I.V) iron (in-lieu of oral iron) to bypass blocked ferroportin in the gut[5]. Some short-term trials have shown modest or no benefits of I.V iron on exercise capacity, risk of hospitalisation and of cardiovascular death in CHF and AHF[6,7]. Despite this, the European Society of Cardiology (ESC) now recommends the use of I.V iron in all HF patients with iron deficiency[8]. I.V iron is also increasingly being used for correction of iron deficiency in otherwise healthy individuals, as well as in critical care setting. Consequently, NHS expenditure on I.V iron has grown exponentially in recent years. However, there is a scarcity of data on the long-term effects and the mechanisms of action of I.V iron, and persisting safety concerns over increased risk of infection and organ iron toxicity.

**Key question with safety implications**- We do not know how I.V iron formulations affect myocardial iron levels and cardiac function in the long term. Answering these questions is important for ensuring that correction of iron deficiency is not accompanied by myocardial iron toxicity. It is widely assumed that I.V iron formulations such as Ferinject (ferric carboxymaltose or FCM) are taken up and degraded by macrophages, allowing slow and controlled release of iron into the circulation, and subsequent loading onto the plasma iron chaperone transferrin for uptake by tissues. Most tissues can prevent excess iron accumulation by downregulating the levels of transferrin receptor expression (this is achieved through the iron-sensing function of iron-regulatory proteins IRPs)[9]. However, kinetic studies of plasma iron indices, together with the speed of manifestation of associated physiological effects demonstrate that FCM infusion results in rapid and physiologically-relevant rise in non-transferrin bound iron (NTBI) in the plasma[10-12]. Excess NTBI in the plasma is dangerous because it is taken up by tissues through L-type and T-type calcium channels, which are not regulated by IRPs[13]. These channels are highly expressed in the heart and it is well established that they are responsible for the iron overload cardiomyopathy that occurs in genetic disorders of iron overload such as hemochromatosis (where NTBI is also raised)[14]. In pre-clinical mouse models, we found that a patient-equivalent dose of intravenous iron resulted in ~400% increase in cardiac iron levels[15].

© Copyright: The University of Oxford and Oxford University Hospitals NHS Foundation Trust 2019

**Study rationale** - This is a pump-priming study to establish if the standard intravenous iron treatment, given as part of standard clinical care to correct iron deficiency, results in an acute rise in tissue iron levels, including myocardial iron at the early timepoints, which would be evidence for the direct, rapid unregulated route of iron uptake. It is an observational cohort study measuring changes in tissue iron levels from baseline following infusion of intravenous iron.Tissue iron levels will naturally vary between individuals. Because of this, the study will not use a placebo arm. Instead, it will measure changes against baseline tissue iron levels.

**Value** -This work is a first and essential step towards understanding the effects of this emerging iron replacement therapy on tissue iron levels, and identifying the features that distinguish patients who will benefit from those who could be harmed by I.V iron treatment.

#### 6. OBJECTIVES AND OUTCOME MEASURES

| Objectives | Outcome Measures | Timepoint(s) of evaluation of this outcome measure (if applicable) |
|---|---|---|
| **Primary Objective**<br>To establish the kinetics of iron uptake into the heart, liver, spleen, kidney, skeletal muscle and blood following a single intravenous iron infusion (Ferric carboxymaltose, Ferinject, 1000mg) | Changes from baseline in cardiac structure and function (Cine MRI) and multi-organ magnetic resonance relaxometries (delta R1/R2/R2*) for each participant | Baseline, prior to infusion (day 1)<br>Timepoint 1- 3hr post infusion (day 1)<br>Timepoint 2- ~2 weeks post infusion<br>Timepoint 3- ~6 weeks post infusion |
| **Secondary Objective**<br>Determine the effects of iron infusion (Ferric carboxymaltose, Ferinject, 1000mg) on serum iron indices and serum markers of tissue iron damage. | Change from baseline in:<br>1. Serum iron indices: iron, ferritin, transferrin saturation, non-transferrin bound iron<br>2. Serum markers of tissue iron damage, including but not limited to lipid peroxidation markers malondialdehyde (MDA) and 4-Hydroxynonenal (HNE) | Baseline, prior to infusion (day 1)<br>Timepoint 1- 3hr post infusion (day 1)<br>Timepoint 2- ~2 weeks post infusion<br>Timepoint 3-~6 weeks post infusion |

© Copyright: The University of Oxford and Oxford University Hospitals NHS Foundation Trust 2019

#### 7. STUDY DESIGN

This is a prospective, observational, study exploring the kinetics of tissue iron uptake following intravenous iron infusion (Ferinject), received as part of standard clinical care in a group of iron-deficient patients.

Participants will be recruited through the Iron Deficiency Management Service, part of the Oxford University Hospitals NHS Foundation Trust (OUHFT) .

Participants will receive their intravenous iron infusion (Ferinject) as part of their standard clinical care by an NHS clinician. They will undergo all the study MRI scans and additional study procedures at the Oxford Centre for Clinical Magnetic Resonance Research (OCMR), which is based at the John Radcliffe Hospital and is part of the Division of Cardiovascular Medicine within the Radcliffe Department of Medicine at the University of Oxford.

A study flowchart is shown in Appendix A.

**Data and Sample Collection**

***MRI –*** MRI scans of the target organs of interest (such as heart, liver, spleen, kidney, blood and skeletal muscle) will be undertaken in OCMR at the JR Hospital, using established methods for cardiac Cine MRI and for measuring R1, R2 and R2* relaxometries for tissues.

***Peripheral blood samples –*** 20 ml (~4 teaspoons) of blood will be collected by a nurse for study investigations. A seprate sample collected as part of standard clinical care will be obtained. Serum from study blood samples will be extracted and stored at -80C in OCMR for all participants. At the end of the study, study serum samples will be transported to the Lakhal-Littleton Lab at the Department of Physiology, Anatomy and Genetics (DPAG), Part of the Medical Sciences Division of the University of Oxford (see section 9.10).

#### 8. PARTICIPANT IDENTIFICATION

##### 8.1. Study Participants

- Participant is willing and able to give informed consent for participation in the study.
- Aged 18 years or above.
- Anaemia (haemoglobin less than 120g/L for women and less than 130g/L for men) and/or confirmed iron deficiency (ferritin less than 100mcg/L and/or transferrin saturation less than 20%).
- Scheduled to receive intravenous iron (Ferinject) for correction of iron deficiency.

##### 8.2. Exclusion Criteria

The participant may not enter the study if ANY of the following apply:

- Any MRI incompatible implants (e.g. cardiac, neuro, ocular implants, surgical clips, aneurysm clips, shrapnel/bullets)
- Pregnant or lactating participants

© Copyright: The University of Oxford and Oxford University Hospitals NHS Foundation Trust 2019

- Acute decompensated heart failure
- Unstable clinical status
- Any other medical conditions which would influence the reliability of the study results determined by the investigators.
- Any other contraindication to MRI to be confirmed by the qualified MRI operator, e.g. tattoos containing traces of metal.

#### 9. PROTOCOL PROCEDURES

A schedule of procedures is shown in Appendix B.

##### 9.1. Recruitment

Participants will be recruited from referrals to the Iron Deficiency Management Service, part of Oxford University Hospitals NHS Foundation Trust (OUHFT). This service receives referrals from multiple sources across Oxfordshire. Patient referrals will be screened by members of the NHS Clinical team at IDMS to identify potentially eligible participants. For further details see section 9.2 below.

##### 9.2. Screening and Eligibility Assessment

Eligible participants will be enrolled only if this would not cause any delay to their care.

**Screening procedure**

Patients referred to the IDMS are typically anaemic (haemoglobin less than 120g/L for women or 130g/L for men) and will have a ferritin less than 100 ug/L and/or transferrin saturation less than 20%. These tests are carried out as part of routine clinical care. All referrals to IDMS will be screened by a member of NHS clinical care team. Patients who would require intravenous iron and who are deemed eligible to participate in the study will be sent an invitation letter and patient information sheet. Few days later, a member of the NHS clinical care team at IDMS will contact the patient by telephone to check they have received the information and are happy to be contacted further about the study by a member of the study team. If they are, a member of the study team will telephone the patient to explain the study, and answer any questions the patients might have and check their eligibility. They will also ask the patient for the verbal consent to participate in the study. If verbal consent is received, the member of the study team will then arrange a suitable date for the participant to receive their intravenous iron infusion as part of their standard clinical care, normally within days and proceed with the study as described below.

##### 9.3. Informed Consent

A member of the study team will make telephone contact with eligible patients who have agreed to be contacted about this study. They will detail the exact nature of the study; what it will involve for the participant; the implications and constraints of the protocol; the known side effects and any risks involved in taking part. They will also clearly state that the participant is free to withdraw from the study at any time for any reason without prejudice to future care, without affecting their legal rights, and with no obligation to give the reason for withdrawal. The participant will be allowed as much time as wished

© Copyright: The University of Oxford and Oxford University Hospitals NHS Foundation Trust 2019

to consider the information, and the opportunity to question the member of study team, their GP or other independent parties to decide whether they will participate in the study. Verbal consent will be taken at this stage and recorded in the CRF and the patient's medical notes. Once verbal consent is given, the member of the NHS clinical care team at IDMS will arrange for the participant to be given their intravenous iron treatment, as per standard clinical care, by a member of the NHS clinical care team at the OCMR, which is part of the Oxford University Medical Sciences Division, and is physically located on the John Radcliffe Hospital site.

Written version of the Informed Consent form will be presented to the participant when they attend OCMR on day 1 of the study. This will be done by a suitably qualified and experienced member of the study team that has been authorised to do so by the Chief/Principal Investigator. Again, this member of study team will detail the exact nature of the study; what it will involve for the participant; the implications and constraints of the protocol; the known side effects and any risks involved in taking part. They will also clearly state that the participant is free to withdraw from the study at any time for any reason without prejudice to future care, without affecting their legal rights, and with no obligation to give the reason for withdrawal.

The participant must personally sign and date the latest approved version of the Informed Consent form before any study specific procedures are performed. One copy of the signed Informed Consent will be given to the participant and another copy will be added to the participant's medical notes. The original signed form will be retained at the study site.

#### 9.4. Enrolment

This is a non-interventional observational study and as such does not involve randomisation. Enrolled participants will be allocated a unique study identification number.

#### 9.5. Blinding and code-breaking

There is no blinding in this study.

#### 9.6. Description of study intervention(s), comparators and study procedures (clinical)

##### 9.6.1. Description of study intervention(s)

There are no interventions as part of this study. Intravenous iron infusion (Ferinject) is delivered as part of standard clinical care.

##### 9.6.2. Description of study procedure(s)

***Magnetic Resonance Imaging***

**Overview**

MRI will be performed on clinical standard MRI scanners in OCMR (at conventional field strength 1.5 Tesla for this study). Participants will be grouped into 4 cohorts of 3, so that all participants in the same cohorts are scanned on the same days. Details of how cohorts will be run are shown in section 9.7 and in Appendix D. Each scan will consist of MRI scanning techniques to assess cardiac structure and function by Cine MRI and organ tissue characterisation using R1/R2/R2* relaxometry.

© Copyright: The University of Oxford and Oxford University Hospitals NHS Foundation Trust 2019

The average duration of each MRI scan is envisaged to be around an hour. One participant in each cohort will be chosen to have an extended MRI scan at their first visit, lasting up to 3 hours. This extended MRI scan is needed to optimize the parameters for running of subsequent scans that day. All subsequent scans will be a standard duration of about an hour. This will be communicated to the participants clearly in the PIL and again on day 1 when they attend OCMR for their first scan.

**MRI safety**

MRI is safe and non-invasive and does not involve any ionising radiation (x-rays). However, there are some contraindicatios. Because of this, participants will be asked pre-screening questions at the recruitment stage by a member of the study team, and again by the radiographer when they attend OCMR on day 1, prior to any procedures. Screening questions will check that the particpant is not pregnant, or suspected to be pregnant, does not have claustrophobia, or carry heart pacemaker, mechanical heart valve, mechanical implant such as an aneurysm clip, hip replacement, or any other pieces of metal that have accidentally entered the body. Additionally, some types of tattoo ink contains traces of metal, but most tattoos are safe in an MRI scanner. Participants will be asked to to change into pocketless and metal free "pyjama-style" top and trousers, available in a range of sizes, and will also be asked to remove underwired bras, metal jewellery, body piercings, and make up where applicable. Lockers are provided to secure their personal belongings and clothing. During the scan, participants will additionally be given earplugs, head padding or headphones to reduce the noise perception from the MRI scan, and will have a call button in their hands to alert the radiographer immediately if they feel heat in their tattoo, or any other general discomfort.

The OCMR is fully equipped for resuscitation (including defibrillation) in the unlikely event of a medical emergency during scanning, and doctors performing and/or supervising the scans are trained in magnetic safety, evacuation procedures, and either Basic Life Support (BLS), Intermediate Life Support (ILS) or Advanced Life Support (ALS).

***Research sample taking***

A total of 5 venous blood samples will be collected from each participant, 1 as part of their standard clinical care, and 4 for study investigations**.** Each time, a qualified nurse will collect around 5ml (1 teaspoon) of blood. Serum from study blood samples will be extracted and stored in a secure research -80C freezer in OCMR for all participants. At the end of the study, study serum samples will be transported to the Lakhal-Littleton Lab at the Department of Physiology, Anatomy and Genetics (DPAG), Part of the Medical Sciences Division of the University of Oxford for further investigational assays.

**Investigational assays** - Serum samples derived from all participants and all timepoints will be tested together. Tests include measurements of serum iron indices (iron, ferritin, transferrin and transferrin saturation) using automated analytical Chemistry instrument (Pentra C400) in the Lakhal-Littleton Lab. Markers of tissue iron damage (HNE and MDA) will also be measured in serum samples using standard commercially available kits (Abcam ab118970 and ab238538 respectively).

#### 9.7. **Day1 - Baseline Assessments**

© Copyright: The University of Oxford and Oxford University Hospitals NHS Foundation Trust 2019

Example Study MRI schedule: Cohort 1 (3 participants per cohort)*

| **Day 1 - Baseline & 3h post IV iron infusion** | **2 weeks (14+/-2days) - post IV iron infusion** | **6 weeks (42+/-2days) - post IV iron infusion** |
|---|---|---|
| 9:00 am - Participant #1 baseline (3 hrs)<br>12:00 pm - Participant #2 baseline (1 hr)<br>1:00 pm - Participant #3 baseline (1 hr)<br>2:00 pm – BREAK<br><br>3:00 pm – Participant#1 at 3hr-post iron (1 hr)<br>4:00 pm – Participant #2 at 3hr-post iron (1 hr)<br>5:00 pm - Participant #3 at 3hr-post iron (1 hr)<br>6:00 pm – END | 9:00 am - Participant #1 (1hr)<br>10:00 am - Participant #2 (1hr)<br>11:00 am - Participant #3 (1hr)<br>12:00 pm - END | 9:00 am - Participant #1 (1hr)<br>10:00 am - Participant #2 (1hr)<br>11:00 am - Participant #3 (1hr)<br>12:00 pm - END |

** Repeat for Cohort 2 and Cohort 3, Cohort4.*

Baseline clinical data for each Participant:

- Confirmation against inclusion and exclusion criteria

Baseline MRI scan (for Participant #1 in each cohort; up to 3 hours)

- Cine imaging forcardiac structure and function
- Tissue characterisation by R1/R2/R2* relaxometry of the heart, liver, spleen, blood and skeletal muscle over 3 hours.

Baseline MRI scan (for Participant #2 and #3 in each cohort; up to 1 hour)

- Cine imaging forcardiac structure and function
- Tissue characterisation by R1/R2/R2* relaxometry of the heart, liver, spleen, blood and skeletal muscle over 1 hour.

Baseline laboratory data:

- In addition the research team memeber will collect participant's demographic information and medical history from their medical notes.
- Haemoglobin, iron, transferrin saturation and ferritin data available from medical notes.

Baseline blood samples:

- Two collected, one as part of standard clinical care, and the other for study investigations to measure serum iron indices and serum markers of tissue iron damage.

Intravenous iron infusion:

- Administered by a member of the clinical care team as part of standard clinical care

#### 9.8. **Day 1 - Three-hour post IV iron infusion assessment**

MRI scan (up to 1 hour)

© Copyright: The University of Oxford and Oxford University Hospitals NHS Foundation Trust 2019

- Cine imaging for cardiac structure and function
- Tissue characterisation by R1/R2/R2* relaxometry of the heart, liver, spleen, blood and skeletal muscle

Blood samples:

- One sample collected for study investigations to measure serum iron indices and serum markers of tissue iron damage.

#### 9.9. **Subsequent Visits**

##### 9.9.1. Two weeks (14 days+/-2 days) follow up visit

MRI scan (up to 1 hour)

- Cine imaging for cardiac structure and function
- Tissue characterisation by R1/R2/R2* relaxometry of the heart, liver, spleen, blood and skeletal muscle

Blood samples:

- One sample collected for study investigations to measure serum iron indices and serum markers of tissue iron damage.

##### 9.9.2. Six weeks (42 days+/- 2 days) follow up visit

MRI scan (up to 1 hour)

- Cine imaging for cardiac structure and function
- Tissue characterisation ny R1/R2/R2* relaxometry of the heart, liver, spleen, blood and skeletal muscle

Blood samples:

- One sample collected for study investigations to measure serum iron indices and serum markers of tissue iron damage.

#### 9.10. **Sample Handling**

Study specific blood samples will be taken twice on day 1 (baseline assessment, 3 hour assessment), once on each of the follow up visits (2 weeks and 6 weeks). The total volume of blood will be

© Copyright: The University of Oxford and Oxford University Hospitals NHS Foundation Trust 2019

approximately 20 mls (5ml each time). Blood samples will be processed immediately at OCMR by a member of the research team to extract serum.

Serum samples labelled only with the participants' number/code will initially be stored in a secure research -80C freezer at OCMR. After the last sample from the last participant has been collected, all the samples will be shipped on dry ice to the Lakhal-Littleton Lab at the Department of Physiology Anatomy and Genetics, part of the Medical Sciences Division at the Universiy of Oxford. These samples will be kept in a secure, locked -80 freezer until they are ready to be analysed. Here, samples will be tested together as a single batch for iron indices and markers of tissue iron injury (MDA and 4-HNE) as described above.

Any surplus sample will be stored for potential future ethically approved studies in the UK at the end of the study, subject to participant agreeing to this at the consent stage. Otherwise, after completing the sample analysis for the purpose of the study, any remaining samples will be destroyed. For participants who agree to their to their samples being used in future research, the consent form will be held until the samples have been used up. Only the study team and other delegated members specified in the investigator site file will have access to the samples. A dedicated SOP for sample handling and processing will be followed.

#### 9.11. **Early Discontinuation/Withdrawal of Participants**

During the course of the study a participant may choose to withdraw early from the study treatment at any time. This may happen for several reasons, including but not limited to:

- The occurrence of what the participant perceives as an intolerable AE.
- Inability to comply with study procedures
- Participant decision

Participants who withdraw on day 1 will still receive their intravenous iron infusion as per standard clinical care, but no further data or blood will be collected for the purposes of the study. Any data or study blood sample collected up to the point of withdrawal will be retained for use in the study analysis, unless the participant specifically requests that they are destroyed. No further data or samples would be collected after withdrawal.

In addition, the Investigator may discontinue a participant from the study at any time if the Investigator considers it necessary for any reason including, but not limited to:

- Ineligibility (either arising during the study or retrospectively having been overlooked at screening)
- Significant protocol deviation
- Clinical decision

Participants who withdraw before completing day 1 will be replaced with newly recruited eligible participants. Participants who withdraw after completing day 1 will not be replaced.

The type of withdrawal and reason for withdrawal will be recorded in the Case Report Form (CRF).

© Copyright: The University of Oxford and Oxford University Hospitals NHS Foundation Trust 2019

##### 9.12. **Definition of End of Study**
The end of study will be when the last sample from the last recruit has been analysed.

#### 10. SAFETY REPORTING

##### 10.1. **Definition of Serious Adverse Events**
A serious adverse event is any untoward medical occurrence that:

- results in death
- is life-threatening
- requires inpatient hospitalisation or prolongation of existing hospitalisation
- results in persistent or significant disability/incapacity
- consists of a congenital anomaly or birth defect.

Other ‘important medical events’ may also be considered a serious adverse event when, based upon appropriate medical judgement, the event may jeopardise the participant and may require medical or surgical intervention to prevent one of the outcomes listed above.

NOTE: The term "life-threatening" in the definition of "serious" refers to an event in which the participant was at risk of death at the time of the event; it does not refer to an event which hypothetically might have caused death if it were more severe.

##### 10.2. **Reporting Procedures for Serious Adverse Events**
A serious adverse event (SAE) occurring to a participant should be reported to the REC that gave a favourable opinion of the study where in the opinion of the Chief Investigator the event was ‘related’ (resulted from administration of any of the research procedures) and ‘unexpected’ in relation to those procedures. Reports of related and unexpected SAEs should be submitted within 15 working days of the Chief Investigator becoming aware of the event, using the HRA report of serious adverse event form (see HRA website).

#### 11. STATISTICS AND ANALYSIS
The plan for the statistical analysis of the study are outlined below. There is not a separate SAP document in use for the trial.

##### 11.1. **Description of the Statistical Methods**
The primary outcome is the absolute change from baseline in MR relaxivities R1, R2, R2* at each timepoint post Ferinject infusion, according to the following formulae:

ΔR1=R1 (at timepoint x)-R1(at baseline)

ΔR2=R2 (at timepoint x)-R2(at baseline)

ΔR2*=R2* (at timepoint x)-R2*(at baseline)

© Copyright: The University of Oxford and Oxford University Hospitals NHS Foundation Trust 2019

R1, R2, R2* values are calculated as inverse of T1,T2, T2* relaxation times obtained by the MR relaxometry or mapping techniques. The results will be reported as time series of average values and standard deviations (median and IQR for nonparametric variables) at each timepoint post Ferinject infusion. The presence of statistically significant signal will be assessed using ANOVA.

The secondary outcome measure is the change from baseline in serum concentrations of iron indices (iron, transferrin saturation, ferritin and non-transferrin bound iron) and markers of tissue iron toxicity (MDA ad 4-HNE). Results will be expressed both as absolute change from baseline and as percentage changerelative to baseline.

Further exploratory analyses will be preformed for possible interactions between the observed measures using correlations, crosscorrelations and pharmacodynamic modelling.

##### 11.2. Sample Size Determination

The sample size of 12 participants was not arrived at statistically, due to lack of information on the magnitude of effects of Ferinject on R1/R2/R2* values at early timepoints. However, an exploratory study of MRI imaging following intravenous infucion with ultrasmall superparamagnetic particles of iron oxide (USPIO) at later timepoints used sample sizes of 5-12 participants per group[16]. Of note, the longitudinal design involving repeated measures will increase the statistical power of the study.

##### 11.3. Analysis populations

All participants will be included in the analysis as enrolled.

##### 11.4. Decision points

After the first cohort (3 participants), we will conduct an interim analysis to ensure appropriateness of chosen timepoints.

##### 11.5. Stopping rules

There are no formal stopping rules for this study.

##### 11.6. The Level of Statistical Significance

Tests will be two-sided and considered to provide evidence for a significant difference if p-values are less than 0.05.

##### 11.7. Procedure for Accounting for Missing, Unused, and Spurious Data.

Reasons for missing data, loss to follow-up and participant withdrawals will be carefully considered, reported and assessment for that type of random 'missingness' will be made. Missing data will be minimised by collecting the minimum amount of data required. All data queries will be resolved prior to analysis.

© Copyright: The University of Oxford and Oxford University Hospitals NHS Foundation Trust 2019

##### 11.8. Procedures for Reporting any Deviation(s) from the Original Statistical Plan

Deviations are not anticipated. The analysis outline in this protocol will be followed. Any deviations from this will be reported in the final study report.

##### 11.9. Health Economics Analysis

There are no planned health economic analyses.

#### 12. DATA MANAGEMENT

The plan for the data management of the study are outlined below. There is not a separate Data Management document in use for the study.

##### 12.1. Source Data

Source documents are where data are first recorded, and from which participants' CRF data are obtained. These include, but are not limited to, hospital records (from which medical history and previous and concurrent medication may be summarised into the CRF), clinical and office charts, laboratory and pharmacy records, diaries, microfiches, radiographs, and correspondence.

CRF entries will be considered source data if the CRF is the site of the original recording (e.g. there is no other written or electronic record of data). All documents will be stored safely in confidential format. On all study-specific documents, other than the signed consent, the participant will be referred to by the study participant number/code, not by name.

##### 12.2. Access to Data

Direct access will be granted to authorised representatives from the Sponsor and host institution for monitoring and/or audit of the study to ensure compliance with regulations.

##### 12.3. Data Recording and Record Keeping

All study data will be entered on a research database platform – 'RedCAP', an internationally recognised standard for study database management. This will be hosted on a secure server managed by the University of Oxford. Participants will be identified by a unique trial specific number and/or code in any database. The name and any other identifying detail will NOT be included in any trial data electronic file.

The CI will be data custodian for a separate master list that will link the study ID and participant personal data. This will include participant contact details which will be used to arrange follow up visits during the study. This master list will be kept and maintained securely on a protected server, separate from CRF data. The following patient identifiers will be used – patient name, date of birth, NHS number and hospital number.

Personal data will be held for a maximum of 12 months after the study has ended. Contact details for participants who have consented to be informed about future relevant studies will be retained separately from this study on a password protected computer in DPAG. They will be removed at any point at participants' request. All research data will be anonymised and will be stored for up to 5 years.

© Copyright: The University of Oxford and Oxford University Hospitals NHS Foundation Trust 2019

For partcipants who consent for their samples to be used in future research, a copy of the consent form will be retained until the samples have been depleted or destroyed.

#### 13. The QUALITY ASSURANCE PROCEDURES

The study may be monitored, or audited in accordance with the current approved protocol, GCP, relevant regulations and standard operating procedures.

##### 13.1. **Risk assessment**

No formal risk assessment will be undertaken

##### 13.2. **Study monitoring**

No GCP monitoring will be undertaken, as this is not a CTIMP.

##### 13.3. **Study Committees**

It is not anticipated that study committees will be convened for these studies.

#### 14. PROTOCOL DEVIATIONS

A study related deviation is a departure from the ethically approved study protocol or other study document or process (e.g. consent process or administration of study intervention) or from Good Clinical Practice (GCP) or any applicable regulatory requirements. Any deviations from the protocol will be documented in a protocol deviation form and filed in the study master file.

#### 15. SERIOUS BREACHES

A “serious breach” is a breach of the protocol or of the conditions or principles of Good Clinical Practice which is likely to affect to a significant degree –

(a) the safety or physical or mental integrity of the trial subjects; or

(b) the scientific value of the research.

In the event that a serious breach is suspected the Sponsor must be contacted within 1 working day. In collaboration with the C.I., the serious breach will be reviewed by the Sponsor and, if appropriate, the Sponsor will report it to the approving REC committee and the relevant NHS host organisation within seven calendar days.

#### 16. ETHICAL AND REGULATORY CONSIDERATIONS

##### 16.1. **Declaration of Helsinki**

The Investigator will ensure that this study is conducted in accordance with the principles of the Declaration of Helsinki.

© Copyright: The University of Oxford and Oxford University Hospitals NHS Foundation Trust 2019

##### 16.2. Guidelines for Good Clinical Practice

The Investigator will ensure that this study is conducted in accordance with relevant regulations and with Good Clinical Practice.

##### 16.3. Approvals

Following Sponsor approval the protocol, informed consent form, participant information sheet and any proposed advertising material will be submitted to an appropriate Research Ethics Committee (REC), and HRA (where required) and host institutions for written approval.

The Investigator will submit and, where necessary, obtain approval from the above parties for all substantial amendments to the original approved documents.

##### 16.4. Other Ethical Considerations

The risks of collecting study boold samples are minimal. This study requires the collection of a small volume of blood without detriment to the participant's medical condition.

Research scans are performed primarily for research purposes, and are not routinely checkedby clinicians for diagnoses or for screening. They are not suitable for diagnosing or screening for disease, such as early detection of cancers, etc. Incidental findings on research scans are handled by OCMR SOP (outlined below).

In the event of seeing any structural abnormalities on a scan (e.g. by a radiographer or a researcher), the scan will be checked by a clinical specialist. If the specialist feels that the abnormality was medically important, they will discuss the implications with the participant and arrange for further investigations as necessary. Participants will not be informed unless the clinical specialistconsiders the finding has clear implications for their current or future health. .

For patients who have been diagnosed and undergoing treatment for cancer, they would have undergone comprehensive staging, including CT and/or MRI or other types of clinical scans, by their oncologists and clinical care team. As such,these patients typically will have been informed of their clinical status with regard to their cancer diagnosis and extent of involvement (e.g. any metastatic disease, or not). Reporting of extra-cardiac incidental findings based on these research scans has the very real potential of causing unnecessary anxiety, extra and duplicate diagnostic tests for these participants who already had undergone/will undergo comprehensive clinical staging as part of their cancer treatment. As such, any incidental findings on the research CMR scans will be limited to reporting of cardiac incidental findings.

##### 16.5. Reporting

The CI shall submit once a year throughout the study, or on request, an Annual Progress report to the REC Committee, HRA (where required) host organisation, Sponsor and funder (where required). In addition, an End of Study notification and final report will be submitted to the same parties.

##### 16.6. Transparency in Research

© Copyright: The University of Oxford and Oxford University Hospitals NHS Foundation Trust 2019

In line with good practice guidelines set out by the HRA, we will register our study on the International Standard Randomised Controlled Trial Number (ISRCTN) Registry, which is a publically accessible registry recongised by the World Health Organisation and by International Committee of Medical Journal Editors.

##### 16.7. Participant Confidentiality

The study will comply with the UK General Data Protection Regulation (UK GDPR) and Data Protection Act 2018, which require data to be de-identified as soon as it is practical to do so. The processing of the personal data of participants will be minimised by making use of a unique participant study number only on all study documents and any electronic database(s), with the exception of the CRF, where participant initials may be added. All documents will be stored securely and only accessible by study staff and authorised personnel. The study staff will safeguard the privacy of participants' personal data.

##### 16.8. Expenses and Benefits

Participants will be compensated for their time and travel expenses, to a total £500 if they complete this study. They will receive £100 after their first visit, £100 after their second visit, and £300 immediately after their third and final visit.

#### 17. FINANCE AND INSURANCE

##### 17.1. Funding

Funding for this study has been secured from the British Heart Foundation Centre for Research Excellence (BHF-CRE). Any additional costs relating to the CI's time beyond the current end date will be covered from the CI's existing funding. Letter of support from the CI's Department is enclosed.

##### 17.2. Insurance

The University has a specialist insurance policy in place which would operate in the event of any participant suffering harm as a result of their involvement in the research (Newline Underwriting Management Ltd, at Lloyd's of London). NHS indemnity operates in respect of the clinical treatment that is provided.

##### 17.3. Contractual arrangements

Appropriate contractual arrangements will be put in place with all third parties.

#### 18. PUBLICATION POLICY

Study reaults will be published in peer-reviewed scientific and medical journals and presented at research conferences. In all cases, participant information will be provided in an anonymised manner. The investigators will be involved in reviewing drafts of the manuscripts, abstracts, press releases and any other publications arising from the study. Authors will acknowledge that the study was funded by the British Heart Foundation Centre for Research Excellence. Authorship will be determined in

© Copyright: The University of Oxford and Oxford University Hospitals NHS Foundation Trust 2019

accordance with the ICMJE guidelines and other contributors will be acknowledged. Upon request, a summary of the study results will be provided to participants by a member of the study team.

#### 19. DEVELOPMENT OF A NEW PRODUCT/ PROCESS OR THE GENERATION OF INTELLECTUAL PROPERTY

Ownership of IP generated by employees of the University vests in the University. The University will ensure appropriate arrangements are in place as regards any new IP arising from the trial. .

#### 20. ARCHIVING

Data will be stored in a secure server after the study has ended. Paper data will the stored in the Trial Master File in secure locker access in the Department of Physiology, Anatomy and Genetics, University of Oxford. Anonymised research data will be kept for a minimum of 5 years.

#### 21. REFERENCES

1. WHO , The Global Prevalence of Anaemia in 2011 (World Health Organization, Geneva, 2015).

2. Comín-Colet J, Enjuanes C, González G, Torrens A, Cladellas M, Meroño O, Ribas N, Ruiz S, Gómez M, Verdú JM, Bruguera J. Iron deficiency is a key determinant of health-related quality of life in patients with chronic heart failure regardless of anaemia status. European Journal of Heart Failure. 2013;15:1164–1172. doi: 10.1093/eurjhf/hft083.

3. Jankowska EA, Kasztura M, Sokolski M, Bronisz M, Nawrocka S, Oleśkowska-Florek W, Zymliński R, Biegus J, Siwołowski P, Banasiak W, Anker SD, Filippatos G, Cleland JG, Ponikowski P. Iron deficiency defined as depleted iron stores accompanied by unmet cellular iron requirements identifies patients at the highest risk of death after an episode of acute heart failure. European Heart Journal. 2014;35:2468–2476.

4. Donovan A, Lima CA, Pinkus JL, Pinkus GS, Zon LI, Robine S, Andrews NC. The iron exporter ferroportin/Slc40a1 is essential for iron homeostasis. Cell Metabolism. 2005;1:191–200.

5. Nemeth E, Tuttle MS, Powelson J, Vaughn MB, Donovan A, Ward DM, Ganz T, Kaplan J. Hepcidin regulates cellular iron efflux by binding to ferroportin and inducing its internalization. Science. 2004;306:2090–2093.

6. Nemeth E, Ganz T. Anemia of inflammation. Hematology/Oncology Clinics of North America. 2014;28:671–681.

7. Anker SD, Comin Colet J, Filippatos G, Willenheimer R, Dickstein K, Drexler H, Lüscher TF, Bart B, Banasiak W, Niegowska J, Kirwan BA, Mori C, von Eisenhart Rothe B, Pocock SJ, Poole-Wilson PA, Ponikowski P, FAIR-HF Trial Investigators Ferric carboxymaltose in patients with heart failure and iron deficiency. New England Journal of Medicine. 2009;361:2436–2448.

8. Ponikowski P, van Veldhuisen DJ, Comin-Colet J, Ertl G, Komajda M, Mareev V, McDonagh T, Parkhomenko A, Tavazzi L, Levesque V, Mori C, Roubert B, Filippatos G, Ruschitzka F, Anker SD, CONFIRM-HF Investigators Beneficial effects of long-term intravenous iron therapy with ferric

© Copyright: The University of Oxford and Oxford University Hospitals NHS Foundation Trust 2019

carboxymaltose in patients with symptomatic heart failure and iron deficiency† European Heart Journal. 2015;36:657–668. doi: 10.1093/eurheartj/ehu385.

9. Lam CSP, Doehner W, Comin-Colet J; IRON CORE Group. Iron deficiency in chronic heart failure: case-based practical guidance. ESC Heart Fail. 2018 Oct;5(5):764-771.

10. Rouault TA. The role of iron regulatory proteins in mammalian iron homeostasis and disease. Nature Chemical Biology. 2006;2:406–414.

11. Garbowski MW, Bansal S, Porter JB, Mori C, Burckhardt S, Hider RC. Intravenous iron preparations transiently generate non-transferrin-bound iron from two proposed pathways. Haematologica. 2021 Nov 1;106(11):2885-2896.

12. Frise MC, Cheng HY, Nickol AH, Curtis MK, Pollard KA, Roberts DJ, Ratcliffe PJ, Dorrington KL, Robbins PA. Clinical iron deficiency disturbs normal human responses to hypoxia. J Clin Invest. 2016 Jun 1;126(6):2139-50.

13. Pai AB, Conner T, McQuade CR, Olp J, Hicks P. Non-transferrin bound iron, cytokine activation and intracellular reactive oxygen species generation in hemodialysis patients receiving intravenous iron dextran or iron sucrose. Biometals. 2011 Aug;24(4):603-13.

14. Noetzli LJ, Carson SM, Nord AS, Coates TD, Wood JC. Longitudinal analysis of heart and liver iron in thalassemia major. Blood. 2008;112:2973–2978.

15. Gulati V, Harikrishnan P, Palaniswamy C, Aronow WS, Jain D, Frishman WH. Cardiac involvement in hemochromatosis. Cardiology in Review. 2014;22:56–68.

16. Lagan J, Naish JH, Simpson K, Zi M, Cartwright EJ, Foden P, Morris J, Clark D, Birchall L, Caldwell J, Trafford A, Fortune C, Cullen M, Chaudhuri N, Fildes J, Sarma J, Schelbert EB, Schmitt M, Piper Hanley K, Miller CA. Substrate for the Myocardial Inflammation-Heart Failure Hypothesis Identified Using Novel USPIO Methodology. JACC Cardiovasc Imaging. 2021 Feb;14(2):365-376.

© Copyright: The University of Oxford and Oxford University Hospitals NHS Foundation Trust 2019

#### 22. APPENDIX A: STUDY FLOW CHART

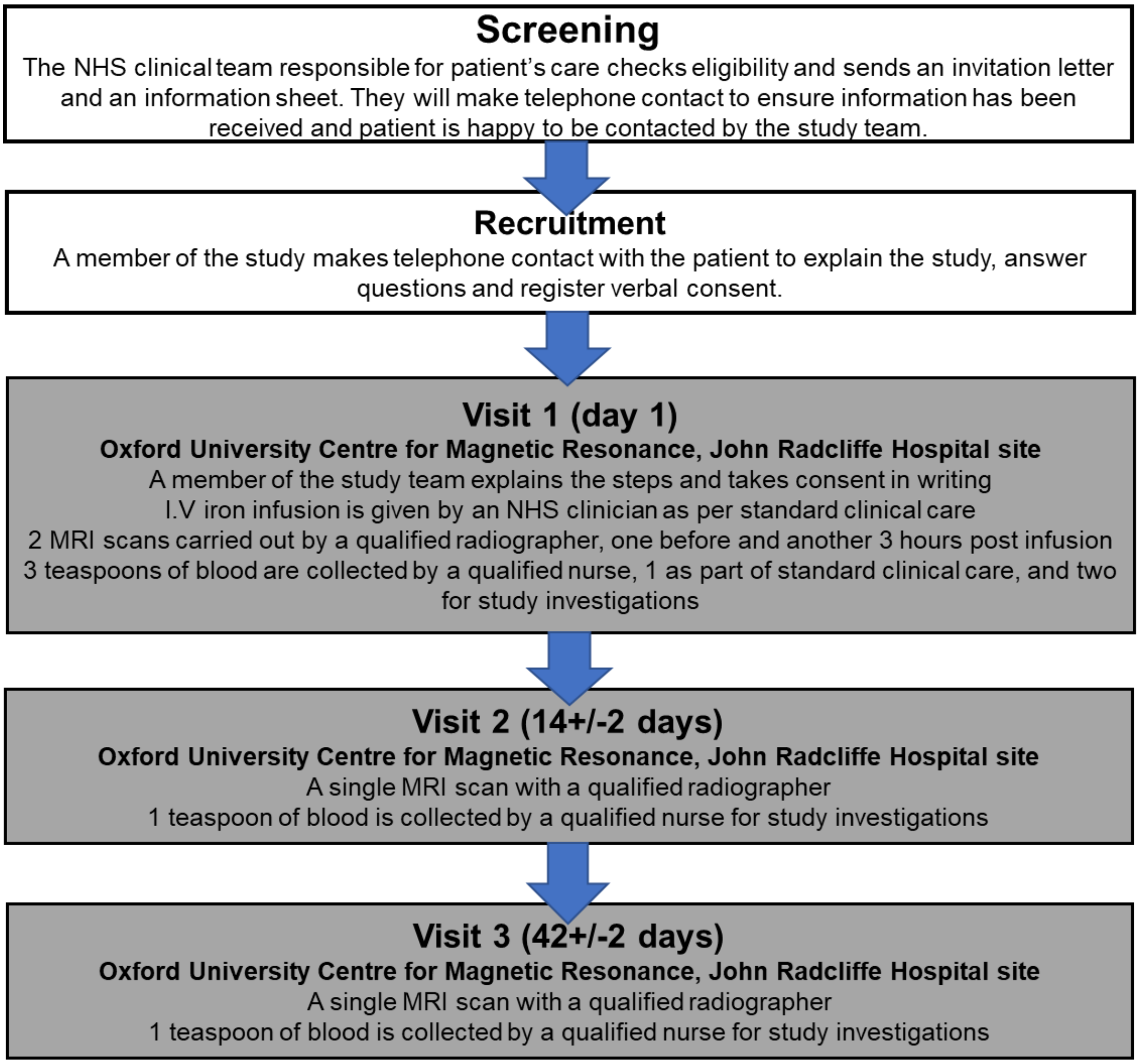

© Copyright: The University of Oxford and Oxford University Hospitals NHS Foundation Trust 2019

#### 23. APPENDIX B: SCHEDULE OF STUDY PROCEDURES

| **Procedures** | **Screening** | **Baseline (OCMR)** | **Timepoint 1 (OCMR)** | **Timepoint 2 (OCMR)** | **Time point 3 (OCMR)** |
|---|---|---|---|---|---|
| | | **Day 1 - prior to Ferinject infusion** | **Day 1- 3hours post Ferinject infusion** | **Day 14 (+/- 2days) post Ferinject infusion** | **Day 42 (+/- 2 days post Ferinject infusion** |
| Eligibility check | X | | | | |
| Informed consent | X (verbal) | X | | | |
| Demographics | | X | | | |
| Medical history | | X | | | |
| Research blood samples | | X | X | X | X |
| Clinical data | | X | | | |
| MRI scan | | X | X | X | X |

© Copyright: The University of Oxford and Oxford University Hospitals NHS Foundation Trust 2019

#### 24. APPENDIX C: AMENDMENT HISTORY

| Amendment No. | Protocol Version No. | Date issued | Author(s) of changes | Details of Changes made |
|---|---|---|---|---|

List details of all protocol amendments here whenever a new version of the protocol is produced.

Protocol amendments must be submitted to the Sponsor for approval prior to submission to the REC committee and HRA (where required).

#### 25. APPENDIX D: STUDY PLAN

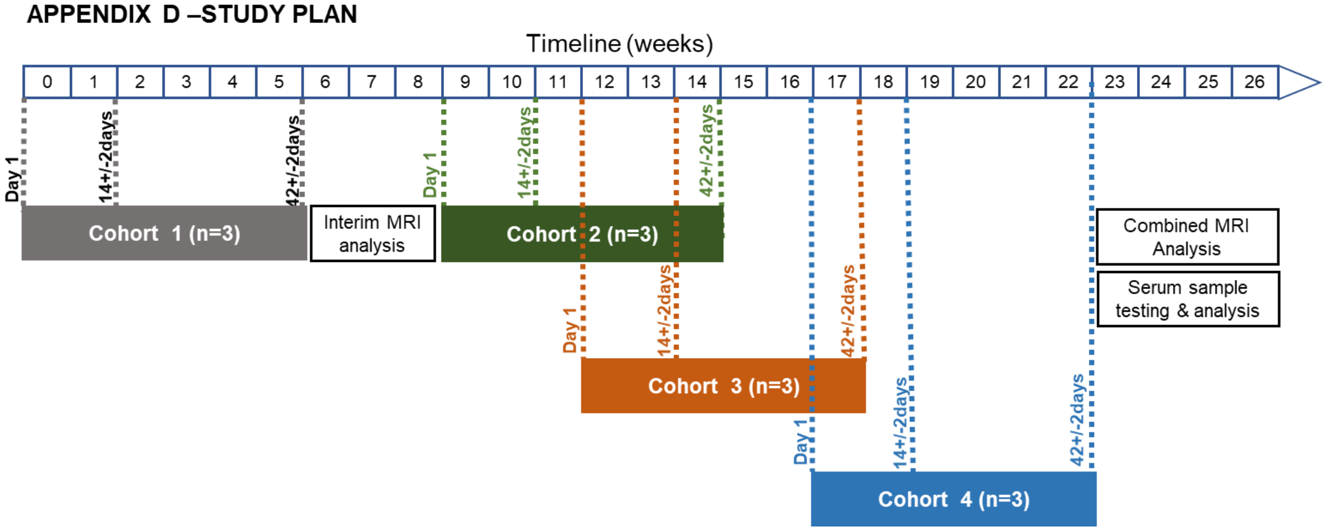

© Copyright: The University of Oxford and Oxford University Hospitals NHS Foundation Trust 2019

**Table of contents**

\\USER

Cardiac Research Protocols

IV IRON

IV IRON4

trufi_loc_multi_iPAT@c FOV500
trufi_loc_multi_iPAT@c FOV500
Trans HASTE- 10mm no gap non bh
Cor HASTE- 10mm no gap non bh
trufi_2-chamber_iPAT
trufi_4-chamber_iPAT
trufi_shortaxis_iPAT
HLA tf2d15_retro_iPAT3
VLA tf2d15_retro_iPAT3
LVOT tf2d15_retro_iPAT3
SA STACK tf2d15_retro_iPAT3
VLS_ShMOLLI_192i_d11_nFilt
KLS_ShMOLLI_192i_d11_nFilt_FOV460
VLST2Map_TrueFISP
VLSShMOLLI_C2P_m
VLST2StarMap_8echo_heart
VLSFB_MOCO_gt_T2star_DB_8e_128
FS_HiFE
VLSFB_MOCO_gt_T2star_DB_8e_160
FS_LowNFE
VLSnonBH_T2StarMap_12echo_liver
KLST2Map_TrueFISP
KLSShMOLLI_C2P_m
KLST2StarMap_8echo_heart
KLSFB_MOCO_gt_T2star_DB_8e_128
FS_HiFE
KLSFB_MOCO_gt_T2star_DB_8e_160
FS_LowNFE
KLSnonBH_T2StarMap_12echo_liver

| \\USER\Cardiac Research Protocols\IV IRON\IV IRON4\trufi_loc_multi_iPAT@c FOV500 |
|---|
| TA: 7.2 s PM: ISO Voxel size: 2.0×2.0×8.0 mmPAT: 2 Rel. SNR: 1.00 : tfi |

#### Properties

| | |
|---|---|
| Prio recon | Off |
| Load images to viewer | On |
| Inline movie | Off |
| Auto store images | On |
| Load images to stamp segments | On |
| Load images to graphic segments | On |
| Auto open inline display | Off |
| Auto close inline display | Off |
| Start measurement without further preparation | On |
| Wait for user to start | Off |
| Start measurements | Single measurement |

#### Routine

| | |
|---|---|
| Slice group | 1 |
| Slices | 3 |
| Dist. factor | 300 % |
| Position | L0.0 P0.0 H52.3 mm |
| Orientation | Transversal |
| Phase enc. dir. | A >> P |
| Slice group | 2 |
| Slices | 3 |
| Dist. factor | 300 % |
| Position | L30.0 P30.0 H52.3 mm |
| Orientation | Sagittal |
| Phase enc. dir. | A >> P |
| Slice group | 3 |
| Slices | 3 |
| Dist. factor | 300 % |
| Position | L0.0 P40.0 H52.3 mm |
| Orientation | Coronal |
| Phase enc. dir. | R >> L |
| AutoAlign | --- |
| Phase oversampling | 0 % |
| FoV read | 500 mm |
| FoV phase | 100.0 % |
| Slice thickness | 8.0 mm |
| TR | 288.36 ms |
| TE | 1.06 ms |
| Averages | 1 |
| Concatenations | 9 |
| Filter | Distortion Corr.(2D), Prescan Normalize |
| Coil elements | BO1-3;SP1-3 |

#### Contrast - Common

| | |
|---|---|
| TR | 288.36 ms |
| TE | 1.06 ms |
| TD | 0 ms |
| Magn. preparation | None |
| Flip angle | 80 deg |
| Fat suppr. | None |
| Wrap-up Magn. | Restore |

#### Contrast - Dynamic

| | |
|---|---|
| Averages | 1 |
| Averaging mode | Short term |
| Reconstruction | Magnitude |
| Measurements | 1 |
| Multiple series | Each measurement |

#### Resolution - Common

| | |
|---|---|
| FoV read | 500 mm |
| FoV phase | 100.0 % |
| Slice thickness | 8.0 mm |
| Base resolution | 256 |
| Phase resolution | 66 % |
| Phase partial Fourier | Off |
| Trajectory | Cartesian |
| Interpolation | Off |

#### Resolution - iPAT

| | |
|---|---|
| PAT mode | GRAPPA |
| Accel. factor PE | 2 |
| Ref. lines PE | 24 |
| Reference scan mode | Integrated |

#### Resolution - Filter Image

| | |
|---|---|
| Image Filter | Off |
| Distortion Corr. | On |
| Mode | 2D |
| Unfiltered images | Off |
| Prescan Normalize | On |
| Unfiltered images | Off |
| Normalize | Off |
| B1 filter | Off |

#### Resolution - Filter Rawdata

| | |
|---|---|
| Raw filter | Off |
| Elliptical filter | Off |
| POCS | Off |

#### Geometry - Common

| | |
|---|---|
| Slice group | 1 |
| Slices | 3 |
| Dist. factor | 300 % |
| Position | L0.0 P0.0 H52.3 mm |
| Orientation | Transversal |
| Phase enc. dir. | A >> P |
| Slice group | 2 |
| Slices | 3 |
| Dist. factor | 300 % |
| Position | L30.0 P30.0 H52.3 mm |
| Orientation | Sagittal |
| Phase enc. dir. | A >> P |
| Slice group | 3 |
| Slices | 3 |
| Dist. factor | 300 % |
| Position | L0.0 P40.0 H52.3 mm |
| Orientation | Coronal |
| Phase enc. dir. | R >> L |
| FoV read | 500 mm |
| FoV phase | 100.0 % |
| Slice thickness | 8.0 mm |
| TR | 288.36 ms |
| Multi-slice mode | Sequential |
| Series | Descending |
| Concatenations | 9 |

#### Geometry - AutoAlign

| | |
|---|---|
| Slice group | 1 |
| Position | L0.0 P0.0 H52.3 mm |

#### Geometry - AutoAlign

| | |
|---|---|
| Orientation | Transversal |
| Phase enc. dir. | A >> P |
| Slice group | 2 |
| Position | L30.0 P30.0 H52.3 mm |
| Orientation | Sagittal |
| Phase enc. dir. | A >> P |
| Slice group | 3 |
| Position | L0.0 P40.0 H52.3 mm |
| Orientation | Coronal |
| Phase enc. dir. | R >> L |
| AutoAlign | --- |
| Initial Position | L0.0 P0.0 H52.3 |
| Phase | 0.0 mm |
| Read | 0.0 mm |
| Shift | 52.3 mm |
| Initial Rotation | 0.00 deg |
| Initial Orientation | Transversal |

#### Geometry - Saturation

| | |
|---|---|
| Fat suppr. | None |
| Wrap-up Magn. | Restore |
| Special sat. | None |

#### Geometry - Navigator

#### System - Miscellaneous

| | |
|---|---|
| Positioning mode | ISO |
| Table position | H |
| Table position | 52 mm |
| MSMA | S - C - T |
| Sagittal | R >> L |
| Coronal | A >> P |
| Transversal | F >> H |
| Coil Combine Mode | Adaptive Combine |
| Save uncombined | Off |
| Matrix Optimization | Off |
| Coil Focus | Flat |
| AutoAlign | --- |
| Coil Select Mode | Default |

#### System - Adjustments

| | |
|---|---|
| B0 Shim mode | Tune up |
| Adjust with body coil | Off |
| Confirm freq. adjustment | Off |
| Assume Dominant Fat | Off |
| Assume Silicone | Off |
| Adjustment Tolerance | Auto |

#### System - Adjust Volume

| | |
|---|---|
| Position | Isocenter |
| Orientation | Transversal |
| Rotation | 0.00 deg |
| A >> P | 263 mm |
| R >> L | 350 mm |
| F >> H | 350 mm |
| Reset | Off |

#### System - Tx/Rx

| | |
|---|---|
| Frequency 1H | 63.678323 MHz |
| Correction factor | 1 |
| Gain | High |
| Img. Scale Cor. | 1.000 |
| Reset | Off |
| ? Ref. amplitude 1H | 0.000 V |

#### Physio - Signal1

| | |
|---|---|
| 1st Signal/Mode | ECG/Trigger |
| Average cycle | No Signal ms |
| Average cycle | No Signal ms |
| Captured cycle | -not set- |
| Acquisition window | 800 ms |
| Trigger pulse | 1 |
| Trigger delay | 400 ms |
| TR | 288.36 ms |
| Concatenations | 9 |
| Segments | 96 |
| Phases | 1 |
| Adaptive Triggering | Off |

#### Physio - Cardiac

| | |
|---|---|
| Tagging | None |
| Magn. preparation | None |
| Fat suppr. | None |
| Dark blood | Off |
| FoV read | 500 mm |
| FoV phase | 100.0 % |
| Phase resolution | 66 % |
| Cine | Off |
| Trajectory | Cartesian |
| Dummy heartbeats | 0 |

#### Physio - PACE

| | |
|---|---|
| Resp. control | Off |
| Concatenations | 9 |

#### Inline - Common

| | |
|---|---|
| Subtract | Off |
| Measurements | 1 |
| StdDev | Off |
| Save original images | On |

#### Inline - Cardiac

| | |
|---|---|
| Inline Evaluation | Off |
| Magn. preparation | None |
| Contrasts | 1 |
| TE | 1.06 ms |
| TR | 288.36 ms |
| Save original images | On |

#### Inline - MIP

| | |
|---|---|
| MIP-Sag | Off |
| MIP-Cor | Off |
| MIP-Tra | Off |
| MIP-Time | Off |
| Save original images | On |

#### Inline - Composing

| | |
|---|---|
| Inline Composing | Off |
| Distortion Corr. | On |
| Mode | 2D |
| Unfiltered images | Off |

#### Sequence - Part 1

| | |
|---|---|
| Introduction | Off |
| Dimension | 2D |
| Reordering | Linear |
| Asymmetric echo | Weak |
| Contrasts | 1 |
| Optimization | Min. TE |

##### Sequence - Part 1

| | |
|---|---|
| Multi-slice mode | Sequential |
| Echo spacing | 2.5 ms |
| Sequence type | Trufi |
| Bandwidth | 1149 Hz/Px |

##### Sequence - Part 2

| | |
|---|---|
| Define | Shots |
| Shots per slice | 1 |
| Segments | 96 |
| Trufi delta freq. | 0 Hz |
| RF pulse type | Fast |
| Gradient mode | Fast |
| Excitation | Slice-sel. |
| Flip angle mode | Constant |
| Cine | Off |

##### Sequence - Assistant

| | |
|---|---|
| Mode | Min flip angle |
| Min flip angle | 50 deg |
| Allowed delay | 0 s |

| \\USER\Cardiac Research Protocols\IV IRON\IV IRON4\trufi_loc_multi_iPAT@c FOV500 |
|---|
| TA: 7.2 s PM: ISO Voxel size: 2.0×2.0×8.0 mmPAT: 2 Rel. SNR: 1.00 : tfi |

#### Properties

| | |
|---|---|
| Prio recon | Off |
| Load images to viewer | On |
| Inline movie | Off |
| Auto store images | On |
| Load images to stamp segments | On |
| Load images to graphic segments | On |
| Auto open inline display | Off |
| Auto close inline display | Off |
| Start measurement without further preparation | Off |
| Wait for user to start | Off |
| Start measurements | Single measurement |

#### Routine

| | |
|---|---|
| Slice group | 1 |
| Slices | 3 |
| Dist. factor | 300 % |
| Position | L0.0 A30.0 H0.0 mm |
| Orientation | Transversal |
| Phase enc. dir. | A >> P |
| Slice group | 2 |
| Slices | 3 |
| Dist. factor | 300 % |
| Position | L30.0 P0.0 H0.0 mm |
| Orientation | Sagittal |
| Phase enc. dir. | A >> P |
| Slice group | 3 |
| Slices | 3 |
| Dist. factor | 300 % |
| Position | L0.0 P10.0 H0.0 mm |
| Orientation | Coronal |
| Phase enc. dir. | R >> L |
| AutoAlign | --- |
| Phase oversampling | 0 % |
| FoV read | 500 mm |
| FoV phase | 100.0 % |
| Slice thickness | 8.0 mm |
| TR | 288.36 ms |
| TE | 1.06 ms |
| Averages | 1 |
| Concatenations | 9 |
| Filter | Distortion Corr.(2D), Prescan Normalize |
| Coil elements | BO1-3;SP1-3 |

#### Contrast - Common

| | |
|---|---|
| TR | 288.36 ms |
| TE | 1.06 ms |
| TD | 0 ms |
| Magn. preparation | None |
| Flip angle | 80 deg |
| Fat suppr. | None |
| Wrap-up Magn. | Restore |

#### Contrast - Dynamic

| | |
|---|---|
| Averages | 1 |
| Averaging mode | Short term |
| Reconstruction | Magnitude |
| Measurements | 1 |
| Multiple series | Each measurement |

#### Resolution - Common

| | |
|---|---|
| FoV read | 500 mm |
| FoV phase | 100.0 % |
| Slice thickness | 8.0 mm |
| Base resolution | 256 |
| Phase resolution | 66 % |
| Phase partial Fourier | Off |
| Trajectory | Cartesian |
| Interpolation | Off |

#### Resolution - iPAT

| | |
|---|---|
| PAT mode | GRAPPA |
| Accel. factor PE | 2 |
| Ref. lines PE | 24 |
| Reference scan mode | Integrated |

#### Resolution - Filter Image

| | |
|---|---|
| Image Filter | Off |
| Distortion Corr. | On |
| Mode | 2D |
| Unfiltered images | Off |
| Prescan Normalize | On |
| Unfiltered images | Off |
| Normalize | Off |
| B1 filter | Off |

#### Resolution - Filter Rawdata

| | |
|---|---|
| Raw filter | Off |
| Elliptical filter | Off |
| POCS | Off |

#### Geometry - Common

| | |
|---|---|
| Slice group | 1 |
| Slices | 3 |
| Dist. factor | 300 % |
| Position | L0.0 A30.0 H0.0 mm |
| Orientation | Transversal |
| Phase enc. dir. | A >> P |
| Slice group | 2 |
| Slices | 3 |
| Dist. factor | 300 % |
| Position | L30.0 P0.0 H0.0 mm |
| Orientation | Sagittal |
| Phase enc. dir. | A >> P |
| Slice group | 3 |
| Slices | 3 |
| Dist. factor | 300 % |
| Position | L0.0 P10.0 H0.0 mm |
| Orientation | Coronal |
| Phase enc. dir. | R >> L |
| FoV read | 500 mm |
| FoV phase | 100.0 % |
| Slice thickness | 8.0 mm |
| TR | 288.36 ms |
| Multi-slice mode | Sequential |
| Series | Descending |
| Concatenations | 9 |

#### Geometry - AutoAlign

| | |
|---|---|
| Slice group | 1 |
| Position | L0.0 A30.0 H0.0 mm |

#### Geometry - AutoAlign

| | |
|---|---|
| Orientation | Transversal |
| Phase enc. dir. | A >> P |
| Slice group | 2 |
| Position | L30.0 P0.0 H0.0 mm |
| Orientation | Sagittal |
| Phase enc. dir. | A >> P |
| Slice group | 3 |
| Position | L0.0 P10.0 H0.0 mm |
| Orientation | Coronal |
| Phase enc. dir. | R >> L |
| AutoAlign | --- |
| Initial Position | L0.0 A30.0 H0.0 |
| Phase | -30.0 mm |
| Read | 0.0 mm |
| Shift | 0.0 mm |
| Initial Rotation | 0.00 deg |
| Initial Orientation | Transversal |

#### Geometry - Saturation

| | |
|---|---|
| Fat suppr. | None |
| Wrap-up Magn. | Restore |
| Special sat. | None |

#### Geometry - Navigator

#### System - Miscellaneous

| | |
|---|---|
| Positioning mode | ISO |
| Table position | H |
| Table position | 0 mm |
| MSMA | S - C - T |
| Sagittal | R >> L |
| Coronal | A >> P |
| Transversal | F >> H |
| Coil Combine Mode | Adaptive Combine |
| Save uncombined | Off |
| Matrix Optimization | Off |
| Coil Focus | Flat |
| AutoAlign | --- |
| Coil Select Mode | Default |

#### System - Adjustments

| | |
|---|---|
| B0 Shim mode | Tune up |
| Adjust with body coil | Off |
| Confirm freq. adjustment | Off |
| Assume Dominant Fat | Off |
| Assume Silicone | Off |
| Adjustment Tolerance | Auto |

#### System - Adjust Volume

| | |
|---|---|
| Position | Isocenter |
| Orientation | Transversal |
| Rotation | 0.00 deg |
| A >> P | 263 mm |
| R >> L | 350 mm |
| F >> H | 350 mm |
| Reset | Off |

#### System - Tx/Rx

| | |
|---|---|
| Frequency 1H | 63.678323 MHz |
| Correction factor | 1 |
| Gain | High |
| Img. Scale Cor. | 1.000 |
| Reset | Off |
| ? Ref. amplitude 1H | 0.000 V |

#### Physio - Signal1

| | |
|---|---|
| 1st Signal/Mode | ECG/Trigger |
| Average cycle | No Signal ms |
| Average cycle | No Signal ms |
| Captured cycle | -not set- |
| Acquisition window | 800 ms |
| Trigger pulse | 1 |
| Trigger delay | 400 ms |
| TR | 288.36 ms |
| Concatenations | 9 |
| Segments | 96 |
| Phases | 1 |
| Adaptive Triggering | Off |

#### Physio - Cardiac

| | |
|---|---|
| Tagging | None |
| Magn. preparation | None |
| Fat suppr. | None |
| Dark blood | Off |
| FoV read | 500 mm |
| FoV phase | 100.0 % |
| Phase resolution | 66 % |
| Cine | Off |
| Trajectory | Cartesian |
| Dummy heartbeats | 0 |

#### Physio - PACE

| | |
|---|---|
| Resp. control | Off |
| Concatenations | 9 |

#### Inline - Common

| | |
|---|---|
| Subtract | Off |
| Measurements | 1 |
| StdDev | Off |
| Save original images | On |

#### Inline - Cardiac

| | |
|---|---|
| Inline Evaluation | Off |
| Magn. preparation | None |
| Contrasts | 1 |
| TE | 1.06 ms |
| TR | 288.36 ms |
| Save original images | On |

#### Inline - MIP

| | |
|---|---|
| MIP-Sag | Off |
| MIP-Cor | Off |
| MIP-Tra | Off |
| MIP-Time | Off |
| Save original images | On |

#### Inline - Composing

| | |
|---|---|
| Inline Composing | Off |
| Distortion Corr. | On |
| Mode | 2D |
| Unfiltered images | Off |

#### Sequence - Part 1

| | |
|---|---|
| Introduction | Off |
| Dimension | 2D |
| Reordering | Linear |
| Asymmetric echo | Weak |
| Contrasts | 1 |
| Optimization | Min. TE |

##### Sequence - Part 1

| | |
|---|---|
| Multi-slice mode | Sequential |
| Echo spacing | 2.5 ms |
| Sequence type | Trufi |
| Bandwidth | 1149 Hz/Px |

##### Sequence - Part 2

| | |
|---|---|
| Define | Shots |
| Shots per slice | 1 |
| Segments | 96 |
| Trufi delta freq. | 0 Hz |
| RF pulse type | Fast |
| Gradient mode | Fast |
| Excitation | Slice-sel. |
| Flip angle mode | Constant |
| Cine | Off |

##### Sequence - Assistant

| | |
|---|---|
| Mode | Min flip angle |
| Min flip angle | 50 deg |
| Allowed delay | 0 s |

| \\USER\Cardiac Research Protocols\IV IRON\IV IRON4\Trans HASTE- 10mm no gap non bh |
|---|
| TA: 0:56 PM: REF Voxel size: 1.4×1.4×10.0 mmPAT: Off Rel. SNR: 1.00 : h |

#### Properties

| | |
|---|---|
| Prio recon | Off |
| Load images to viewer | On |
| Inline movie | Off |
| Auto store images | On |
| Load images to stamp segments | On |
| Load images to graphic segments | On |
| Auto open inline display | Off |
| Auto close inline display | Off |
| Start measurement without further preparation | Off |
| Wait for user to start | On |
| Start measurements | Single measurement |

#### Routine

| | |
|---|---|
| Slice group | 1 |
| Slices | 35 |
| Dist. factor | 0 % |
| Position | L0.0 A49.4 H32.4 mm |
| Orientation | Transversal |
| Phase enc. dir. | A >> P |
| AutoAlign | --- |
| Phase oversampling | 0 % |
| FoV read | 360 mm |
| FoV phase | 78.1 % |
| Slice thickness | 10.0 mm |
| TR | 800.0 ms |
| TE | 26 ms |
| Averages | 1 |
| Concatenations | 2 |
| Filter | Distortion Corr.(2D), Prescan Normalize, Elliptical filter, Image Filter |
| Coil elements | BO1-3;SP1-4 |

#### Contrast - Common

| | |
|---|---|
| TR | 800.0 ms |
| TE | 26 ms |
| TD | 0.0 ms |
| MTC | Off |
| Magn. preparation | None |
| Flip angle | 160 deg |
| Fat suppr. | None |
| Water suppr. | None |
| Restore magn. | Off |

#### Contrast - Dynamic

| | |
|---|---|
| Averages | 1 |
| Averaging mode | Long term |
| Reconstruction | Magnitude |
| Measurements | 1 |
| Multiple series | Off |

#### Resolution - Common

| | |
|---|---|
| FoV read | 360 mm |
| FoV phase | 78.1 % |
| Slice thickness | 10.0 mm |
| Base resolution | 256 |
| Phase resolution | 59 % |
| Phase partial Fourier | 5/8 |
| Interpolation | Off |

#### Resolution - iPAT

| | |
|---|---|
| PAT mode | None |

#### Resolution - Filter Image

| | |
|---|---|
| Image Filter | On |
| ! Intensity | Medium |
| Edge Enhancement | 1 |
| Smoothing | 1 |
| Unfiltered images | Off |
| Distortion Corr. | On |
| Mode | 2D |
| Unfiltered images | Off |
| Prescan Normalize | On |
| Unfiltered images | Off |
| Normalize | Off |
| B1 filter | Off |

#### Resolution - Filter Rawdata

| | |
|---|---|
| Raw filter | Off |
| Elliptical filter | On |

#### Geometry - Common

| | |
|---|---|
| Slice group | 1 |
| Slices | 35 |
| Dist. factor | 0 % |
| Position | L0.0 A49.4 H32.4 mm |
| Orientation | Transversal |
| Phase enc. dir. | A >> P |
| FoV read | 360 mm |
| FoV phase | 78.1 % |
| Slice thickness | 10.0 mm |
| TR | 800.0 ms |
| Multi-slice mode | Single shot |
| Series | Interleaved |
| Concatenations | 2 |

#### Geometry - AutoAlign

| | |
|---|---|
| Slice group | 1 |
| Position | L0.0 A49.4 H32.4 mm |
| Orientation | Transversal |
| Phase enc. dir. | A >> P |
| AutoAlign | --- |
| Initial Position | L0.0 A49.4 H18.4 |
| Phase | -49.4 mm |
| Read | 0.0 mm |
| Shift | 18.4 mm |
| Initial Rotation | 0.00 deg |
| Initial Orientation | Transversal |

#### Geometry - Saturation

| | |
|---|---|
| Fat suppr. | None |
| Water suppr. | None |
| Restore magn. | Off |
| Special sat. | None |

#### Geometry - Navigator

#### System - Miscellaneous

| | |
|---|---|
| Positioning mode | REF |
| Table position | H |
| Table position | 14 mm |

#### System - Miscellaneous

| | |
|---|---|
| MSMA | S - C - T |
| Sagittal | R >> L |
| Coronal | A >> P |
| Transversal | F >> H |
| Coil Combine Mode | Adaptive Combine |
| Save uncombined | Off |
| Matrix Optimization | Off |
| Coil Focus | Flat |
| AutoAlign | --- |
| Coil Select Mode | Default |

#### System - Adjustments

| | |
|---|---|
| B0 Shim mode | Tune up |
| Adjust with body coil | On |
| Confirm freq. adjustment | Off |
| Assume Dominant Fat | Off |
| Assume Silicone | Off |
| Adjustment Tolerance | Auto |

#### System - Adjust Volume

| | |
|---|---|
| Position | Isocenter |
| Orientation | Transversal |
| Rotation | 0.00 deg |
| A >> P | 263 mm |
| R >> L | 350 mm |
| F >> H | 350 mm |
| Reset | Off |

#### System - Tx/Rx

| | |
|---|---|
| Frequency 1H | 63.678323 MHz |
| Correction factor | 1 |
| Gain | High |
| Img. Scale Cor. | 1.000 |
| Reset | Off |
| ? Ref. amplitude 1H | 0.000 V |

#### Physio - Signal1

| | |
|---|---|
| 1st Signal/Mode | ECG/Trigger |
| Average cycle | No Signal ms |
| Average cycle | No Signal ms |
| Captured cycle | -not set- |
| Acquisition window | 800 ms |
| Trigger pulse | 2 |
| Trigger delay | 0 ms |
| TR | 800.0 ms |
| Concatenations | 2 |
| Phases | 1 |

#### Physio - Cardiac

| | |
|---|---|
| Magn. preparation | None |
| Fat suppr. | None |
| Dark blood | On |
| Dark blood thickness | 200 % |
| FoV read | 360 mm |
| FoV phase | 78.1 % |
| Phase resolution | 59 % |

#### Physio - PACE

| | |
|---|---|
| Resp. control | Off |
| Concatenations | 2 |

#### Inline - Common

| | |
|---|---|
| Subtract | Off |
| Measurements | 1 |
| StdDev | Off |
| Save original images | On |

#### Inline - MIP

| | |
|---|---|
| MIP-Sag | Off |
| MIP-Cor | Off |
| MIP-Tra | Off |
| MIP-Time | Off |
| Save original images | On |

#### Inline - Composing

| | |
|---|---|
| Inline Composing | Off |
| Distortion Corr. | On |
| Mode | 2D |
| Unfiltered images | Off |

#### Sequence - Part 1

| | |
|---|---|
| Introduction | Off |
| Dimension | 2D |
| Contrasts | 1 |
| Flow comp. | No |
| Multi-slice mode | Single shot |
| Echo spacing | 2.92 ms |
| Bandwidth | 781 Hz/Px |

#### Sequence - Part 2

| | |
|---|---|
| RF pulse type | Fast |
| Gradient mode | Fast |
| Turbo factor | 118 |

#### Sequence - Assistant

| | |
|---|---|
| Mode | Off |
| Allowed delay | 30 s |

| \\USER\Cardiac Research Protocols\IV IRON\IV IRON4\Cor HASTE- 10mm no gap non bh |
|---|
| TA: 0:27 PM: REF Voxel size: 1.8×1.8×10.0 mmPAT: Off Rel. SNR: 1.00 : h |

#### Properties

| | |
|---|---|
| Prio recon | Off |
| Load images to viewer | On |
| Inline movie | Off |
| Auto store images | On |
| Load images to stamp segments | On |
| Load images to graphic segments | On |
| Auto open inline display | Off |
| Auto close inline display | Off |
| Start measurement without further preparation | Off |
| Wait for user to start | On |
| Start measurements | Single measurement |

#### Routine

| | |
|---|---|
| Slice group | 1 |
| Slices | 17 |
| Dist. factor | 0 % |
| Position | L4.7 P16.3 F10.4 mm |
| Orientation | Coronal |
| Phase enc. dir. | R >> L |
| AutoAlign | --- |
| Phase oversampling | 0 % |
| FoV read | 460 mm |
| FoV phase | 78.1 % |
| Slice thickness | 10.0 mm |
| TR | 800.0 ms |
| TE | 26 ms |
| Averages | 1 |
| Concatenations | 2 |
| Filter | Distortion Corr.(2D), Prescan Normalize, Elliptical filter, Image Filter |
| Coil elements | B1-5;SP3-6 |

#### Contrast - Common

| | |
|---|---|
| TR | 800.0 ms |
| TE | 26 ms |
| TD | 0.0 ms |
| MTC | Off |
| Magn. preparation | None |
| Flip angle | 160 deg |
| Fat suppr. | None |
| Water suppr. | None |
| Restore magn. | Off |

#### Contrast - Dynamic

| | |
|---|---|
| Averages | 1 |
| Averaging mode | Long term |
| Reconstruction | Magnitude |
| Measurements | 1 |
| Multiple series | Off |

#### Resolution - Common

| | |
|---|---|
| FoV read | 460 mm |
| FoV phase | 78.1 % |
| Slice thickness | 10.0 mm |
| Base resolution | 256 |
| Phase resolution | 59 % |
| Phase partial Fourier | 5/8 |
| Interpolation | Off |

#### Resolution - iPAT

| | |
|---|---|
| PAT mode | None |

#### Resolution - Filter Image

| | |
|---|---|
| Image Filter | On |
| ! Intensity | Medium |
| Edge Enhancement | 1 |
| Smoothing | 1 |
| Unfiltered images | Off |
| Distortion Corr. | On |
| Mode | 2D |
| Unfiltered images | Off |
| Prescan Normalize | On |
| Unfiltered images | Off |
| Normalize | Off |
| B1 filter | Off |

#### Resolution - Filter Rawdata

| | |
|---|---|
| Raw filter | Off |
| Elliptical filter | On |

#### Geometry - Common

| | |
|---|---|
| Slice group | 1 |
| Slices | 17 |
| Dist. factor | 0 % |
| Position | L4.7 P16.3 F10.4 mm |
| Orientation | Coronal |
| Phase enc. dir. | R >> L |
| FoV read | 460 mm |
| FoV phase | 78.1 % |
| Slice thickness | 10.0 mm |
| TR | 800.0 ms |
| Multi-slice mode | Single shot |
| Series | Interleaved |
| Concatenations | 2 |

#### Geometry - AutoAlign

| | |
|---|---|
| Slice group | 1 |
| Position | L4.7 P16.3 F10.4 mm |
| Orientation | Coronal |
| Phase enc. dir. | R >> L |
| AutoAlign | --- |
| Initial Position | L4.7 P16.3 F24.4 |
| Phase | 4.7 mm |
| Read | 24.4 mm |
| Shift | 16.3 mm |
| Initial Rotation | 0.00 deg |
| Initial Orientation | Coronal |

#### Geometry - Saturation

| | |
|---|---|
| Fat suppr. | None |
| Water suppr. | None |
| Restore magn. | Off |
| Special sat. | None |

#### Geometry - Navigator

#### System - Miscellaneous

| | |
|---|---|
| Positioning mode | REF |
| Table position | H |
| Table position | 14 mm |

#### System - Miscellaneous

| | |
|---|---|
| MSMA | S - C - T |
| Sagittal | R >> L |
| Coronal | A >> P |
| Transversal | F >> H |
| Coil Combine Mode | Adaptive Combine |
| Save uncombined | Off |
| Matrix Optimization | Off |
| Coil Focus | Flat |
| AutoAlign | --- |
| Coil Select Mode | Default |

#### System - Adjustments

| | |
|---|---|
| B0 Shim mode | Tune up |
| Adjust with body coil | On |
| Confirm freq. adjustment | Off |
| Assume Dominant Fat | Off |
| Assume Silicone | Off |
| Adjustment Tolerance | Auto |

#### System - Adjust Volume

| | |
|---|---|
| ! Position | L7.9 P14.3 F13.0 mm |
| ! Orientation | Transversal |
| ! Rotation | -0.63 deg |
| ! A >> P | 176 mm |
| ! R >> L | 350 mm |
| ! F >> H | 457 mm |
| Reset | Off |

#### System - Tx/Rx

| | |
|---|---|
| Frequency 1H | 63.678323 MHz |
| Correction factor | 1 |
| Gain | High |
| Img. Scale Cor. | 1.000 |
| Reset | Off |
| ? Ref. amplitude 1H | 0.000 V |

#### Physio - Signal1

| | |
|---|---|
| 1st Signal/Mode | ECG/Trigger |
| Average cycle | No Signal ms |
| Average cycle | No Signal ms |
| Captured cycle | -not set- |
| Acquisition window | 800 ms |
| Trigger pulse | 2 |
| Trigger delay | 0 ms |
| TR | 800.0 ms |
| Concatenations | 2 |
| Phases | 1 |

#### Physio - Cardiac

| | |
|---|---|
| Magn. preparation | None |
| Fat suppr. | None |
| Dark blood | On |
| Dark blood thickness | 200 % |
| FoV read | 460 mm |
| FoV phase | 78.1 % |
| Phase resolution | 59 % |

#### Physio - PACE

| | |
|---|---|
| Resp. control | Off |
| Concatenations | 2 |

#### Inline - Common

| | |
|---|---|
| Subtract | Off |

#### Inline - Common

| | |
|---|---|
| Measurements | 1 |
| StdDev | Off |
| Save original images | On |

#### Inline - MIP

| | |
|---|---|
| MIP-Sag | Off |
| MIP-Cor | Off |
| MIP-Tra | Off |
| MIP-Time | Off |
| Save original images | On |

#### Inline - Composing

| | |
|---|---|
| Inline Composing | Off |
| Distortion Corr. | On |
| Mode | 2D |
| Unfiltered images | Off |

#### Sequence - Part 1

| | |
|---|---|
| Introduction | Off |
| Dimension | 2D |
| Contrasts | 1 |
| Flow comp. | No |
| Multi-slice mode | Single shot |
| Echo spacing | 2.92 ms |
| Bandwidth | 781 Hz/Px |

#### Sequence - Part 2

| | |
|---|---|
| RF pulse type | Fast |
| Gradient mode | Fast |
| Turbo factor | 118 |

#### Sequence - Assistant

| | |
|---|---|
| Mode | Off |
| Allowed delay | 30 s |

**\\USER\Cardiac Research Protocols\IV IRON\IV IRON4\trufi_2-chamber_iPAT**

TA: 0.8 s PM: REF Voxel size: 1.5×1.5×8.0 mmPAT: 2 Rel. SNR: 1.00 : tfi

#### Properties

| | |
|---|---|
| Prio recon | Off |
| Load images to viewer | On |
| Inline movie | Off |
| Auto store images | On |
| Load images to stamp segments | Off |
| Load images to graphic segments | On |
| Auto open inline display | Off |
| Auto close inline display | Off |
| Start measurement without further preparation | Off |
| Wait for user to start | Off |
| Start measurements | Single measurement |

#### Routine

| | |
|---|---|
| Slice group | 1 |
| Slices | 1 |
| Dist. factor | 20 % |
| Position | R4.2 P39.2 H108.8 mm |
| Orientation | T > S12.4 > C-6.2 |
| Phase enc. dir. | A >> P |
| AutoAlign | --- |
| Phase oversampling | 0 % |
| FoV read | 380 mm |
| FoV phase | 87.5 % |
| Slice thickness | 8.0 mm |
| TR | 260.22 ms |
| TE | 1.16 ms |
| Averages | 1 |
| Concatenations | 1 |
| Filter | Distortion Corr.(2D), Prescan Normalize |
| Coil elements | B1,2;SP3,4 |

#### Contrast - Common

| | |
|---|---|
| TR | 260.22 ms |
| TE | 1.16 ms |
| Magn. preparation | None |
| Flip angle | 80 deg |
| Fat suppr. | None |
| Wrap-up Magn. | Restore |

#### Contrast - Dynamic

| | |
|---|---|
| Averages | 1 |
| Averaging mode | Short term |
| Reconstruction | Magnitude |
| Measurements | 1 |
| Multiple series | Each measurement |

#### Resolution - Common

| | |
|---|---|
| FoV read | 380 mm |
| FoV phase | 87.5 % |
| Slice thickness | 8.0 mm |
| Base resolution | 256 |
| Phase resolution | 64 % |
| Phase partial Fourier | Off |
| Trajectory | Cartesian |
| Interpolation | Off |

#### Resolution - iPAT

| | |
|---|---|
| PAT mode | GRAPPA |
| Accel. factor PE | 2 |
| Ref. lines PE | 24 |
| Reference scan mode | Integrated |

#### Resolution - Filter Image

| | |
|---|---|
| Image Filter | Off |
| Distortion Corr. | On |
| Mode | 2D |
| Unfiltered images | Off |
| Prescan Normalize | On |
| Unfiltered images | Off |
| Normalize | Off |
| B1 filter | Off |

#### Resolution - Filter Rawdata

| | |
|---|---|
| Raw filter | Off |
| Elliptical filter | Off |
| POCS | Off |

#### Geometry - Common

| | |
|---|---|
| Slice group | 1 |
| Slices | 1 |
| Dist. factor | 20 % |
| Position | R4.2 P39.2 H108.8 mm |
| Orientation | T > S12.4 > C-6.2 |
| Phase enc. dir. | A >> P |
| FoV read | 380 mm |
| FoV phase | 87.5 % |
| Slice thickness | 8.0 mm |
| TR | 260.22 ms |
| Multi-slice mode | Sequential |
| Series | Interleaved |
| Concatenations | 1 |

#### Geometry - AutoAlign

| | |
|---|---|
| Slice group | 1 |
| Position | R4.2 P39.2 H108.8 mm |
| Orientation | T > S12.4 > C-6.2 |
| Phase enc. dir. | A >> P |
| AutoAlign | --- |
| Initial Position | R4.2 P39.2 H56.8 |
| Phase | 32.9 mm |
| Read | -8.1 mm |
| Shift | 60.3 mm |
| Initial Rotation | 1.35 deg |
| Initial Orientation | T > S |
| T > S | 12.4 |
| > C | -6.2 |

#### Geometry - Saturation

| | |
|---|---|
| Fat suppr. | None |
| Wrap-up Magn. | Restore |
| Special sat. | None |

#### Geometry - Navigator

#### System - Miscellaneous

| | |
|---|---|
| Positioning mode | REF |
| Table position | H |
| Table position | 52 mm |

#### System - Miscellaneous

| | |
|---|---|
| MSMA | S - C - T |
| Sagittal | R >> L |
| Coronal | A >> P |
| Transversal | F >> H |
| Coil Combine Mode | Adaptive Combine |
| Save uncombined | Off |
| Matrix Optimization | Off |
| Coil Focus | Flat |
| AutoAlign | --- |
| Coil Select Mode | Default |

#### System - Adjustments

| | |
|---|---|
| B0 Shim mode | Tune up |
| Adjust with body coil | Off |
| Confirm freq. adjustment | Off |
| Assume Dominant Fat | Off |
| Assume Silicone | Off |
| Adjustment Tolerance | Auto |

#### System - Adjust Volume

| | |
|---|---|
| Position | Isocenter |
| Orientation | Transversal |
| Rotation | 0.00 deg |
| A >> P | 263 mm |
| R >> L | 350 mm |
| F >> H | 350 mm |
| Reset | Off |

#### System - Tx/Rx

| | |
|---|---|
| Frequency 1H | 63.678323 MHz |
| Correction factor | 1 |
| Gain | High |
| Img. Scale Cor. | 1.000 |
| Reset | Off |
| ? Ref. amplitude 1H | 0.000 V |

#### Physio - Signal1

| | |
|---|---|
| 1st Signal/Mode | ECG/Trigger |
| Average cycle | No Signal ms |
| Average cycle | No Signal ms |
| Captured cycle | -not set- |
| Acquisition window | 800 ms |
| Trigger pulse | 1 |
| Trigger delay | 400 ms |
| TR | 260.22 ms |
| Concatenations | 1 |
| Segments | 84 |
| Phases | 1 |
| Adaptive Triggering | Off |

#### Physio - Cardiac

| | |
|---|---|
| Tagging | None |
| Magn. preparation | None |
| Fat suppr. | None |
| Dark blood | Off |
| FoV read | 380 mm |
| FoV phase | 87.5 % |
| Phase resolution | 64 % |
| Cine | Off |
| Trajectory | Cartesian |
| Dummy heartbeats | 0 |

#### Physio - PACE

| | |
|---|---|
| Resp. control | Off |

#### Physio - PACE

| | |
|---|---|
| Concatenations | 1 |

#### Inline - Common

| | |
|---|---|
| Subtract | Off |
| Measurements | 1 |
| StdDev | Off |
| Save original images | On |

#### Inline - Cardiac

| | |
|---|---|
| Inline Evaluation | Off |
| Magn. preparation | None |
| Contrasts | 1 |
| TE | 1.16 ms |
| TR | 260.22 ms |
| Save original images | On |

#### Inline - MIP

| | |
|---|---|
| MIP-Sag | Off |
| MIP-Cor | Off |
| MIP-Tra | Off |
| MIP-Time | Off |
| Save original images | On |

#### Inline - Composing

| | |
|---|---|
| Inline Composing | Off |
| Distortion Corr. | On |
| Mode | 2D |
| Unfiltered images | Off |

#### Sequence - Part 1

| | |
|---|---|
| Introduction | Off |
| Dimension | 2D |
| Reordering | Linear |
| Asymmetric echo | Weak |
| Contrasts | 1 |
| Optimization | Min. TE |
| Multi-slice mode | Sequential |
| Echo spacing | 2.7 ms |
| Sequence type | Trufi |
| Bandwidth | 1149 Hz/Px |

#### Sequence - Part 2

| | |
|---|---|
| Define | Shots |
| Shots per slice | 1 |
| Segments | 84 |
| Trufi delta freq. | 0 Hz |
| RF pulse type | Fast |
| Gradient mode | Fast |
| Excitation | Slice-sel. |
| Flip angle mode | Constant |
| Cine | Off |

#### Sequence - Assistant

| | |
|---|---|
| Mode | Min flip angle |
| Min flip angle | 50 deg |
| Allowed delay | 0 s |

**\\USER\Cardiac Research Protocols\IV IRON\IV IRON4\trufi_4-chamber_iPAT**

TA: 0.8 s PM: REF Voxel size: 1.5×1.5×8.0 mmPAT: 2 Rel. SNR: 1.00 : tfi

#### Properties

| | |
|---|---|
| Prio recon | Off |
| Load images to viewer | On |
| Inline movie | Off |
| Auto store images | On |
| Load images to stamp segments | Off |
| Load images to graphic segments | On |
| Auto open inline display | Off |
| Auto close inline display | Off |
| Start measurement without further preparation | Off |
| Wait for user to start | Off |
| Start measurements | Single measurement |

#### Routine

| | |
|---|---|
| Slice group | 1 |
| Slices | 1 |
| Dist. factor | 20 % |
| Position | R9.1 P7.9 F6.1 mm |
| Orientation | T > S-14.6 > C12.4 |
| Phase enc. dir. | A >> P |
| AutoAlign | --- |
| Phase oversampling | 0 % |
| FoV read | 380 mm |
| FoV phase | 93.8 % |
| Slice thickness | 8.0 mm |
| TR | 275.34 ms |
| TE | 1.16 ms |
| Averages | 1 |
| Concatenations | 1 |
| Filter | Distortion Corr.(2D), Prescan Normalize |
| Coil elements | B2-4;SP4,5 |

#### Contrast - Common

| | |
|---|---|
| TR | 275.34 ms |
| TE | 1.16 ms |
| Magn. preparation | None |
| Flip angle | 80 deg |
| Fat suppr. | None |
| Wrap-up Magn. | Restore |

#### Contrast - Dynamic

| | |
|---|---|
| Averages | 1 |
| Averaging mode | Short term |
| Reconstruction | Magnitude |
| Measurements | 1 |
| Multiple series | Each measurement |

#### Resolution - Common

| | |
|---|---|
| FoV read | 380 mm |
| FoV phase | 93.8 % |
| Slice thickness | 8.0 mm |
| Base resolution | 256 |
| Phase resolution | 60 % |
| Phase partial Fourier | Off |
| Trajectory | Cartesian |
| Interpolation | Off |

#### Resolution - iPAT

| | |
|---|---|
| PAT mode | GRAPPA |

#### Resolution - iPAT

| | |
|---|---|
| Accel. factor PE | 2 |
| Ref. lines PE | 24 |
| Reference scan mode | Integrated |

#### Resolution - Filter Image

| | |
|---|---|
| Image Filter | Off |
| Distortion Corr. | On |
| Mode | 2D |
| Unfiltered images | Off |
| Prescan Normalize | On |
| Unfiltered images | Off |
| Normalize | Off |
| B1 filter | Off |

#### Resolution - Filter Rawdata

| | |
|---|---|
| Raw filter | Off |
| Elliptical filter | Off |
| POCS | Off |

#### Geometry - Common

| | |
|---|---|
| Slice group | 1 |
| Slices | 1 |
| Dist. factor | 20 % |
| Position | R9.1 P7.9 F6.1 mm |
| Orientation | T > S-14.6 > C12.4 |
| Phase enc. dir. | A >> P |
| FoV read | 380 mm |
| FoV phase | 93.8 % |
| Slice thickness | 8.0 mm |
| TR | 275.34 ms |
| Multi-slice mode | Sequential |
| Series | Interleaved |
| Concatenations | 1 |

#### Geometry - AutoAlign

| | |
|---|---|
| Slice group | 1 |
| Position | R9.1 P7.9 F6.1 mm |
| Orientation | T > S-14.6 > C12.4 |
| Phase enc. dir. | A >> P |
| AutoAlign | --- |
| Initial Position | R9.1 P7.9 F6.1 |
| Phase | 6.0 mm |
| Read | 7.3 mm |
| Shift | -9.7 mm |
| Initial Rotation | 3.20 deg |
| Initial Orientation | T > S |
| T > S | -14.6 |
| > C | 12.4 |

#### Geometry - Saturation

| | |
|---|---|
| Fat suppr. | None |
| Wrap-up Magn. | Restore |
| Special sat. | None |

#### Geometry - Navigator

#### System - Miscellaneous

| | |
|---|---|
| Positioning mode | REF |
| Table position | H |
| Table position | 0 mm |

#### System - Miscellaneous

| | |
|---|---|
| MSMA | S - C - T |
| Sagittal | R >> L |
| Coronal | A >> P |
| Transversal | F >> H |
| Coil Combine Mode | Adaptive Combine |
| Save uncombined | Off |
| Matrix Optimization | Off |
| Coil Focus | Flat |
| AutoAlign | --- |
| Coil Select Mode | Default |

#### System - Adjustments

| | |
|---|---|
| B0 Shim mode | Tune up |
| Adjust with body coil | Off |
| Confirm freq. adjustment | Off |
| Assume Dominant Fat | Off |
| Assume Silicone | Off |
| Adjustment Tolerance | Auto |

#### System - Adjust Volume

| | |
|---|---|
| Position | Isocenter |
| Orientation | Transversal |
| Rotation | 0.00 deg |
| A >> P | 263 mm |
| R >> L | 350 mm |
| F >> H | 350 mm |
| Reset | Off |

#### System - Tx/Rx

| | |
|---|---|
| Frequency 1H | 63.678323 MHz |
| Correction factor | 1 |
| Gain | High |
| Img. Scale Cor. | 1.000 |
| Reset | Off |
| ? Ref. amplitude 1H | 0.000 V |

#### Physio - Signal1

| | |
|---|---|
| 1st Signal/Mode | ECG/Trigger |
| Average cycle | No Signal ms |
| Average cycle | No Signal ms |
| Captured cycle | -not set- |
| Acquisition window | 800 ms |
| Trigger pulse | 1 |
| Trigger delay | 400 ms |
| TR | 275.34 ms |
| Concatenations | 1 |
| Segments | 84 |
| Phases | 1 |
| Adaptive Triggering | Off |

#### Physio - Cardiac

| | |
|---|---|
| Tagging | None |
| Magn. preparation | None |
| Fat suppr. | None |
| Dark blood | Off |
| FoV read | 380 mm |
| FoV phase | 93.8 % |
| Phase resolution | 60 % |
| Cine | Off |
| Trajectory | Cartesian |
| Dummy heartbeats | 0 |

#### Physio - PACE

| | |
|---|---|
| Resp. control | Off |
| Concatenations | 1 |

#### Inline - Common

| | |
|---|---|
| Subtract | Off |
| Measurements | 1 |
| StdDev | Off |
| Save original images | On |

#### Inline - Cardiac

| | |
|---|---|
| Inline Evaluation | Off |
| Magn. preparation | None |
| Contrasts | 1 |
| TE | 1.16 ms |
| TR | 275.34 ms |
| Save original images | On |

#### Inline - MIP

| | |
|---|---|
| MIP-Sag | Off |
| MIP-Cor | Off |
| MIP-Tra | Off |
| MIP-Time | Off |
| Save original images | On |

#### Inline - Composing

| | |
|---|---|
| Inline Composing | Off |
| Distortion Corr. | On |
| Mode | 2D |
| Unfiltered images | Off |

#### Sequence - Part 1

| | |
|---|---|
| Introduction | Off |
| Dimension | 2D |
| Reordering | Linear |
| Asymmetric echo | Weak |
| Contrasts | 1 |
| Optimization | Min. TE |
| Multi-slice mode | Sequential |
| Echo spacing | 2.7 ms |
| Sequence type | Trufi |
| Bandwidth | 1149 Hz/Px |

#### Sequence - Part 2

| | |
|---|---|
| Define | Shots |
| Shots per slice | 1 |
| Segments | 84 |
| Trufi delta freq. | 0 Hz |
| RF pulse type | Fast |
| Gradient mode | Fast |
| Excitation | Slice-sel. |
| Flip angle mode | Constant |
| Cine | Off |

#### Sequence - Assistant

| | |
|---|---|
| Mode | Min flip angle |
| Min flip angle | 50 deg |
| Allowed delay | 0 s |

| \\USER\Cardiac Research Protocols\IV IRON\IV IRON4\trufi_shortaxis_iPAT |
|---|
| TA: 5.4 s PM: REF Voxel size: 1.5×1.5×8.0 mmPAT: 2 Rel. SNR: 1.00 : tfi |

#### Properties

| | |
|---|---|
| Prio recon | Off |
| Load images to viewer | On |
| Inline movie | Off |
| Auto store images | On |
| Load images to stamp segments | Off |
| Load images to graphic segments | On |
| Auto open inline display | Off |
| Auto close inline display | Off |
| Start measurement without further preparation | Off |
| Wait for user to start | Off |
| Start measurements | Single measurement |

#### Routine

| | |
|---|---|
| Slice group | 1 |
| Slices | 7 |
| Dist. factor | 100 % |
| Position | R10.3 P40.1 F36.0 mm |
| Orientation | Coronal |
| Phase enc. dir. | R >> L |
| AutoAlign | --- |
| Phase oversampling | 0 % |
| FoV read | 380 mm |
| FoV phase | 87.5 % |
| Slice thickness | 8.0 mm |
| TR | 275.34 ms |
| TE | 1.16 ms |
| Averages | 1 |
| Concatenations | 7 |
| Filter | Distortion Corr.(2D), Prescan Normalize |
| Coil elements | B1-5;SP3-6 |

#### Contrast - Common

| | |
|---|---|
| TR | 275.34 ms |
| TE | 1.16 ms |
| TD | 0 ms |
| Magn. preparation | None |
| Flip angle | 80 deg |
| Fat suppr. | None |
| Wrap-up Magn. | Restore |

#### Contrast - Dynamic

| | |
|---|---|
| Averages | 1 |
| Averaging mode | Short term |
| Reconstruction | Magnitude |
| Measurements | 1 |
| Multiple series | Each measurement |

#### Resolution - Common

| | |
|---|---|
| FoV read | 380 mm |
| FoV phase | 87.5 % |
| Slice thickness | 8.0 mm |
| Base resolution | 256 |
| Phase resolution | 64 % |
| Phase partial Fourier | Off |
| Trajectory | Cartesian |
| Interpolation | Off |

#### Resolution - iPAT

| | |
|---|---|
| PAT mode | GRAPPA |
| Accel. factor PE | 2 |
| Ref. lines PE | 24 |
| Reference scan mode | Integrated |

#### Resolution - Filter Image

| | |
|---|---|
| Image Filter | Off |
| Distortion Corr. | On |
| Mode | 2D |
| Unfiltered images | Off |
| Prescan Normalize | On |
| Unfiltered images | Off |
| Normalize | Off |
| B1 filter | Off |

#### Resolution - Filter Rawdata

| | |
|---|---|
| Raw filter | Off |
| Elliptical filter | Off |
| POCS | Off |

#### Geometry - Common

| | |
|---|---|
| Slice group | 1 |
| Slices | 7 |
| Dist. factor | 100 % |
| Position | R10.3 P40.1 F36.0 mm |
| Orientation | Coronal |
| Phase enc. dir. | R >> L |
| FoV read | 380 mm |
| FoV phase | 87.5 % |
| Slice thickness | 8.0 mm |
| TR | 275.34 ms |
| Multi-slice mode | Sequential |
| Series | Descending |
| Concatenations | 7 |

#### Geometry - AutoAlign

| | |
|---|---|
| Slice group | 1 |
| Position | R10.3 P40.1 F36.0 mm |
| Orientation | Coronal |
| Phase enc. dir. | R >> L |
| AutoAlign | --- |
| Initial Position | R10.3 P40.1 F88.0 |
| Phase | -10.3 mm |
| Read | 88.0 mm |
| Shift | 40.1 mm |
| Initial Rotation | 0.00 deg |
| Initial Orientation | Coronal |

#### Geometry - Saturation

| | |
|---|---|
| Fat suppr. | None |
| Wrap-up Magn. | Restore |
| Special sat. | None |

#### Geometry - Navigator

#### System - Miscellaneous

| | |
|---|---|
| Positioning mode | REF |
| Table position | H |
| Table position | 52 mm |
| MSMA | S - C - T |

#### System - Miscellaneous

| | |
|---|---|
| Sagittal | R >> L |
| Coronal | A >> P |
| Transversal | F >> H |
| Coil Combine Mode | Adaptive Combine |
| Save uncombined | Off |
| Matrix Optimization | Off |
| Coil Focus | Flat |
| AutoAlign | --- |
| Coil Select Mode | Default |

#### System - Adjustments

| | |
|---|---|
| B0 Shim mode | Tune up |
| Adjust with body coil | Off |
| Confirm freq. adjustment | Off |
| Assume Dominant Fat | Off |
| Assume Silicone | Off |
| Adjustment Tolerance | Auto |

#### System - Adjust Volume

| | |
|---|---|
| Position | Isocenter |
| Orientation | Transversal |
| Rotation | 0.00 deg |
| A >> P | 263 mm |
| R >> L | 350 mm |
| F >> H | 350 mm |
| Reset | Off |

#### System - Tx/Rx

| | |
|---|---|
| Frequency 1H | 63.678323 MHz |
| Correction factor | 1 |
| Gain | High |
| Img. Scale Cor. | 1.000 |
| Reset | Off |
| ? Ref. amplitude 1H | 0.000 V |

#### Physio - Signal1

| | |
|---|---|
| 1st Signal/Mode | ECG/Trigger |
| Average cycle | No Signal ms |
| Average cycle | No Signal ms |
| Captured cycle | -not set- |
| Acquisition window | 774 ms |
| Trigger pulse | 1 |
| Trigger delay | 498 ms |
| TR | 275.34 ms |
| Concatenations | 7 |
| Segments | 84 |
| Phases | 1 |
| Adaptive Triggering | Off |

#### Physio - Cardiac

| | |
|---|---|
| Tagging | None |
| Magn. preparation | None |
| Fat suppr. | None |
| Dark blood | Off |
| FoV read | 380 mm |
| FoV phase | 87.5 % |
| Phase resolution | 64 % |
| Cine | Off |
| Trajectory | Cartesian |
| Dummy heartbeats | 0 |

#### Physio - PACE

| | |
|---|---|
| Resp. control | Off |
| Concatenations | 7 |

#### Inline - Common

| | |
|---|---|
| Subtract | Off |
| Measurements | 1 |
| StdDev | Off |
| Save original images | On |

#### Inline - Cardiac

| | |
|---|---|
| Inline Evaluation | Off |
| Magn. preparation | None |
| Contrasts | 1 |
| TE | 1.16 ms |
| TR | 275.34 ms |
| Save original images | On |

#### Inline - MIP

| | |
|---|---|
| MIP-Sag | Off |
| MIP-Cor | Off |
| MIP-Tra | Off |
| MIP-Time | Off |
| Save original images | On |

#### Inline - Composing

| | |
|---|---|
| Inline Composing | Off |
| Distortion Corr. | On |
| Mode | 2D |
| Unfiltered images | Off |

#### Sequence - Part 1

| | |
|---|---|
| Introduction | Off |
| Dimension | 2D |
| Reordering | Linear |
| Asymmetric echo | Weak |
| Contrasts | 1 |
| Optimization | Min. TE |
| Multi-slice mode | Sequential |
| Echo spacing | 2.7 ms |
| Sequence type | Trufi |
| Bandwidth | 1149 Hz/Px |

#### Sequence - Part 2

| | |
|---|---|
| Define | Shots |
| Shots per slice | 1 |
| Segments | 84 |
| Trufi delta freq. | 0 Hz |
| RF pulse type | Fast |
| Gradient mode | Fast |
| Excitation | Slice-sel. |
| Flip angle mode | Constant |
| Cine | Off |

#### Sequence - Assistant

| | |
|---|---|
| Mode | Min flip angle |
| Min flip angle | 50 deg |
| Allowed delay | 0 s |

**\\USER\Cardiac Research Protocols\IV IRON\IV IRON4\HLA tf2d15_retro_iPAT3**

TA: 5.0 s PM: REF Voxel size: 1.9×1.9×8.0 mmPAT: 3 Rel. SNR: 1.00 : tfi

#### Properties

| | |
|---|---|
| Prio recon | Off |
| Load images to viewer | On |
| Inline movie | On |
| Auto store images | On |
| Load images to stamp segments | Off |
| Load images to graphic segments | On |
| Auto open inline display | Off |
| Auto close inline display | Off |
| Start measurement without further preparation | Off |
| Wait for user to start | Off |
| Start measurements | Single measurement |

#### Routine

| | |
|---|---|
| Slice group | 1 |
| Slices | 1 |
| Dist. factor | 25 % |
| Position | L46.1 A28.2 H1.7 mm |
| Orientation | T > C28.3 > S5.5 |
| Phase enc. dir. | A >> P |
| AutoAlign | --- |
| Phase oversampling | 0 % |
| FoV read | 360 mm |
| FoV phase | 100.0 % |
| Slice thickness | 8.0 mm |
| TR | 40.65 ms |
| TE | 1.15 ms |
| Averages | 1 |
| Concatenations | 1 |
| Filter | Distortion Corr.(2D), Prescan Normalize, Image Filter |
| Coil elements | B2-4;SP4,5 |

#### Contrast - Common

| | |
|---|---|
| TR | 40.65 ms |
| TE | 1.15 ms |
| Magn. preparation | None |
| Flip angle | 53 deg |
| Fat suppr. | None |
| Wrap-up Magn. | Restore |

#### Contrast - Dynamic

| | |
|---|---|
| Averages | 1 |
| Averaging mode | Short term |
| Reconstruction | Magnitude |
| Measurements | 1 |
| Multiple series | Each slice |

#### Resolution - Common

| | |
|---|---|
| FoV read | 360 mm |
| FoV phase | 100.0 % |
| Slice thickness | 8.0 mm |
| Base resolution | 192 |
| Phase resolution | 100 % |
| Phase partial Fourier | Off |
| Trajectory | Cartesian |
| View sharing | Off |
| Interpolation | Off |

#### Resolution - iPAT

| | |
|---|---|
| PAT mode | GRAPPA |
| Accel. factor PE | 3 |
| Ref. lines PE | 24 |
| Reference scan mode | GRE/separate |

#### Resolution - Filter Image

| | |
|---|---|
| Image Filter | On |
| ! Intensity | Medium |
| Edge Enhancement | 2 |
| Smoothing | 2 |
| Unfiltered images | Off |
| Distortion Corr. | On |
| Mode | 2D |
| Unfiltered images | Off |
| Prescan Normalize | On |
| Unfiltered images | Off |
| Normalize | Off |
| B1 filter | Off |

#### Resolution - Filter Rawdata

| | |
|---|---|
| Raw filter | Off |
| Elliptical filter | Off |
| POCS | Off |

#### Geometry - Common

| | |
|---|---|
| Slice group | 1 |
| Slices | 1 |
| Dist. factor | 25 % |
| Position | L46.1 A28.2 H1.7 mm |
| Orientation | T > C28.3 > S5.5 |
| Phase enc. dir. | A >> P |
| FoV read | 360 mm |
| FoV phase | 100.0 % |
| Slice thickness | 8.0 mm |
| TR | 40.65 ms |
| Multi-slice mode | Sequential |
| Series | Base To Apex |
| Concatenations | 1 |

#### Geometry - AutoAlign

| | |
|---|---|
| Slice group | 1 |
| Position | L46.1 A28.2 H1.7 mm |
| Orientation | T > C28.3 > S5.5 |
| Phase enc. dir. | A >> P |
| AutoAlign | --- |
| Initial Position | L46.1 A28.2 H1.7 |
| Phase | -20.4 mm |
| Read | -48.9 mm |
| Shift | 10.4 mm |
| Initial Rotation | 4.22 deg |
| Initial Orientation | T > C |
| T > C | 28.3 |
| > S | 5.5 |

#### Geometry - Saturation

| | |
|---|---|
| Fat suppr. | None |
| Wrap-up Magn. | Restore |
| Special sat. | None |

#### Geometry - Navigator

#### System - Miscellaneous

| | |
|---|---|
| Positioning mode | REF |
| Table position | H |
| Table position | 0 mm |
| MSMA | S - C - T |
| Sagittal | R >> L |
| Coronal | A >> P |
| Transversal | F >> H |
| Coil Combine Mode | Adaptive Combine |
| Save uncombined | Off |
| Matrix Optimization | Off |
| Coil Focus | Flat |
| AutoAlign | --- |
| Coil Select Mode | Default |

#### System - Adjustments

| | |
|---|---|
| B0 Shim mode | Tune up |
| Adjust with body coil | On |
| Confirm freq. adjustment | Off |
| Assume Dominant Fat | Off |
| Assume Silicone | Off |
| Adjustment Tolerance | Auto |

#### System - Adjust Volume

| | |
|---|---|
| Position | Isocenter |
| Orientation | Transversal |
| Rotation | 0.00 deg |
| A >> P | 263 mm |
| R >> L | 350 mm |
| F >> H | 350 mm |
| Reset | Off |

#### System - Tx/Rx

| | |
|---|---|
| Frequency 1H | 63.678323 MHz |
| Correction factor | 1 |
| Gain | High |
| Img. Scale Cor. | 1.000 |
| Reset | Off |
| ? Ref. amplitude 1H | 0.000 V |

#### Physio - Signal1

| | |
|---|---|
| 1st Signal/Mode | ECG/Retro |
| Average cycle | No Signal ms |
| Average cycle | No Signal ms |
| Calculated phases | 25 |
| TR | 40.65 ms |
| Concatenations | 1 |
| Segments | 15 |
| Arrhythmia detection | None |

#### Physio - Cardiac

| | |
|---|---|
| Tagging | None |
| Magn. preparation | None |
| Fat suppr. | None |
| Dark blood | Off |
| FoV read | 360 mm |
| FoV phase | 100.0 % |
| Phase resolution | 100 % |
| Cine | On |
| Trajectory | Cartesian |
| View sharing | Off |
| Dummy heartbeats | 1 |

#### Physio - PACE

| | |
|---|---|
| Resp. control | Breath-hold |

#### Physio - PACE

| | |
|---|---|
| Concatenations | 1 |

#### Inline - Common

| | |
|---|---|
| Subtract | Off |
| Measurements | 1 |
| StdDev | Off |
| Save original images | On |

#### Inline - Cardiac

| | |
|---|---|
| Inline Evaluation | Off |
| Magn. preparation | None |
| Contrasts | 1 |
| TE | 1.15 ms |
| TR | 40.65 ms |
| Save original images | On |

#### Inline - MIP

| | |
|---|---|
| MIP-Sag | Off |
| MIP-Cor | Off |
| MIP-Tra | Off |
| MIP-Time | Off |
| Save original images | On |

#### Inline - Composing

| | |
|---|---|
| Inline Composing | Off |
| Distortion Corr. | On |
| Mode | 2D |
| Unfiltered images | Off |

#### Sequence - Part 1

| | |
|---|---|
| Introduction | Off |
| Dimension | 2D |
| Reordering | Linear |
| Asymmetric echo | Weak |
| Contrasts | 1 |
| Optimization | Min. TE TR |
| Multi-slice mode | Sequential |
| Echo spacing | 2.7 ms |
| Sequence type | Trufi |
| Bandwidth | 930 Hz/Px |

#### Sequence - Part 2

| | |
|---|---|
| Define | Segments |
| Segments | 15 |
| Trufi delta freq. | 0 Hz |
| RF pulse type | Fast |
| Gradient mode | Fast* |
| Excitation | Slice-sel. |
| Flip angle mode | Constant |
| Cine | On |

#### Sequence - Assistant

| | |
|---|---|
| Mode | Off |
| Allowed delay | 0 s |

**\\USER\Cardiac Research Protocols\IV IRON\IV IRON4\VLA tf2d15_retro_iPAT3**

TA: 6.0 s PM: REF Voxel size: 1.9×1.9×8.0 mmPAT: 3 Rel. SNR: 1.00 : tfi

#### Properties

| | |
|---|---|
| Prio recon | Off |
| Load images to viewer | On |
| Inline movie | On |
| Auto store images | On |
| Load images to stamp segments | Off |
| Load images to graphic segments | On |
| Auto open inline display | Off |
| Auto close inline display | Off |
| Start measurement without further preparation | Off |
| Wait for user to start | Off |
| Start measurements | Single measurement |

#### Routine

| | |
|---|---|
| Slice group | 1 |
| Slices | 1 |
| Dist. factor | 25 % |
| Position | L13.6 P5.0 H2.3 mm |
| Orientation | C > S-41.6 > T-12.5 |
| Phase enc. dir. | R >> L |
| AutoAlign | --- |
| Phase oversampling | 0 % |
| FoV read | 360 mm |
| FoV phase | 100.0 % |
| Slice thickness | 8.0 mm |
| TR | 40.65 ms |
| TE | 1.15 ms |
| Averages | 1 |
| Concatenations | 1 |
| Filter | Distortion Corr.(2D), Prescan Normalize, Image Filter |
| Coil elements | B1-5;SP3-6 |

#### Contrast - Common

| | |
|---|---|
| TR | 40.65 ms |
| TE | 1.15 ms |
| Magn. preparation | None |
| Flip angle | 53 deg |
| Fat suppr. | None |
| Wrap-up Magn. | Restore |

#### Contrast - Dynamic

| | |
|---|---|
| Averages | 1 |
| Averaging mode | Short term |
| Reconstruction | Magnitude |
| Measurements | 1 |
| Multiple series | Each slice |

#### Resolution - Common

| | |
|---|---|
| FoV read | 360 mm |
| FoV phase | 100.0 % |
| Slice thickness | 8.0 mm |
| Base resolution | 192 |
| Phase resolution | 100 % |
| Phase partial Fourier | Off |
| Trajectory | Cartesian |
| View sharing | Off |
| Interpolation | Off |

#### Resolution - iPAT

| | |
|---|---|
| PAT mode | GRAPPA |
| Accel. factor PE | 3 |
| Ref. lines PE | 24 |
| Reference scan mode | GRE/separate |

#### Resolution - Filter Image

| | |
|---|---|
| Image Filter | On |
| ! Intensity | Medium |
| Edge Enhancement | 2 |
| Smoothing | 2 |
| Unfiltered images | Off |
| Distortion Corr. | On |
| Mode | 2D |
| Unfiltered images | Off |
| Prescan Normalize | On |
| Unfiltered images | Off |
| Normalize | Off |
| B1 filter | Off |

#### Resolution - Filter Rawdata

| | |
|---|---|
| Raw filter | Off |
| Elliptical filter | Off |
| POCS | Off |

#### Geometry - Common

| | |
|---|---|
| Slice group | 1 |
| Slices | 1 |
| Dist. factor | 25 % |
| Position | L13.6 P5.0 H2.3 mm |
| Orientation | C > S-41.6 > T-12.5 |
| Phase enc. dir. | R >> L |
| FoV read | 360 mm |
| FoV phase | 100.0 % |
| Slice thickness | 8.0 mm |
| TR | 40.65 ms |
| Multi-slice mode | Sequential |
| Series | Base To Apex |
| Concatenations | 1 |

#### Geometry - AutoAlign

| | |
|---|---|
| Slice group | 1 |
| Position | L13.6 P5.0 H2.3 mm |
| Orientation | C > S-41.6 > T-12.5 |
| Phase enc. dir. | R >> L |
| AutoAlign | --- |
| Initial Position | L13.6 P5.0 H2.3 |
| Phase | 6.8 mm |
| Read | 1.0 mm |
| Shift | 13.0 mm |
| Initial Rotation | 4.22 deg |
| Initial Orientation | C > S |
| C > S | -41.6 |
| > T | -12.5 |

#### Geometry - Saturation

| | |
|---|---|
| Fat suppr. | None |
| Wrap-up Magn. | Restore |
| Special sat. | None |

#### Geometry - Navigator

#### System - Miscellaneous

| | |
|---|---|
| Positioning mode | REF |
| Table position | H |
| Table position | 0 mm |
| MSMA | S - C - T |
| Sagittal | R >> L |
| Coronal | A >> P |
| Transversal | F >> H |
| Coil Combine Mode | Adaptive Combine |
| Save uncombined | Off |
| Matrix Optimization | Off |
| Coil Focus | Flat |
| AutoAlign | --- |
| Coil Select Mode | Default |

#### System - Adjustments

| | |
|---|---|
| B0 Shim mode | Tune up |
| Adjust with body coil | On |
| Confirm freq. adjustment | Off |
| Assume Dominant Fat | Off |
| Assume Silicone | Off |
| Adjustment Tolerance | Auto |

#### System - Adjust Volume

| | |
|---|---|
| Position | Isocenter |
| Orientation | Transversal |
| Rotation | 0.00 deg |
| A >> P | 263 mm |
| R >> L | 350 mm |
| F >> H | 350 mm |
| Reset | Off |

#### System - Tx/Rx

| | |
|---|---|
| Frequency 1H | 63.678323 MHz |
| Correction factor | 1 |
| Gain | High |
| Img. Scale Cor. | 1.000 |
| Reset | Off |
| ? Ref. amplitude 1H | 0.000 V |

#### Physio - Signal1

| | |
|---|---|
| 1st Signal/Mode | ECG/Retro |
| Average cycle | No Signal ms |
| Average cycle | No Signal ms |
| Calculated phases | 25 |
| TR | 40.65 ms |
| Concatenations | 1 |
| Segments | 15 |
| Arrhythmia detection | None |

#### Physio - Cardiac

| | |
|---|---|
| Tagging | None |
| Magn. preparation | None |
| Fat suppr. | None |
| Dark blood | Off |
| FoV read | 360 mm |
| FoV phase | 100.0 % |
| Phase resolution | 100 % |
| Cine | On |
| Trajectory | Cartesian |
| View sharing | Off |
| Dummy heartbeats | 1 |

#### Physio - PACE

| | |
|---|---|
| Resp. control | Breath-hold |

#### Physio - PACE

| | |
|---|---|
| Concatenations | 1 |

#### Inline - Common

| | |
|---|---|
| Subtract | Off |
| Measurements | 1 |
| StdDev | Off |
| Save original images | On |

#### Inline - Cardiac

| | |
|---|---|
| Inline Evaluation | Off |
| Magn. preparation | None |
| Contrasts | 1 |
| TE | 1.15 ms |
| TR | 40.65 ms |
| Save original images | On |

#### Inline - MIP

| | |
|---|---|
| MIP-Sag | Off |
| MIP-Cor | Off |
| MIP-Tra | Off |
| MIP-Time | Off |
| Save original images | On |

#### Inline - Composing

| | |
|---|---|
| Inline Composing | Off |
| Distortion Corr. | On |
| Mode | 2D |
| Unfiltered images | Off |

#### Sequence - Part 1

| | |
|---|---|
| Introduction | Off |
| Dimension | 2D |
| Reordering | Linear |
| Asymmetric echo | Weak |
| Contrasts | 1 |
| Optimization | Min. TE TR |
| Multi-slice mode | Sequential |
| Echo spacing | 2.7 ms |
| Sequence type | Trufi |
| Bandwidth | 930 Hz/Px |

#### Sequence - Part 2

| | |
|---|---|
| Define | Segments |
| Segments | 15 |
| Trufi delta freq. | 0 Hz |
| RF pulse type | Fast |
| Gradient mode | Fast* |
| Excitation | Slice-sel. |
| Flip angle mode | Constant |
| Cine | On |

#### Sequence - Assistant

| | |
|---|---|
| Mode | Off |
| Allowed delay | 0 s |

**\\USER\Cardiac Research Protocols\IV IRON\IV IRON4\LVOT tf2d15_retro_iPAT3**

TA: 6.0 s PM: REF Voxel size: 1.9×1.9×8.0 mmPAT: 3 Rel. SNR: 1.00 : tfi

#### Properties

| | |
|---|---|
| Prio recon | Off |
| Load images to viewer | On |
| Inline movie | On |
| Auto store images | On |
| Load images to stamp segments | Off |
| Load images to graphic segments | On |
| Auto open inline display | Off |
| Auto close inline display | Off |
| Start measurement without further preparation | Off |
| Wait for user to start | Off |
| Start measurements | Single measurement |

#### Routine

| | |
|---|---|
| Slice group | 1 |
| Slices | 1 |
| Dist. factor | 25 % |
| Position | L12.6 P6.7 H15.9 mm |
| Orientation | T > S-17.0 > C-0.4 |
| Phase enc. dir. | A >> P |
| AutoAlign | --- |
| Phase oversampling | 0 % |
| FoV read | 360 mm |
| FoV phase | 100.0 % |
| Slice thickness | 8.0 mm |
| TR | 40.65 ms |
| TE | 1.15 ms |
| Averages | 1 |
| Concatenations | 1 |
| Filter | Distortion Corr.(2D), Prescan Normalize, Image Filter |
| Coil elements | B2,3;SP4 |

#### Contrast - Common

| | |
|---|---|
| TR | 40.65 ms |
| TE | 1.15 ms |
| Magn. preparation | None |
| Flip angle | 53 deg |
| Fat suppr. | None |
| Wrap-up Magn. | Restore |

#### Contrast - Dynamic

| | |
|---|---|
| Averages | 1 |
| Averaging mode | Short term |
| Reconstruction | Magnitude |
| Measurements | 1 |
| Multiple series | Each slice |

#### Resolution - Common

| | |
|---|---|
| FoV read | 360 mm |
| FoV phase | 100.0 % |
| Slice thickness | 8.0 mm |
| Base resolution | 192 |
| Phase resolution | 100 % |
| Phase partial Fourier | Off |
| Trajectory | Cartesian |
| View sharing | Off |
| Interpolation | Off |

#### Resolution - iPAT

| | |
|---|---|
| PAT mode | GRAPPA |
| Accel. factor PE | 3 |
| Ref. lines PE | 24 |
| Reference scan mode | GRE/separate |

#### Resolution - Filter Image

| | |
|---|---|
| Image Filter | On |
| ! Intensity | Medium |
| Edge Enhancement | 2 |
| Smoothing | 2 |
| Unfiltered images | Off |
| Distortion Corr. | On |
| Mode | 2D |
| Unfiltered images | Off |
| Prescan Normalize | On |
| Unfiltered images | Off |
| Normalize | Off |
| B1 filter | Off |

#### Resolution - Filter Rawdata

| | |
|---|---|
| Raw filter | Off |
| Elliptical filter | Off |
| POCS | Off |

#### Geometry - Common

| | |
|---|---|
| Slice group | 1 |
| Slices | 1 |
| Dist. factor | 25 % |
| Position | L12.6 P6.7 H15.9 mm |
| Orientation | T > S-17.0 > C-0.4 |
| Phase enc. dir. | A >> P |
| FoV read | 360 mm |
| FoV phase | 100.0 % |
| Slice thickness | 8.0 mm |
| TR | 40.65 ms |
| Multi-slice mode | Sequential |
| Series | Base To Apex |
| Concatenations | 1 |

#### Geometry - AutoAlign

| | |
|---|---|
| Slice group | 1 |
| Position | L12.6 P6.7 H15.9 mm |
| Orientation | T > S-17.0 > C-0.4 |
| Phase enc. dir. | A >> P |
| AutoAlign | --- |
| Initial Position | L12.6 P6.7 H15.9 |
| Phase | 7.1 mm |
| Read | -6.9 mm |
| Shift | 18.9 mm |
| Initial Rotation | 4.22 deg |
| Initial Orientation | T > S |
| T > S | -17.0 |
| > C | -0.4 |

#### Geometry - Saturation

| | |
|---|---|
| Fat suppr. | None |
| Wrap-up Magn. | Restore |
| Special sat. | None |

#### Geometry - Navigator

#### System - Miscellaneous

| | |
|---|---|
| Positioning mode | REF |
| Table position | H |
| Table position | 0 mm |
| MSMA | S - C - T |
| Sagittal | R >> L |
| Coronal | A >> P |
| Transversal | F >> H |
| Coil Combine Mode | Adaptive Combine |
| Save uncombined | Off |
| Matrix Optimization | Off |
| Coil Focus | Flat |
| AutoAlign | --- |
| Coil Select Mode | Default |

#### System - Adjustments

| | |
|---|---|
| B0 Shim mode | Tune up |
| Adjust with body coil | On |
| Confirm freq. adjustment | Off |
| Assume Dominant Fat | Off |
| Assume Silicone | Off |
| Adjustment Tolerance | Auto |

#### System - Adjust Volume

| | |
|---|---|
| Position | Isocenter |
| Orientation | Transversal |
| Rotation | 0.00 deg |
| A >> P | 263 mm |
| R >> L | 350 mm |
| F >> H | 350 mm |
| Reset | Off |

#### System - Tx/Rx

| | |
|---|---|
| Frequency 1H | 63.678323 MHz |
| Correction factor | 1 |
| Gain | High |
| Img. Scale Cor. | 1.000 |
| Reset | Off |
| ? Ref. amplitude 1H | 0.000 V |

#### Physio - Signal1

| | |
|---|---|
| 1st Signal/Mode | ECG/Retro |
| Average cycle | No Signal ms |
| Average cycle | No Signal ms |
| Calculated phases | 25 |
| TR | 40.65 ms |
| Concatenations | 1 |
| Segments | 15 |
| Arrhythmia detection | None |

#### Physio - Cardiac

| | |
|---|---|
| Tagging | None |
| Magn. preparation | None |
| Fat suppr. | None |
| Dark blood | Off |
| FoV read | 360 mm |
| FoV phase | 100.0 % |
| Phase resolution | 100 % |
| Cine | On |
| Trajectory | Cartesian |
| View sharing | Off |
| Dummy heartbeats | 1 |

#### Physio - PACE

| | |
|---|---|
| Resp. control | Breath-hold |

#### Physio - PACE

| | |
|---|---|
| Concatenations | 1 |

#### Inline - Common

| | |
|---|---|
| Subtract | Off |
| Measurements | 1 |
| StdDev | Off |
| Save original images | On |

#### Inline - Cardiac

| | |
|---|---|
| Inline Evaluation | Off |
| Magn. preparation | None |
| Contrasts | 1 |
| TE | 1.15 ms |
| TR | 40.65 ms |
| Save original images | On |

#### Inline - MIP

| | |
|---|---|
| MIP-Sag | Off |
| MIP-Cor | Off |
| MIP-Tra | Off |
| MIP-Time | Off |
| Save original images | On |

#### Inline - Composing

| | |
|---|---|
| Inline Composing | Off |
| Distortion Corr. | On |
| Mode | 2D |
| Unfiltered images | Off |

#### Sequence - Part 1

| | |
|---|---|
| Introduction | Off |
| Dimension | 2D |
| Reordering | Linear |
| Asymmetric echo | Weak |
| Contrasts | 1 |
| Optimization | Min. TE TR |
| Multi-slice mode | Sequential |
| Echo spacing | 2.7 ms |
| Sequence type | Trufi |
| Bandwidth | 930 Hz/Px |

#### Sequence - Part 2

| | |
|---|---|
| Define | Segments |
| Segments | 15 |
| Trufi delta freq. | 0 Hz |
| RF pulse type | Fast |
| Gradient mode | Fast* |
| Excitation | Slice-sel. |
| Flip angle mode | Constant |
| Cine | On |

#### Sequence - Assistant

| | |
|---|---|
| Mode | Off |
| Allowed delay | 0 s |

#### \\USER\Cardiac Research Protocols\IV IRON\IV IRON4\SA STACK tf2d15_retro_iPAT3

TA: 2:35 PM: REF Voxel size: 1.9×1.9×8.0 mmPAT: 3 Rel. SNR: 1.00 : tfi

##### Properties

| | |
|---|---|
| Prio recon | Off |
| Load images to viewer | On |
| Inline movie | On |
| Auto store images | On |
| Load images to stamp segments | Off |
| Load images to graphic segments | On |
| Auto open inline display | Off |
| Auto close inline display | Off |
| Start measurement without further preparation | Off |
| Wait for user to start | Off |
| Start measurements | Single measurement |

##### Routine

| | |
|---|---|
| Slice group | 1 |
| Slices | 11 |
| Dist. factor | 25 % |
| Position | L54.9 A12.1 F10.8 mm |
| Orientation | S > C31.4 > T16.5 |
| Phase enc. dir. | A >> P |
| AutoAlign | --- |
| Phase oversampling | 0 % |
| FoV read | 360 mm |
| FoV phase | 90.6 % |
| Slice thickness | 8.0 mm |
| TR | 40.35 ms |
| TE | 1.13 ms |
| Averages | 1 |
| Concatenations | 11 |
| Filter | Distortion Corr.(2D), Prescan Normalize, Image Filter |
| Coil elements | B1-5;SP3-6 |

##### Contrast - Common

| | |
|---|---|
| TR | 40.35 ms |
| TE | 1.13 ms |
| Magn. preparation | None |
| Flip angle | 52 deg |
| Fat suppr. | None |
| Wrap-up Magn. | Restore |

##### Contrast - Dynamic

| | |
|---|---|
| Averages | 1 |
| Averaging mode | Short term |
| Reconstruction | Magnitude |
| Measurements | 1 |
| Multiple series | Each slice |

##### Resolution - Common

| | |
|---|---|
| FoV read | 360 mm |
| FoV phase | 90.6 % |
| Slice thickness | 8.0 mm |
| Base resolution | 192 |
| Phase resolution | 100 % |
| Phase partial Fourier | Off |
| Trajectory | Cartesian |
| View sharing | Off |
| Interpolation | Off |

##### Resolution - iPAT

| | |
|---|---|
| PAT mode | GRAPPA |
| Accel. factor PE | 3 |
| Ref. lines PE | 24 |
| Reference scan mode | GRE/separate |

##### Resolution - Filter Image

| | |
|---|---|
| Image Filter | On |
| ! Intensity | Medium |
| Edge Enhancement | 2 |
| Smoothing | 2 |
| Unfiltered images | Off |
| Distortion Corr. | On |
| Mode | 2D |
| Unfiltered images | Off |
| Prescan Normalize | On |
| Unfiltered images | Off |
| Normalize | Off |
| B1 filter | Off |

##### Resolution - Filter Rawdata

| | |
|---|---|
| Raw filter | Off |
| Elliptical filter | Off |
| POCS | Off |

##### Geometry - Common

| | |
|---|---|
| Slice group | 1 |
| Slices | 11 |
| Dist. factor | 25 % |
| Position | L54.9 A12.1 F10.8 mm |
| Orientation | S > C31.4 > T16.5 |
| Phase enc. dir. | A >> P |
| FoV read | 360 mm |
| FoV phase | 90.6 % |
| Slice thickness | 8.0 mm |
| TR | 40.35 ms |
| Multi-slice mode | Sequential |
| Series | Base To Apex |
| Concatenations | 11 |

##### Geometry - AutoAlign

| | |
|---|---|
| Slice group | 1 |
| Position | L54.9 A12.1 F10.8 mm |
| Orientation | S > C31.4 > T16.5 |
| Phase enc. dir. | A >> P |
| AutoAlign | --- |
| Initial Position | L54.9 A12.1 F10.8 |
| Phase | 17.1 mm |
| Read | 8.1 mm |
| Shift | 54.0 mm |
| Initial Rotation | 11.00 deg |
| Initial Orientation | S > C |
| S > C | 31.4 |
| > T | 16.5 |

##### Geometry - Saturation

| | |
|---|---|
| Fat suppr. | None |
| Wrap-up Magn. | Restore |
| Special sat. | None |

##### Geometry - Navigator

#### System - Miscellaneous

| | |
|---|---|
| Positioning mode | REF |
| Table position | H |
| Table position | 0 mm |
| MSMA | S - C - T |
| Sagittal | R >> L |
| Coronal | A >> P |
| Transversal | F >> H |
| Coil Combine Mode | Adaptive Combine |
| Save uncombined | Off |
| Matrix Optimization | Off |
| Coil Focus | Flat |
| AutoAlign | --- |
| Coil Select Mode | Default |

#### System - Adjustments

| | |
|---|---|
| B0 Shim mode | Tune up |
| Adjust with body coil | On |
| Confirm freq. adjustment | Off |
| Assume Dominant Fat | Off |
| Assume Silicone | Off |
| Adjustment Tolerance | Auto |

#### System - Adjust Volume

| | |
|---|---|
| Position | Isocenter |
| Orientation | Transversal |
| Rotation | 0.00 deg |
| A >> P | 263 mm |
| R >> L | 350 mm |
| F >> H | 350 mm |
| Reset | Off |

#### System - Tx/Rx

| | |
|---|---|
| Frequency 1H | 63.678323 MHz |
| Correction factor | 1 |
| Gain | High |
| Img. Scale Cor. | 1.000 |
| Reset | Off |
| ? Ref. amplitude 1H | 0.000 V |

#### Physio - Signal1

| | |
|---|---|
| 1st Signal/Mode | ECG/Retro |
| Average cycle | No Signal ms |
| Average cycle | No Signal ms |
| Calculated phases | 25 |
| TR | 40.35 ms |
| Concatenations | 11 |
| Segments | 15 |
| Arrhythmia detection | None |

#### Physio - Cardiac

| | |
|---|---|
| Tagging | None |
| Magn. preparation | None |
| Fat suppr. | None |
| Dark blood | Off |
| FoV read | 360 mm |
| FoV phase | 90.6 % |
| Phase resolution | 100 % |
| Cine | On |
| Trajectory | Cartesian |
| View sharing | Off |
| Dummy heartbeats | 1 |

#### Physio - PACE

| | |
|---|---|
| Resp. control | Breath-hold |

#### Physio - PACE

| | |
|---|---|
| Concatenations | 11 |

#### Inline - Common

| | |
|---|---|
| Subtract | Off |
| Measurements | 1 |
| StdDev | Off |
| Save original images | On |

#### Inline - Cardiac

| | |
|---|---|
| Inline Evaluation | Off |
| Magn. preparation | None |
| Contrasts | 1 |
| TE | 1.13 ms |
| TR | 40.35 ms |
| Save original images | On |

#### Inline - MIP

| | |
|---|---|
| MIP-Sag | Off |
| MIP-Cor | Off |
| MIP-Tra | Off |
| MIP-Time | Off |
| Save original images | On |

#### Inline - Composing

| | |
|---|---|
| Inline Composing | Off |
| Distortion Corr. | On |
| Mode | 2D |
| Unfiltered images | Off |

#### Sequence - Part 1

| | |
|---|---|
| Introduction | Off |
| Dimension | 2D |
| Reordering | Linear |
| Asymmetric echo | Weak |
| Contrasts | 1 |
| Optimization | Min. TE TR |
| Multi-slice mode | Sequential |
| Echo spacing | 2.7 ms |
| Sequence type | Trufi |
| Bandwidth | 930 Hz/Px |

#### Sequence - Part 2

| | |
|---|---|
| Define | Segments |
| Segments | 15 |
| Trufi delta freq. | 0 Hz |
| RF pulse type | Fast |
| Gradient mode | Fast* |
| Excitation | Slice-sel. |
| Flip angle mode | Constant |
| Cine | On |

#### Sequence - Assistant

| | |
|---|---|
| Mode | Off |
| Allowed delay | 0 s |

| \\USER\Cardiac Research Protocols\IV IRON\IV IRON4\VLS_ShMOLLI_192i_d11_nFilt |
|---|
| TA: 5.3 s PM: REF Voxel size: 0.9×0.9×8.0 mmPAT: 2 Rel. SNR: 1.00 : tfi |

#### Properties

| | |
|---|---|
| Prio recon | Off |
| Load images to viewer | On |
| Inline movie | Off |
| Auto store images | On |
| Load images to stamp segments | Off |
| Load images to graphic segments | On |
| Auto open inline display | Off |
| Auto close inline display | Off |
| Start measurement without further preparation | Off |
| Wait for user to start | Off |
| Start measurements | Single measurement |

#### Routine

| | |
|---|---|
| Slice group | 1 |
| Slices | 1 |
| Dist. factor | 25 % |
| Position | Isocenter |
| Orientation | Transversal |
| Phase enc. dir. | A >> P |
| AutoAlign | --- |
| Phase oversampling | 0 % |
| FoV read | 360 mm |
| FoV phase | 75.0 % |
| Slice thickness | 8.0 mm |
| TR | 378.98 ms |
| TE | 1.07 ms |
| Averages | 1 |
| Concatenations | 1 |
| Filter | Raw filter, Distortion Corr.(2D) |
| Coil elements | BO2;SP2,3 |

#### Contrast - Common

| | |
|---|---|
| TR | 378.98 ms |
| TE | 1.07 ms |
| Magn. preparation | Non-sel. IR |
| TI | 260 ms |
| Flip angle | 35 deg |
| Fat suppr. | None |
| Wrap-up Magn. | None |

#### Contrast - Dynamic

| | |
|---|---|
| Averages | 1 |
| Averaging mode | Short term |
| Reconstruction | Magn./Phase |
| Measurements | 1 |
| Multiple series | Off |

#### Resolution - Common

| | |
|---|---|
| FoV read | 360 mm |
| FoV phase | 75.0 % |
| Slice thickness | 8.0 mm |
| Base resolution | 192 |
| Phase resolution | 100 % |
| Phase partial Fourier | 6/8 |
| Trajectory | Cartesian |
| Interpolation | On |

#### Resolution - iPAT

| | |
|---|---|
| PAT mode | GRAPPA |
| Accel. factor PE | 2 |
| Ref. lines PE | 24 |
| Reference scan mode | Integrated |

#### Resolution - Filter Image

| | |
|---|---|
| Image Filter | Off |
| Distortion Corr. | On |
| Mode | 2D |
| Unfiltered images | Off |
| Prescan Normalize | Off |
| Normalize | Off |
| B1 filter | Off |

#### Resolution - Filter Rawdata

| | |
|---|---|
| Raw filter | On |
| Elliptical filter | Off |
| POCS | Off |

#### Geometry - Common

| | |
|---|---|
| Slice group | 1 |
| Slices | 1 |
| Dist. factor | 25 % |
| Position | Isocenter |
| Orientation | Transversal |
| Phase enc. dir. | A >> P |
| FoV read | 360 mm |
| FoV phase | 75.0 % |
| Slice thickness | 8.0 mm |
| TR | 378.98 ms |
| Multi-slice mode | Sequential |
| Series | Interleaved |
| Concatenations | 1 |

#### Geometry - AutoAlign

| | |
|---|---|
| Slice group | 1 |
| Position | Isocenter |
| Orientation | Transversal |
| Phase enc. dir. | A >> P |
| AutoAlign | --- |
| Initial Position | Isocenter |
| Phase | 0.0 mm |
| Read | 0.0 mm |
| Shift | 0.0 mm |
| Initial Rotation | 0.00 deg |
| Initial Orientation | Transversal |

#### Geometry - Saturation

| | |
|---|---|
| Fat suppr. | None |
| Wrap-up Magn. | None |
| Special sat. | None |

#### Geometry - Navigator

#### System - Miscellaneous

| | |
|---|---|
| Positioning mode | REF |
| Table position | H |
| Table position | 0 mm |
| MSMA | S - C - T |
| Sagittal | R >> L |

#### System - Miscellaneous

| | |
|---|---|
| Coronal | A >> P |
| Transversal | F >> H |
| Coil Combine Mode | Adaptive Combine |
| Save uncombined | Off |
| Matrix Optimization | Off |
| Coil Focus | Flat |
| AutoAlign | --- |
| Coil Select Mode | Default |

#### System - Adjustments

| | |
|---|---|
| B0 Shim mode | Cardiac |
| Adjust with body coil | On |
| Confirm freq. adjustment | Off |
| Assume Dominant Fat | Off |
| Assume Silicone | Off |
| Adjustment Tolerance | Auto |

#### System - Adjust Volume

| | |
|---|---|
| ! Position | Isocenter |
| ! Orientation | Transversal |
| ! Rotation | 0.00 deg |
| ! A >> P | 150 mm |
| ! R >> L | 150 mm |
| ! F >> H | 150 mm |
| Reset | Off |

#### System - Tx/Rx

| | |
|---|---|
| Frequency 1H | 63.678323 MHz |
| Correction factor | 1 |
| Gain | High |
| Img. Scale Cor. | 1.000 |
| Reset | Off |
| ? Ref. amplitude 1H | 0.000 V |

#### Physio - Signal1

| | |
|---|---|
| 1st Signal/Mode | ECG/Trigger |
| Average cycle | No Signal ms |
| Average cycle | No Signal ms |
| Captured cycle | -not set- |
| Acquisition window | 591 ms |
| Trigger pulse | 1 |
| Trigger delay | 212 ms |
| TR | 378.98 ms |
| Concatenations | 1 |
| Segments | 84 |
| Phases | 1 |
| Adaptive Triggering | Off |

#### Physio - Cardiac

| | |
|---|---|
| Tagging | None |
| Magn. preparation | Non-sel. IR |
| TI | 260 ms |
| Fat suppr. | None |
| Dark blood | Off |
| FoV read | 360 mm |
| FoV phase | 75.0 % |
| Phase resolution | 100 % |
| Cine | Off |
| Trajectory | Cartesian |
| Dummy heartbeats | 0 |
| Motion Correction | None |

#### Physio - PACE

| | |
|---|---|
| Resp. control | Off |

#### Physio - PACE

| | |
|---|---|
| Concatenations | 1 |

#### Sequence - Part 1

| | |
|---|---|
| Introduction | Off |
| Dimension | 2D |
| Reordering | Linear |
| Asymmetric echo | Weak |
| Contrasts | 1 |
| Optimization | Min. TE TR |
| Multi-slice mode | Sequential |
| Sequence type | Trufi |
| Bandwidth | 898 Hz/Px |

#### Sequence - Part 2

| | |
|---|---|
| Define | Shots |
| Shots per slice | 1 |
| Segments | 84 |
| Trufi delta freq. | 0 Hz |
| RF pulse type | Fast |
| Gradient mode | Fast |
| Excitation | Slice-sel. |
| Flip angle mode | Constant |
| Cine | Off |

#### Sequence - Assistant

| | |
|---|---|
| Mode | Off |
| Allowed delay | 0 s |

| \\USER\Cardiac Research Protocols\IV IRON\IV IRON4\KLS_ShMOLLI_192i_d11_nFilt_FOV460 |
|---|
| TA: 5.3 s PM: REF Voxel size: 1.2×1.2×8.0 mmPAT: 2 Rel. SNR: 1.00 : tfi |

#### Properties

| | |
|---|---|
| Prio recon | Off |
| Load images to viewer | On |
| Inline movie | Off |
| Auto store images | On |
| Load images to stamp segments | Off |
| Load images to graphic segments | On |
| Auto open inline display | Off |
| Auto close inline display | Off |
| Start measurement without further preparation | Off |
| Wait for user to start | Off |
| Start measurements | Single measurement |

#### Routine

| | |
|---|---|
| Slice group | 1 |
| Slices | 1 |
| Dist. factor | 25 % |
| Position | Isocenter |
| Orientation | Coronal |
| Phase enc. dir. | R >> L |
| AutoAlign | --- |
| Phase oversampling | 0 % |
| FoV read | 460 mm |
| FoV phase | 75.0 % |
| Slice thickness | 8.0 mm |
| TR | 375.62 ms |
| TE | 1.07 ms |
| Averages | 1 |
| Concatenations | 1 |
| Filter | Raw filter, Distortion Corr.(2D) |
| Coil elements | BO2;SP2,3 |

#### Contrast - Common

| | |
|---|---|
| TR | 375.62 ms |
| TE | 1.07 ms |
| Magn. preparation | Non-sel. IR |
| TI | 260 ms |
| Flip angle | 35 deg |
| Fat suppr. | None |
| Wrap-up Magn. | None |

#### Contrast - Dynamic

| | |
|---|---|
| Averages | 1 |
| Averaging mode | Short term |
| Reconstruction | Magn./Phase |
| Measurements | 1 |
| Multiple series | Off |

#### Resolution - Common

| | |
|---|---|
| FoV read | 460 mm |
| FoV phase | 75.0 % |
| Slice thickness | 8.0 mm |
| Base resolution | 192 |
| Phase resolution | 100 % |
| Phase partial Fourier | 6/8 |
| Trajectory | Cartesian |
| Interpolation | On |

#### Resolution - iPAT

| | |
|---|---|
| PAT mode | GRAPPA |
| Accel. factor PE | 2 |
| Ref. lines PE | 24 |
| Reference scan mode | Integrated |

#### Resolution - Filter Image

| | |
|---|---|
| Image Filter | Off |
| Distortion Corr. | On |
| Mode | 2D |
| Unfiltered images | Off |
| Prescan Normalize | Off |
| Normalize | Off |
| B1 filter | Off |

#### Resolution - Filter Rawdata

| | |
|---|---|
| Raw filter | On |
| Elliptical filter | Off |
| POCS | Off |

#### Geometry - Common

| | |
|---|---|
| Slice group | 1 |
| Slices | 1 |
| Dist. factor | 25 % |
| Position | Isocenter |
| Orientation | Coronal |
| Phase enc. dir. | R >> L |
| FoV read | 460 mm |
| FoV phase | 75.0 % |
| Slice thickness | 8.0 mm |
| TR | 375.62 ms |
| Multi-slice mode | Sequential |
| Series | Interleaved |
| Concatenations | 1 |

#### Geometry - AutoAlign

| | |
|---|---|
| Slice group | 1 |
| Position | Isocenter |
| Orientation | Coronal |
| Phase enc. dir. | R >> L |
| AutoAlign | --- |
| Initial Position | Isocenter |
| Phase | 0.0 mm |
| Read | 0.0 mm |
| Shift | 0.0 mm |
| Initial Rotation | 0.00 deg |
| Initial Orientation | Coronal |

#### Geometry - Saturation

| | |
|---|---|
| Fat suppr. | None |
| Wrap-up Magn. | None |
| Special sat. | None |

#### Geometry - Navigator

#### System - Miscellaneous

| | |
|---|---|
| Positioning mode | REF |
| Table position | H |
| Table position | 0 mm |
| MSMA | S - C - T |
| Sagittal | R >> L |

#### System - Miscellaneous

| | |
|---|---|
| Coronal | A >> P |
| Transversal | F >> H |
| Coil Combine Mode | Adaptive Combine |
| Save uncombined | Off |
| Matrix Optimization | Off |
| Coil Focus | Flat |
| AutoAlign | --- |
| Coil Select Mode | Default |

#### System - Adjustments

| | |
|---|---|
| B0 Shim mode | Cardiac |
| Adjust with body coil | On |
| Confirm freq. adjustment | Off |
| Assume Dominant Fat | Off |
| Assume Silicone | Off |
| Adjustment Tolerance | Auto |

#### System - Adjust Volume

| | |
|---|---|
| ! Position | Isocenter |
| ! Orientation | Transversal |
| ! Rotation | 0.00 deg |
| ! A >> P | 150 mm |
| ! R >> L | 150 mm |
| ! F >> H | 150 mm |
| Reset | Off |

#### System - Tx/Rx

| | |
|---|---|
| Frequency 1H | 63.678323 MHz |
| Correction factor | 1 |
| Gain | High |
| Img. Scale Cor. | 1.000 |
| Reset | Off |
| ? Ref. amplitude 1H | 0.000 V |

#### Physio - Signal1

| | |
|---|---|
| 1st Signal/Mode | ECG/Trigger |
| Average cycle | No Signal ms |
| Average cycle | No Signal ms |
| Captured cycle | -not set- |
| Acquisition window | 591 ms |
| Trigger pulse | 1 |
| Trigger delay | 215 ms |
| TR | 375.62 ms |
| Concatenations | 1 |
| Segments | 84 |
| Phases | 1 |
| Adaptive Triggering | Off |

#### Physio - Cardiac

| | |
|---|---|
| Tagging | None |
| Magn. preparation | Non-sel. IR |
| TI | 260 ms |
| Fat suppr. | None |
| Dark blood | Off |
| FoV read | 460 mm |
| FoV phase | 75.0 % |
| Phase resolution | 100 % |
| Cine | Off |
| Trajectory | Cartesian |
| Dummy heartbeats | 0 |
| Motion Correction | None |

#### Physio - PACE

| | |
|---|---|
| Resp. control | Off |
| Concatenations | 1 |

#### Sequence - Part 1

| | |
|---|---|
| Introduction | Off |
| Dimension | 2D |
| Reordering | Linear |
| Asymmetric echo | Weak |
| Contrasts | 1 |
| Optimization | Min. TE TR |
| Multi-slice mode | Sequential |
| Sequence type | Trufi |
| Bandwidth | 898 Hz/Px |

#### Sequence - Part 2

| | |
|---|---|
| Define | Shots |
| Shots per slice | 1 |
| Segments | 84 |
| Trufi delta freq. | 0 Hz |
| RF pulse type | Fast |
| Gradient mode | Fast |
| Excitation | Slice-sel. |
| Flip angle mode | Constant |
| Cine | Off |

#### Sequence - Assistant

| | |
|---|---|
| Mode | Off |
| Allowed delay | 0 s |

| \\USER\Cardiac Research Protocols\IV IRON\IV IRON4\VLST2Map_TrueFISP |
|---|
| TA: 5.3 s PM: REF Voxel size: 1.9×1.9×8.0 mmPAT: 2 Rel. SNR: 1.00 : tfi |

#### Properties

| | |
|---|---|
| Prio recon | Off |
| Load images to viewer | On |
| Inline movie | Off |
| Auto store images | On |
| Load images to stamp segments | Off |
| Load images to graphic segments | On |
| Auto open inline display | Off |
| Auto close inline display | Off |
| Start measurement without further preparation | Off |
| Wait for user to start | On |
| Start measurements | Single measurement |

#### Routine

| | |
|---|---|
| Slice group | 1 |
| Slices | 1 |
| Dist. factor | 20 % |
| Position | Isocenter |
| Orientation | Transversal |
| Phase enc. dir. | A >> P |
| AutoAlign | --- |
| Phase oversampling | 0 % |
| FoV read | 360 mm |
| FoV phase | 75.0 % |
| Slice thickness | 8.0 mm |
| TR | 185.80 ms |
| TE | 1.07 ms |
| Averages | 1 |
| Concatenations | 1 |
| Filter | Distortion Corr.(2D) |
| Coil elements | BO1-3;SP1-3 |

#### Contrast - Common

| | |
|---|---|
| TR | 185.80 ms |
| TE | 1.07 ms |
| Magn. preparation | T2 prep. adiab. |
| T2 prep. duration 1 | 0 ms |
| T2 prep. duration 2 | 25 ms |
| T2 prep. duration 3 | 55 ms |
| Flip angle | 70 deg |
| Fat suppr. | None |
| Wrap-up Magn. | None |

#### Contrast - Dynamic

| | |
|---|---|
| Averages | 1 |
| Averaging mode | Short term |
| Reconstruction | Magnitude |
| Measurements | 1 |
| Multiple series | Off |

#### Resolution - Common

| | |
|---|---|
| FoV read | 360 mm |
| FoV phase | 75.0 % |
| Slice thickness | 8.0 mm |
| Base resolution | 192 |
| Phase resolution | 75 % |
| Phase partial Fourier | 6/8 |
| Trajectory | Cartesian |
| Interpolation | Off |

#### Resolution - iPAT

| | |
|---|---|
| PAT mode | GRAPPA |
| Accel. factor PE | 2 |
| Ref. lines PE | 36 |
| Reference scan mode | GRE/separate |

#### Resolution - Filter Image

| | |
|---|---|
| Image Filter | Off |
| Distortion Corr. | On |
| Mode | 2D |
| Unfiltered images | Off |
| Prescan Normalize | Off |
| Normalize | Off |
| B1 filter | Off |

#### Resolution - Filter Rawdata

| | |
|---|---|
| Raw filter | Off |
| Elliptical filter | Off |
| POCS | Off |

#### Geometry - Common

| | |
|---|---|
| Slice group | 1 |
| Slices | 1 |
| Dist. factor | 20 % |
| Position | Isocenter |
| Orientation | Transversal |
| Phase enc. dir. | A >> P |
| FoV read | 360 mm |
| FoV phase | 75.0 % |
| Slice thickness | 8.0 mm |
| TR | 185.80 ms |
| Multi-slice mode | Sequential |
| Series | Base To Apex |
| Concatenations | 1 |

#### Geometry - AutoAlign

| | |
|---|---|
| Slice group | 1 |
| Position | Isocenter |
| Orientation | Transversal |
| Phase enc. dir. | A >> P |
| AutoAlign | --- |
| Initial Position | Isocenter |
| Phase | 0.0 mm |
| Read | 0.0 mm |
| Shift | 0.0 mm |
| Initial Rotation | 0.00 deg |
| Initial Orientation | Transversal |

#### Geometry - Saturation

| | |
|---|---|
| Fat suppr. | None |
| Wrap-up Magn. | None |
| Special sat. | None |

#### Geometry - Navigator

#### System - Miscellaneous

| | |
|---|---|
| Positioning mode | REF |
| Table position | H |
| Table position | 0 mm |
| MSMA | S - C - T |
| Sagittal | R >> L |

#### System - Miscellaneous

| | |
|---|---|
| Coronal | A >> P |
| Transversal | F >> H |
| Coil Combine Mode | Adaptive Combine |
| Save uncombined | Off |
| Matrix Optimization | Off |
| Coil Focus | Flat |
| AutoAlign | --- |
| Coil Select Mode | Default |

#### System - Adjustments

| | |
|---|---|
| B0 Shim mode | Tune up |
| Adjust with body coil | Off |
| Confirm freq. adjustment | Off |
| Assume Dominant Fat | Off |
| Assume Silicone | Off |
| Adjustment Tolerance | Auto |

#### System - Adjust Volume

| | |
|---|---|
| Position | Isocenter |
| Orientation | Transversal |
| Rotation | 0.00 deg |
| A >> P | 263 mm |
| R >> L | 350 mm |
| F >> H | 350 mm |
| Reset | Off |

#### System - Tx/Rx

| | |
|---|---|
| Frequency 1H | 63.678323 MHz |
| Correction factor | 1 |
| Gain | High |
| Img. Scale Cor. | 1.000 |
| Reset | Off |
| ? Ref. amplitude 1H | 0.000 V |

#### Physio - Signal1

| | |
|---|---|
| 1st Signal/Mode | ECG/Trigger |
| Average cycle | No Signal ms |
| Average cycle | No Signal ms |
| Captured cycle | -not set- |
| Acquisition window | 591 ms |
| Trigger pulse | 1 |
| Trigger delay | 405 ms |
| TR | 185.80 ms |
| Concatenations | 1 |
| Segments | 54 |
| Phases | 1 |
| Adaptive Triggering | Off |

#### Physio - Cardiac

| | |
|---|---|
| Tagging | None |
| Magn. preparation | T2 prep. adiab. |
| T2 prep. duration 1 | 0 ms |
| T2 prep. duration 2 | 25 ms |
| T2 prep. duration 3 | 55 ms |
| Fat suppr. | None |
| Dark blood | Off |
| FoV read | 360 mm |
| FoV phase | 75.0 % |
| Phase resolution | 75 % |
| Cine | Off |
| Trajectory | Cartesian |
| Dummy heartbeats | 0 |
| Motion Correction | Standard |

#### Physio - PACE

| | |
|---|---|
| Resp. control | Breath-hold |
| Concatenations | 1 |

#### Sequence - Part 1

| | |
|---|---|
| Introduction | Off |
| Dimension | 2D |
| Reordering | Linear |
| Asymmetric echo | Weak |
| Contrasts | 1 |
| Optimization | Min. TE TR |
| Multi-slice mode | Sequential |
| Sequence type | Trufi |
| Bandwidth | 1184 Hz/Px |

#### Sequence - Part 2

| | |
|---|---|
| Define | Shots |
| Shots per slice | 1 |
| Segments | 54 |
| Trufi delta freq. | 0 Hz |
| RF pulse type | Fast |
| Gradient mode | Fast |
| Excitation | Slice-sel. |
| Flip angle mode | Constant |
| Cine | Off |

#### Sequence - Assistant

| | |
|---|---|
| Mode | Off |
| Allowed delay | 0 s |

| \\USER\Cardiac Research Protocols\IV IRON\IV IRON4\VLSShMOLLI_C2P_m |
|---|
| TA: 5.3 s PM: FIX Voxel size: 0.9×0.9×8.0 mmPAT: 2 Rel. SNR: 1.00 : tfi |

#### Properties

| | |
|---|---|
| Prio recon | Off |
| Load images to viewer | On |
| Inline movie | Off |
| Auto store images | On |
| Load images to stamp segments | Off |
| Load images to graphic segments | On |
| Auto open inline display | Off |
| Auto close inline display | Off |
| Start measurement without further preparation | On |
| Wait for user to start | Off |
| Start measurements | Single measurement |

#### Routine

| | |
|---|---|
| Slice group | 1 |
| Slices | 1 |
| Dist. factor | 25 % |
| Position | Isocenter |
| Orientation | Transversal |
| Phase enc. dir. | A >> P |
| AutoAlign | --- |
| Phase oversampling | 0 % |
| FoV read | 360 mm |
| FoV phase | 75.0 % |
| Slice thickness | 8.0 mm |
| TR | 378.98 ms |
| TE | 1.07 ms |
| Averages | 1 |
| Concatenations | 1 |
| Filter | Raw filter, Distortion Corr.(2D) |
| Coil elements | BO1-3;SP1-3 |

#### Contrast - Common

| | |
|---|---|
| TR | 378.98 ms |
| TE | 1.07 ms |
| Magn. preparation | Non-sel. IR |
| TI | 260 ms |
| Flip angle | 35 deg |
| Fat suppr. | None |
| Wrap-up Magn. | None |

#### Contrast - Dynamic

| | |
|---|---|
| Averages | 1 |
| Averaging mode | Short term |
| Reconstruction | Magn./Phase |
| Measurements | 1 |
| Multiple series | Off |

#### Resolution - Common

| | |
|---|---|
| FoV read | 360 mm |
| FoV phase | 75.0 % |
| Slice thickness | 8.0 mm |
| Base resolution | 192 |
| Phase resolution | 100 % |
| Phase partial Fourier | 6/8 |
| Trajectory | Cartesian |
| Interpolation | On |

#### Resolution - iPAT

| | |
|---|---|
| PAT mode | GRAPPA |
| Accel. factor PE | 2 |
| Ref. lines PE | 24 |
| Reference scan mode | Integrated |

#### Resolution - Filter Image

| | |
|---|---|
| Image Filter | Off |
| Distortion Corr. | On |
| Mode | 2D |
| Unfiltered images | Off |
| Prescan Normalize | Off |
| Normalize | Off |
| B1 filter | Off |

#### Resolution - Filter Rawdata

| | |
|---|---|
| Raw filter | On |
| Elliptical filter | Off |
| POCS | Off |

#### Geometry - Common

| | |
|---|---|
| Slice group | 1 |
| Slices | 1 |
| Dist. factor | 25 % |
| Position | Isocenter |
| Orientation | Transversal |
| Phase enc. dir. | A >> P |
| FoV read | 360 mm |
| FoV phase | 75.0 % |
| Slice thickness | 8.0 mm |
| TR | 378.98 ms |
| Multi-slice mode | Sequential |
| Series | Interleaved |
| Concatenations | 1 |

#### Geometry - AutoAlign

| | |
|---|---|
| Slice group | 1 |
| Position | Isocenter |
| Orientation | Transversal |
| Phase enc. dir. | A >> P |
| AutoAlign | --- |
| Initial Position | Isocenter |
| Phase | 0.0 mm |
| Read | 0.0 mm |
| Shift | 0.0 mm |
| Initial Rotation | 0.00 deg |
| Initial Orientation | Transversal |

#### Geometry - Saturation

| | |
|---|---|
| Fat suppr. | None |
| Wrap-up Magn. | None |
| Special sat. | None |

#### Geometry - Navigator

#### System - Miscellaneous

| | |
|---|---|
| Positioning mode | FIX |
| Table position | H |
| Table position | 0 mm |
| MSMA | S - C - T |
| Sagittal | R >> L |

#### System - Miscellaneous

| | |
|---|---|
| Coronal | A >> P |
| Transversal | F >> H |
| Coil Combine Mode | Adaptive Combine |
| Save uncombined | Off |
| Matrix Optimization | Off |
| Coil Focus | Flat |
| AutoAlign | --- |
| Coil Select Mode | Default |

#### System - Adjustments

| | |
|---|---|
| B0 Shim mode | Tune up |
| Adjust with body coil | On |
| Confirm freq. adjustment | Off |
| Assume Dominant Fat | Off |
| Assume Silicone | Off |
| Adjustment Tolerance | Auto |

#### System - Adjust Volume

| | |
|---|---|
| ! Position | Isocenter |
| ! Orientation | Transversal |
| ! Rotation | 0.00 deg |
| ! A >> P | 263 mm |
| ! R >> L | 350 mm |
| ! F >> H | 350 mm |
| Reset | Off |

#### System - Tx/Rx

| | |
|---|---|
| Frequency 1H | 63.678323 MHz |
| Correction factor | 1 |
| Gain | High |
| Img. Scale Cor. | 1.000 |
| Reset | Off |
| ? Ref. amplitude 1H | 0.000 V |

#### Physio - Signal1

| | |
|---|---|
| 1st Signal/Mode | ECG/Trigger |
| Average cycle | No Signal ms |
| Average cycle | No Signal ms |
| Captured cycle | -not set- |
| Acquisition window | 591 ms |
| Trigger pulse | 1 |
| Trigger delay | 212 ms |
| TR | 378.98 ms |
| Concatenations | 1 |
| Segments | 84 |
| Phases | 1 |
| Adaptive Triggering | Off |

#### Physio - Cardiac

| | |
|---|---|
| Tagging | None |
| Magn. preparation | Non-sel. IR |
| TI | 260 ms |
| Fat suppr. | None |
| Dark blood | Off |
| FoV read | 360 mm |
| FoV phase | 75.0 % |
| Phase resolution | 100 % |
| Cine | Off |
| Trajectory | Cartesian |
| Dummy heartbeats | 0 |
| Motion Correction | Standard |

#### Physio - PACE

| | |
|---|---|
| Resp. control | Off |

#### Physio - PACE

| | |
|---|---|
| Concatenations | 1 |

#### Sequence - Part 1

| | |
|---|---|
| Introduction | Off |
| Dimension | 2D |
| Reordering | Linear |
| Asymmetric echo | Weak |
| Contrasts | 1 |
| Optimization | Min. TE TR |
| Multi-slice mode | Sequential |
| Sequence type | Trufi |
| Bandwidth | 898 Hz/Px |

#### Sequence - Part 2

| | |
|---|---|
| Define | Shots |
| Shots per slice | 1 |
| Segments | 84 |
| Trufi delta freq. | 0 Hz |
| RF pulse type | Fast |
| Gradient mode | Fast |
| Excitation | Slice-sel. |
| Flip angle mode | Constant |
| Cine | Off |

#### Sequence - Assistant

| | |
|---|---|
| Mode | Off |
| Allowed delay | 0 s |

| \\USER\Cardiac Research Protocols\IV IRON\IV IRON4\VLST2StarMap_8echo_heart |
|---|
| TA: 9.4 s PM: FIX Voxel size: 1.4×1.4×8.0 mmPAT: 2 Rel. SNR: 1.00 : fl_r |

#### Properties

| | |
|---|---|
| Prio recon | Off |
| Load images to viewer | On |
| Inline movie | Off |
| Auto store images | On |
| Load images to stamp segments | Off |
| Load images to graphic segments | On |
| Auto open inline display | Off |
| Auto close inline display | Off |
| Start measurement without further preparation | On |
| Wait for user to start | Off |
| Start measurements | Single measurement |

#### Routine

| | |
|---|---|
| Slice group | 1 |
| Slices | 1 |
| Dist. factor | 20 % |
| Position | Isocenter |
| Orientation | Transversal |
| Phase enc. dir. | A >> P |
| AutoAlign | --- |
| Phase oversampling | 0 % |
| FoV read | 360 mm |
| FoV phase | 75.0 % |
| Slice thickness | 8.0 mm |
| TR | 590.00 ms |
| TE 1 | 2.11 ms |
| TE 2 | 4.25 ms |
| TE 3 | 6.39 ms |
| TE 4 | 8.53 ms |
| TE 5 | 10.67 ms |
| TE 6 | 12.81 ms |
| TE 7 | 14.95 ms |
| TE 8 | 17.09 ms |
| Averages | 1 |
| Concatenations | 1 |
| Filter | Distortion Corr.(2D) |
| Coil elements | BO1-3;SP1-3 |

#### Contrast - Common

| | |
|---|---|
| TR | 590.00 ms |
| TE 1 | 2.11 ms |
| TE 2 | 4.25 ms |
| TE 3 | 6.39 ms |
| TE 4 | 8.53 ms |
| TE 5 | 10.67 ms |
| TE 6 | 12.81 ms |
| TE 7 | 14.95 ms |
| TE 8 | 17.09 ms |
| Flip angle | 20 deg |
| Fat suppr. | Fat sat. |
| Wrap-up Magn. | None |

#### Contrast - Dynamic

| | |
|---|---|
| Averages | 1 |
| Averaging mode | Short term |
| Reconstruction | Magnitude |
| Measurements | 1 |
| Multiple series | Off |

#### Resolution - Common

| | |
|---|---|
| FoV read | 360 mm |
| FoV phase | 75.0 % |
| Slice thickness | 8.0 mm |
| Base resolution | 256 |
| Phase resolution | 60 % |
| Phase partial Fourier | Off |
| Trajectory | Cartesian |
| Interpolation | Off |

#### Resolution - iPAT

| | |
|---|---|
| PAT mode | GRAPPA |
| Accel. factor PE | 2 |
| Ref. lines PE | 24 |
| Reference scan mode | Integrated |

#### Resolution - Filter Image

| | |
|---|---|
| Image Filter | Off |
| Distortion Corr. | On |
| Mode | 2D |
| Unfiltered images | Off |
| Prescan Normalize | Off |
| Normalize | Off |
| B1 filter | Off |

#### Resolution - Filter Rawdata

| | |
|---|---|
| Raw filter | Off |
| Elliptical filter | Off |
| POCS | Off |

#### Geometry - Common

| | |
|---|---|
| Slice group | 1 |
| Slices | 1 |
| Dist. factor | 20 % |
| Position | Isocenter |
| Orientation | Transversal |
| Phase enc. dir. | A >> P |
| FoV read | 360 mm |
| FoV phase | 75.0 % |
| Slice thickness | 8.0 mm |
| TR | 590.00 ms |
| Multi-slice mode | Sequential |
| Series | Base To Apex |
| Concatenations | 1 |

#### Geometry - AutoAlign

| | |
|---|---|
| Slice group | 1 |
| Position | Isocenter |
| Orientation | Transversal |
| Phase enc. dir. | A >> P |
| AutoAlign | --- |
| Initial Position | Isocenter |
| Phase | 0.0 mm |
| Read | 0.0 mm |
| Shift | 0.0 mm |
| Initial Rotation | 0.00 deg |
| Initial Orientation | Transversal |

#### Geometry - Saturation

| | |
|---|---|
| Fat suppr. | Fat sat. |
| Wrap-up Magn. | None |

##### Geometry - Saturation

| | |
|---|---|
| Special sat. | None |

##### Geometry - Navigator

##### System - Miscellaneous

| | |
|---|---|
| Positioning mode | FIX |
| Table position | H |
| Table position | 0 mm |
| MSMA | S - C - T |
| Sagittal | R >> L |
| Coronal | A >> P |
| Transversal | F >> H |
| Coil Combine Mode | Adaptive Combine |
| Save uncombined | Off |
| Matrix Optimization | Off |
| Coil Focus | Flat |
| AutoAlign | --- |
| Coil Select Mode | Default |

##### System - Adjustments

| | |
|---|---|
| B0 Shim mode | Tune up |
| Adjust with body coil | Off |
| Confirm freq. adjustment | Off |
| Assume Dominant Fat | Off |
| Assume Silicone | Off |
| Adjustment Tolerance | Auto |

##### System - Adjust Volume

| | |
|---|---|
| ! Position | Isocenter |
| ! Orientation | Transversal |
| ! Rotation | 0.00 deg |
| ! A >> P | 263 mm |
| ! R >> L | 350 mm |
| ! F >> H | 350 mm |
| Reset | Off |

##### System - Tx/Rx

| | |
|---|---|
| Frequency 1H | 63.678323 MHz |
| Correction factor | 1 |
| Gain | High |
| Img. Scale Cor. | 1.000 |
| Reset | Off |
| ? Ref. amplitude 1H | 0.000 V |

##### Physio - Signal1

| | |
|---|---|
| 1st Signal/Mode | ECG/Trigger |
| Average cycle | No Signal ms |
| Average cycle | No Signal ms |
| Captured cycle | -not set- |
| Acquisition window | 590 ms |
| Trigger pulse | 2 |
| Trigger delay | 0 ms |
| TR | 590.00 ms |
| Concatenations | 1 |
| Segments | 9 |
| Phases | 1 |
| Adaptive Triggering | Off |

##### Physio - Cardiac

| | |
|---|---|
| Tagging | None |
| Fat suppr. | Fat sat. |
| Dark blood | On |
| Dark blood thickness | 200 % |

##### Physio - Cardiac

| | |
|---|---|
| FoV read | 360 mm |
| FoV phase | 75.0 % |
| Phase resolution | 60 % |
| Cine | Off |
| Trajectory | Cartesian |
| Dummy heartbeats | 0 |

##### Physio - PACE

| | |
|---|---|
| Resp. control | Breath-hold |
| Concatenations | 1 |

##### Inline - Common

| | |
|---|---|
| Subtract | Off |
| Measurements | 1 |
| StdDev | Off |
| Save original images | On |

##### Inline - Cardiac

| | |
|---|---|
| Inline Evaluation | T2* map |
| Contrasts | 8 |
| TE 1 | 2.11 ms |
| TE 2 | 4.25 ms |
| TE 3 | 6.39 ms |
| TE 4 | 8.53 ms |
| TE 5 | 10.67 ms |
| TE 6 | 12.81 ms |
| TE 7 | 14.95 ms |
| TE 8 | 17.09 ms |
| TR | 590.00 ms |
| Save original images | On |

##### Inline - MIP

| | |
|---|---|
| MIP-Sag | Off |
| MIP-Cor | Off |
| MIP-Tra | Off |
| MIP-Time | Off |
| Save original images | On |

##### Inline - Composing

| | |
|---|---|
| Inline Composing | Off |
| Distortion Corr. | On |
| Mode | 2D |
| Unfiltered images | Off |

##### Sequence - Part 1

| | |
|---|---|
| Introduction | Off |
| Dimension | 2D |
| Reordering | Linear |
| Asymmetric echo | Weak |
| Contrasts | 8 |
| Flow comp. 1 | Yes |
| Readout mode | Monopolar |
| Optimization | Min. TE |
| Multi-slice mode | Sequential |
| Echo spacing | 19.1 ms |
| Sequence type | Gre |
| Bandwidth 1 | 814 Hz/Px |
| Bandwidth 2 | 814 Hz/Px |
| Bandwidth 3 | 814 Hz/Px |
| Bandwidth 4 | 814 Hz/Px |
| Bandwidth 5 | 814 Hz/Px |
| Bandwidth 6 | 814 Hz/Px |
| Bandwidth 7 | 814 Hz/Px |
| Bandwidth 8 | 814 Hz/Px |

#### Sequence - Part 2

| | |
|---|---|
| Define | Segments |
| Segments | 9 |
| RF pulse type | Fast |
| Gradient mode | Fast |
| Excitation | Slice-sel. |
| Flip angle mode | Constant |
| RF spoiling | On |
| Phase Enc. Rewinder | On |
| Cine | Off |

#### Sequence - Assistant

| | |
|---|---|
| Mode | Off |
| Allowed delay | 0 s |

| \\USER\Cardiac Research Protocols\IV IRON\IV IRON4\VLSFB_MOCO_gt_T2star_DB_8e_128 FS_HiFE |
|---|
| TA: 9.4 s PM: FIX Voxel size: 2.8×2.8×8.0 mmPAT: 4 Rel. SNR: 1.00 : tfl |

#### Properties

| | |
|---|---|
| Prio recon | Off |
| Load images to viewer | On |
| Inline movie | Off |
| Auto store images | On |
| Load images to stamp segments | Off |
| Load images to graphic segments | On |
| Auto open inline display | Off |
| Auto close inline display | Off |
| Start measurement without further preparation | On |
| Wait for user to start | Off |
| Start measurements | Single measurement |

#### Routine

| | |
|---|---|
| Slice group | 1 |
| Slices | 1 |
| Dist. factor | 20 % |
| Position | Isocenter |
| Orientation | Transversal |
| Phase enc. dir. | A >> P |
| AutoAlign | --- |
| Phase oversampling | 0 % |
| FoV read | 360 mm |
| FoV phase | 75.0 % |
| Slice thickness | 8.0 mm |
| TR | 590.00 ms |
| TE 1 | 1.07 ms |
| TE 2 | 2.58 ms |
| TE 3 | 4.09 ms |
| TE 4 | 5.6 ms |
| TE 5 | 7.11 ms |
| TE 6 | 8.62 ms |
| TE 7 | 10.13 ms |
| TE 8 | 11.64 ms |
| Averages | 1 |
| Concatenations | 1 |
| Filter | Distortion Corr.(2D) |
| Coil elements | BO1-3;SP1-3 |

#### Contrast - Common

| | |
|---|---|
| TR | 590.00 ms |
| TE 1 | 1.07 ms |
| TE 2 | 2.58 ms |
| TE 3 | 4.09 ms |
| TE 4 | 5.6 ms |
| TE 5 | 7.11 ms |
| TE 6 | 8.62 ms |
| TE 7 | 10.13 ms |
| TE 8 | 11.64 ms |
| Magn. preparation | None |
| Flip angle | 18 deg |
| Fat suppr. | Fat sat. |
| Wrap-up Magn. | None |

#### Contrast - Dynamic

| | |
|---|---|
| Averages | 1 |
| Averaging mode | Long term |
| Reconstruction | Magnitude |
| Measurements | 16 |

#### Contrast - Dynamic

| | |
|---|---|
| Pause after meas. 1 | 0.0 s |
| Pause after meas. 2 | 0.0 s |
| Pause after meas. 3 | 0.0 s |
| Pause after meas. 4 | 0.0 s |
| Pause after meas. 5 | 0.0 s |
| Pause after meas. 6 | 0.0 s |
| Pause after meas. 7 | 0.0 s |
| Pause after meas. 8 | 0.0 s |
| Pause after meas. 9 | 0.0 s |
| Pause after meas. 10 | 0.0 s |
| Pause after meas. 11 | 0.0 s |
| Pause after meas. 12 | 0.0 s |
| Pause after meas. 13 | 0.0 s |
| Pause after meas. 14 | 0.0 s |
| Pause after meas. 15 | 0.0 s |
| Multiple series | Off |

#### Resolution - Common

| | |
|---|---|
| FoV read | 360 mm |
| FoV phase | 75.0 % |
| Slice thickness | 8.0 mm |
| Base resolution | 128 |
| Phase resolution | 100 % |
| Phase partial Fourier | Off |
| Trajectory | Cartesian |
| Interpolation | Off |

#### Resolution - iPAT

| | |
|---|---|
| PAT mode | GRAPPA |
| Accel. factor PE | 4 |
| Reference scan mode | T-PAT |

#### Resolution - Filter Image

| | |
|---|---|
| Image Filter | Off |
| Distortion Corr. | On |
| Mode | 2D |
| Unfiltered images | Off |
| Prescan Normalize | Off |
| Normalize | Off |
| B1 filter | Off |

#### Resolution - Filter Rawdata

| | |
|---|---|
| Raw filter | Off |
| Elliptical filter | Off |
| POCS | Off |

#### Geometry - Common

| | |
|---|---|
| Slice group | 1 |
| Slices | 1 |
| Dist. factor | 20 % |
| Position | Isocenter |
| Orientation | Transversal |
| Phase enc. dir. | A >> P |
| FoV read | 360 mm |
| FoV phase | 75.0 % |
| Slice thickness | 8.0 mm |
| TR | 590.00 ms |
| Multi-slice mode | Single shot |
| Series | Interleaved |

#### Geometry - Common

| | |
|---|---|
| Concatenations | 1 |

#### Geometry - AutoAlign

| | |
|---|---|
| Slice group | 1 |
| Position | Isocenter |
| Orientation | Transversal |
| Phase enc. dir. | A >> P |
| AutoAlign | --- |
| Initial Position | Isocenter |
| Phase | 0.0 mm |
| Read | 0.0 mm |
| Shift | 0.0 mm |
| Initial Rotation | 0.00 deg |
| Initial Orientation | Transversal |

#### Geometry - Saturation

| | |
|---|---|
| Fat suppr. | Fat sat. |
| Wrap-up Magn. | None |
| Special sat. | None |

#### Geometry - Navigator

#### System - Miscellaneous

| | |
|---|---|
| Positioning mode | FIX |
| Table position | H |
| Table position | 0 mm |
| MSMA | S - C - T |
| Sagittal | R >> L |
| Coronal | A >> P |
| Transversal | F >> H |
| Coil Combine Mode | Adaptive Combine |
| Save uncombined | Off |
| Matrix Optimization | Off |
| Coil Focus | Flat |
| AutoAlign | --- |
| Coil Select Mode | Default |

#### System - Adjustments

| | |
|---|---|
| B0 Shim mode | Tune up |
| Adjust with body coil | On |
| Confirm freq. adjustment | Off |
| Assume Dominant Fat | Off |
| Assume Silicone | Off |
| Adjustment Tolerance | Auto |

#### System - Adjust Volume

| | |
|---|---|
| ! Position | Isocenter |
| ! Orientation | Transversal |
| ! Rotation | 0.00 deg |
| ! A >> P | 263 mm |
| ! R >> L | 350 mm |
| ! F >> H | 350 mm |
| Reset | Off |

#### System - Tx/Rx

| | |
|---|---|
| Frequency 1H | 63.678323 MHz |
| Correction factor | 1 |
| Gain | High |
| Img. Scale Cor. | 1.000 |
| Reset | Off |
| ? Ref. amplitude 1H | 0.000 V |

#### Physio - Signal1

| | |
|---|---|
| 1st Signal/Mode | ECG/Trigger |
| Average cycle | No Signal ms |
| Average cycle | No Signal ms |
| Captured cycle | -not set- |
| Acquisition window | 590 ms |
| Trigger pulse | 1 |
| Trigger delay | 0 ms |
| TR | 590.00 ms |
| Concatenations | 1 |
| Segments | 24 |
| Phases | 1 |
| Adaptive Triggering | Off |

#### Physio - Cardiac

| | |
|---|---|
| Tagging | None |
| Magn. preparation | None |
| Fat suppr. | Fat sat. |
| Dark blood | On |
| Dark blood thickness | 300 % |
| FoV read | 360 mm |
| FoV phase | 75.0 % |
| Phase resolution | 100 % |
| Cine | Off |
| Trajectory | Cartesian |
| Dummy heartbeats | 0 |

#### Physio - PACE

| | |
|---|---|
| Resp. control | Off |
| Concatenations | 1 |

#### Inline - Common

| | |
|---|---|
| Subtract | Off |
| Measurements | 16 |
| StdDev | Off |
| Save original images | On |

#### Inline - Cardiac

| | |
|---|---|
| Inline Evaluation | Off |
| Magn. preparation | None |
| Contrasts | 8 |
| TE 1 | 1.07 ms |
| TE 2 | 2.58 ms |
| TE 3 | 4.09 ms |
| TE 4 | 5.6 ms |
| TE 5 | 7.11 ms |
| TE 6 | 8.62 ms |
| TE 7 | 10.13 ms |
| TE 8 | 11.64 ms |
| TR | 590.00 ms |
| Save original images | On |

#### Inline - MIP

| | |
|---|---|
| MIP-Sag | Off |
| MIP-Cor | Off |
| MIP-Tra | Off |
| MIP-Time | Off |
| Save original images | On |

#### Inline - Composing

| | |
|---|---|
| Inline Composing | Off |
| Distortion Corr. | On |
| Mode | 2D |
| Unfiltered images | Off |

#### Sequence - Part 1

| | |
|---|---|
| Introduction | Off |
| Dimension | 2D |
| Reordering | Linear |
| Asymmetric echo | Off |
| Contrasts | 8 |
| Flow comp. 1 | No |
| Readout mode | Monopolar |
| Optimization | Min. TE |
| Multi-slice mode | Single shot |
| Echo spacing | 12.6 ms |
| Sequence type | Gre |
| Bandwidth 1 | 1502 Hz/Px |
| Bandwidth 2 | 1502 Hz/Px |
| Bandwidth 3 | 1502 Hz/Px |
| Bandwidth 4 | 1502 Hz/Px |
| Bandwidth 5 | 1502 Hz/Px |
| Bandwidth 6 | 1502 Hz/Px |
| Bandwidth 7 | 1502 Hz/Px |
| Bandwidth 8 | 1502 Hz/Px |

#### Sequence - Part 2

| | |
|---|---|
| Define | Shots |
| Shots per slice | 1 |
| Segments | 24 |
| RF pulse type | Fast |
| Gradient mode | Fast |
| Excitation | Slice-sel. |
| Flip angle mode | Constant |
| RF spoiling | On |
| Phase Enc. Rewinder | On |
| Cine | Off |

#### Sequence - Special

| | |
|---|---|
| FatWater Separation | On |
| Multi-echo Images | Off |
| In-Opp Phase Images | Off |
| Frequency Map | Off |
| T2* Map | Off |
| Motion Correction | Off |
| MoCo Averaging Mode | Complex MoCo |
| MoCo Images Only? | Off |
| No. of Interleaves | 0 |

#### Sequence - Assistant

| | |
|---|---|
| Mode | Off |
| Allowed delay | 0 s |

**\\USER\Cardiac Research Protocols\IV IRON\IV IRON4\VLSFB_MOCO_gt_T2star_DB_8e_160 FS_LowNFE**

TA: 9.4 s PM: FIX Voxel size: 2.3×2.3×8.0 mmPAT: 4 Rel. SNR: 1.00 : tfl

#### Properties

| | |
|---|---|
| Prio recon | Off |
| Load images to viewer | On |
| Inline movie | Off |
| Auto store images | On |
| Load images to stamp segments | Off |
| Load images to graphic segments | On |
| Auto open inline display | Off |
| Auto close inline display | Off |
| Start measurement without further preparation | On |
| Wait for user to start | Off |
| Start measurements | Single measurement |

#### Routine

| | |
|---|---|
| Slice group | 1 |
| Slices | 1 |
| Dist. factor | 20 % |
| Position | Isocenter |
| Orientation | Transversal |
| Phase enc. dir. | A >> P |
| AutoAlign | --- |
| Phase oversampling | 0 % |
| FoV read | 360 mm |
| FoV phase | 75.0 % |
| Slice thickness | 8.0 mm |
| TR | 590.00 ms |
| TE 1 | 1.22 ms |
| TE 2 | 3.05 ms |
| TE 3 | 4.88 ms |
| TE 4 | 6.71 ms |
| TE 5 | 8.54 ms |
| TE 6 | 10.37 ms |
| TE 7 | 12.2 ms |
| TE 8 | 14.03 ms |
| Averages | 1 |
| Concatenations | 1 |
| Filter | Distortion Corr.(2D) |
| Coil elements | BO1-3;SP1-3 |

#### Contrast - Common

| | |
|---|---|
| TR | 590.00 ms |
| TE 1 | 1.22 ms |
| TE 2 | 3.05 ms |
| TE 3 | 4.88 ms |
| TE 4 | 6.71 ms |
| TE 5 | 8.54 ms |
| TE 6 | 10.37 ms |
| TE 7 | 12.2 ms |
| TE 8 | 14.03 ms |
| Magn. preparation | None |
| Flip angle | 18 deg |
| Fat suppr. | Fat sat. |
| Wrap-up Magn. | None |

#### Contrast - Dynamic

| | |
|---|---|
| Averages | 1 |
| Averaging mode | Long term |
| Reconstruction | Magnitude |
| Measurements | 16 |

#### Contrast - Dynamic

| | |
|---|---|
| Pause after meas. 1 | 0.0 s |
| Pause after meas. 2 | 0.0 s |
| Pause after meas. 3 | 0.0 s |
| Pause after meas. 4 | 0.0 s |
| Pause after meas. 5 | 0.0 s |
| Pause after meas. 6 | 0.0 s |
| Pause after meas. 7 | 0.0 s |
| Pause after meas. 8 | 0.0 s |
| Pause after meas. 9 | 0.0 s |
| Pause after meas. 10 | 0.0 s |
| Pause after meas. 11 | 0.0 s |
| Pause after meas. 12 | 0.0 s |
| Pause after meas. 13 | 0.0 s |
| Pause after meas. 14 | 0.0 s |
| Pause after meas. 15 | 0.0 s |
| Multiple series | Off |

#### Resolution - Common

| | |
|---|---|
| FoV read | 360 mm |
| FoV phase | 75.0 % |
| Slice thickness | 8.0 mm |
| Base resolution | 160 |
| Phase resolution | 87 % |
| Phase partial Fourier | Off |
| Trajectory | Cartesian |
| Interpolation | Off |

#### Resolution - iPAT

| | |
|---|---|
| PAT mode | GRAPPA |
| Accel. factor PE | 4 |
| Reference scan mode | T-PAT |

#### Resolution - Filter Image

| | |
|---|---|
| Image Filter | Off |
| Distortion Corr. | On |
| Mode | 2D |
| Unfiltered images | Off |
| Prescan Normalize | Off |
| Normalize | Off |
| B1 filter | Off |

#### Resolution - Filter Rawdata

| | |
|---|---|
| Raw filter | Off |
| Elliptical filter | Off |
| POCS | Off |

#### Geometry - Common

| | |
|---|---|
| Slice group | 1 |
| Slices | 1 |
| Dist. factor | 20 % |
| Position | Isocenter |
| Orientation | Transversal |
| Phase enc. dir. | A >> P |
| FoV read | 360 mm |
| FoV phase | 75.0 % |
| Slice thickness | 8.0 mm |
| TR | 590.00 ms |
| Multi-slice mode | Single shot |
| Series | Interleaved |

#### Geometry - Common

| | |
|---|---|
| Concatenations | 1 |

#### Geometry - AutoAlign

| | |
|---|---|
| Slice group | 1 |
| Position | Isocenter |
| Orientation | Transversal |
| Phase enc. dir. | A >> P |
| AutoAlign | --- |
| Initial Position | Isocenter |
| Phase | 0.0 mm |
| Read | 0.0 mm |
| Shift | 0.0 mm |
| Initial Rotation | 0.00 deg |
| Initial Orientation | Transversal |

#### Geometry - Saturation

| | |
|---|---|
| Fat suppr. | Fat sat. |
| Wrap-up Magn. | None |
| Special sat. | None |

#### Geometry - Navigator

#### System - Miscellaneous

| | |
|---|---|
| Positioning mode | FIX |
| Table position | H |
| Table position | 0 mm |
| MSMA | S - C - T |
| Sagittal | R >> L |
| Coronal | A >> P |
| Transversal | F >> H |
| Coil Combine Mode | Adaptive Combine |
| Save uncombined | Off |
| Matrix Optimization | Off |
| Coil Focus | Flat |
| AutoAlign | --- |
| Coil Select Mode | Default |

#### System - Adjustments

| | |
|---|---|
| B0 Shim mode | Tune up |
| Adjust with body coil | On |
| Confirm freq. adjustment | Off |
| Assume Dominant Fat | Off |
| Assume Silicone | Off |
| Adjustment Tolerance | Auto |

#### System - Adjust Volume

| | |
|---|---|
| ! Position | Isocenter |
| ! Orientation | Transversal |
| ! Rotation | 0.00 deg |
| ! A >> P | 263 mm |
| ! R >> L | 350 mm |
| ! F >> H | 350 mm |
| Reset | Off |

#### System - Tx/Rx

| | |
|---|---|
| Frequency 1H | 63.678323 MHz |
| Correction factor | 1 |
| Gain | High |
| Img. Scale Cor. | 1.000 |
| Reset | Off |
| ? Ref. amplitude 1H | 0.000 V |

#### Physio - Signal1

| | |
|---|---|
| 1st Signal/Mode | ECG/Trigger |
| Average cycle | No Signal ms |
| Average cycle | No Signal ms |
| Captured cycle | -not set- |
| Acquisition window | 590 ms |
| Trigger pulse | 1 |
| Trigger delay | 0 ms |
| TR | 590.00 ms |
| Concatenations | 1 |
| Segments | 26 |
| Phases | 1 |
| Adaptive Triggering | Off |

#### Physio - Cardiac

| | |
|---|---|
| Tagging | None |
| Magn. preparation | None |
| Fat suppr. | Fat sat. |
| Dark blood | On |
| Dark blood thickness | 300 % |
| FoV read | 360 mm |
| FoV phase | 75.0 % |
| Phase resolution | 87 % |
| Cine | Off |
| Trajectory | Cartesian |
| Dummy heartbeats | 0 |

#### Physio - PACE

| | |
|---|---|
| Resp. control | Off |
| Concatenations | 1 |

#### Inline - Common

| | |
|---|---|
| Subtract | Off |
| Measurements | 16 |
| StdDev | Off |
| Save original images | On |

#### Inline - Cardiac

| | |
|---|---|
| Inline Evaluation | Off |
| Magn. preparation | None |
| Contrasts | 8 |
| TE 1 | 1.22 ms |
| TE 2 | 3.05 ms |
| TE 3 | 4.88 ms |
| TE 4 | 6.71 ms |
| TE 5 | 8.54 ms |
| TE 6 | 10.37 ms |
| TE 7 | 12.2 ms |
| TE 8 | 14.03 ms |
| TR | 590.00 ms |
| Save original images | On |

#### Inline - MIP

| | |
|---|---|
| MIP-Sag | Off |
| MIP-Cor | Off |
| MIP-Tra | Off |
| MIP-Time | Off |
| Save original images | On |

#### Inline - Composing

| | |
|---|---|
| Inline Composing | Off |
| Distortion Corr. | On |
| Mode | 2D |
| Unfiltered images | Off |

#### Sequence - Part 1

| | |
|---|---|
| Introduction | Off |
| Dimension | 2D |
| Reordering | Linear |
| Asymmetric echo | Off |
| Contrasts | 8 |
| Flow comp. 1 | No |
| Readout mode | Monopolar |
| Optimization | Min. TE |
| Multi-slice mode | Single shot |
| Echo spacing | 15.1 ms |
| Sequence type | Gre |
| Bandwidth 1 | 1078 Hz/Px |
| Bandwidth 2 | 1078 Hz/Px |
| Bandwidth 3 | 1078 Hz/Px |
| Bandwidth 4 | 1078 Hz/Px |
| Bandwidth 5 | 1078 Hz/Px |
| Bandwidth 6 | 1078 Hz/Px |
| Bandwidth 7 | 1078 Hz/Px |
| Bandwidth 8 | 1078 Hz/Px |

#### Sequence - Part 2

| | |
|---|---|
| Define | Shots |
| Shots per slice | 1 |
| Segments | 26 |
| RF pulse type | Fast |
| Gradient mode | Fast |
| Excitation | Slice-sel. |
| Flip angle mode | Constant |
| RF spoiling | On |
| Phase Enc. Rewinder | On |
| Cine | Off |

#### Sequence - Special

| | |
|---|---|
| FatWater Separation | On |
| Multi-echo Images | Off |
| In-Opp Phase Images | Off |
| Frequency Map | Off |
| T2* Map | Off |
| Motion Correction | Off |
| MoCo Averaging Mode | Complex MoCo |
| MoCo Images Only? | Off |
| No. of Interleaves | 0 |

#### Sequence - Assistant

| | |
|---|---|
| Mode | Off |
| Allowed delay | 0 s |

| \\USER\Cardiac Research Protocols\IV IRON\IV IRON4\VLSnonBH_T2StarMap_12echo_liver |
|---|
| TA: 0:23 PM: FIX Voxel size: 2.8×2.8×8.0 mmPAT: Off Rel. SNR: 1.00 : fl |

#### Properties

| | |
|---|---|
| Prio recon | Off |
| Load images to viewer | On |
| Inline movie | Off |
| Auto store images | On |
| Load images to stamp segments | Off |
| Load images to graphic segments | On |
| Auto open inline display | Off |
| Auto close inline display | Off |
| Start measurement without further preparation | On |
| Wait for user to start | Off |
| Start measurements | Single measurement |

#### Routine

| | |
|---|---|
| Slice group | 1 |
| Slices | 1 |
| Dist. factor | 20 % |
| Position | Isocenter |
| Orientation | Transversal |
| Phase enc. dir. | A >> P |
| AutoAlign | --- |
| Phase oversampling | 20 % |
| FoV read | 360 mm |
| FoV phase | 75.0 % |
| Slice thickness | 8.0 mm |
| TR | 200.00 ms |
| TE 1 | 0.97 ms |
| TE 2 | 2.29 ms |
| TE 3 | 3.61 ms |
| TE 4 | 4.93 ms |
| TE 5 | 6.25 ms |
| TE 6 | 7.57 ms |
| TE 7 | 8.89 ms |
| TE 8 | 10.21 ms |
| TE 9 | 11.53 ms |
| TE 10 | 12.85 ms |
| TE 11 | 14.17 ms |
| TE 12 | 15.49 ms |
| Averages | 1 |
| Concatenations | 1 |
| Filter | Distortion Corr.(2D) |
| Coil elements | BO1-3;SP1-3 |

#### Contrast - Common

| | |
|---|---|
| TR | 200.00 ms |
| TE 1 | 0.97 ms |
| TE 2 | 2.29 ms |
| TE 3 | 3.61 ms |
| TE 4 | 4.93 ms |
| TE 5 | 6.25 ms |
| TE 6 | 7.57 ms |
| TE 7 | 8.89 ms |
| TE 8 | 10.21 ms |
| TE 9 | 11.53 ms |
| TE 10 | 12.85 ms |
| TE 11 | 14.17 ms |
| TE 12 | 15.49 ms |
| Flip angle | 20 deg |
| Fat suppr. | Fat sat. |
| Wrap-up Magn. | None |

#### Contrast - Dynamic

| | |
|---|---|
| Averages | 1 |
| Averaging mode | Short term |
| Reconstruction | Magnitude |
| Measurements | 1 |
| Multiple series | Each measurement |

#### Resolution - Common

| | |
|---|---|
| FoV read | 360 mm |
| FoV phase | 75.0 % |
| Slice thickness | 8.0 mm |
| Base resolution | 128 |
| Phase resolution | 100 % |
| Phase partial Fourier | Off |
| Trajectory | Cartesian |
| Interpolation | Off |

#### Resolution - iPAT

| | |
|---|---|
| PAT mode | None |

#### Resolution - Filter Image

| | |
|---|---|
| Image Filter | Off |
| Distortion Corr. | On |
| Mode | 2D |
| Unfiltered images | Off |
| Prescan Normalize | Off |
| Normalize | Off |
| B1 filter | Off |

#### Resolution - Filter Rawdata

| | |
|---|---|
| Raw filter | Off |
| Elliptical filter | Off |
| POCS | Off |

#### Geometry - Common

| | |
|---|---|
| Slice group | 1 |
| Slices | 1 |
| Dist. factor | 20 % |
| Position | Isocenter |
| Orientation | Transversal |
| Phase enc. dir. | A >> P |
| FoV read | 360 mm |
| FoV phase | 75.0 % |
| Slice thickness | 8.0 mm |
| TR | 200.00 ms |
| Multi-slice mode | Sequential |
| Series | Ascending |
| Concatenations | 1 |

#### Geometry - AutoAlign

| | |
|---|---|
| Slice group | 1 |
| Position | Isocenter |
| Orientation | Transversal |
| Phase enc. dir. | A >> P |
| AutoAlign | --- |
| Initial Position | Isocenter |
| Phase | 0.0 mm |
| Read | 0.0 mm |
| Shift | 0.0 mm |
| Initial Rotation | 0.00 deg |
| Initial Orientation | Transversal |

#### Geometry - Saturation

| | |
|---|---|
| Fat suppr. | Fat sat. |
| Wrap-up Magn. | None |
| Special sat. | None |

#### Geometry - Navigator

#### System - Miscellaneous

| | |
|---|---|
| Positioning mode | FIX |
| Table position | H |
| Table position | 0 mm |
| MSMA | S - C - T |
| Sagittal | R >> L |
| Coronal | A >> P |
| Transversal | F >> H |
| Coil Combine Mode | Adaptive Combine |
| Save uncombined | Off |
| Matrix Optimization | Off |
| Coil Focus | Flat |
| AutoAlign | --- |
| Coil Select Mode | Default |

#### System - Adjustments

| | |
|---|---|
| B0 Shim mode | Tune up |
| Adjust with body coil | Off |
| Confirm freq. adjustment | Off |
| Assume Dominant Fat | Off |
| Assume Silicone | Off |
| Adjustment Tolerance | Auto |

#### System - Adjust Volume

| | |
|---|---|
| ! Position | Isocenter |
| ! Orientation | Transversal |
| ! Rotation | 0.00 deg |
| ! A >> P | 263 mm |
| ! R >> L | 350 mm |
| ! F >> H | 350 mm |
| Reset | Off |

#### System - Tx/Rx

| | |
|---|---|
| Frequency 1H | 63.678323 MHz |
| Correction factor | 1 |
| Gain | High |
| Img. Scale Cor. | 1.000 |
| Reset | Off |
| ? Ref. amplitude 1H | 0.000 V |

#### Physio - Signal1

| | |
|---|---|
| 1st Signal/Mode | None |
| TR | 200.00 ms |
| Concatenations | 1 |
| Segments | 1 |

#### Physio - Cardiac

| | |
|---|---|
| Tagging | None |
| Fat suppr. | Fat sat. |
| Dark blood | Off |
| FoV read | 360 mm |
| FoV phase | 75.0 % |
| Phase resolution | 100 % |
| Cine | Off |
| Trajectory | Cartesian |
| Dummy heartbeats | 0 |

#### Physio - PACE

| | |
|---|---|
| Resp. control | Off |
| Concatenations | 1 |

#### Inline - Common

| | |
|---|---|
| Subtract | Off |
| Measurements | 1 |
| StdDev | Off |
| Save original images | On |

#### Inline - Cardiac

| | |
|---|---|
| Inline Evaluation | T2* map |
| Contrasts | 12 |
| TE 1 | 0.97 ms |
| TE 2 | 2.29 ms |
| TE 3 | 3.61 ms |
| TE 4 | 4.93 ms |
| TE 5 | 6.25 ms |
| TE 6 | 7.57 ms |
| TE 7 | 8.89 ms |
| TE 8 | 10.21 ms |
| TE 9 | 11.53 ms |
| TE 10 | 12.85 ms |
| TE 11 | 14.17 ms |
| TE 12 | 15.49 ms |
| TR | 200.00 ms |
| Save original images | On |

#### Inline - MIP

| | |
|---|---|
| MIP-Sag | Off |
| MIP-Cor | Off |
| MIP-Tra | Off |
| MIP-Time | Off |
| Save original images | On |

#### Inline - Composing

| | |
|---|---|
| Inline Composing | Off |
| Distortion Corr. | On |
| Mode | 2D |
| Unfiltered images | Off |

#### Sequence - Part 1

| | |
|---|---|
| Introduction | Off |
| Dimension | 2D |
| Reordering | Linear |
| Asymmetric echo | Weak |
| Contrasts | 12 |
| Flow comp. 1 | No |
| Readout mode | Monopolar |
| Optimization | Min. TE |
| Multi-slice mode | Sequential |
| Echo spacing | 16.8 ms |
| Sequence type | Gre |
| Bandwidth 1 | 1953 Hz/Px |
| Bandwidth 2 | 1953 Hz/Px |
| Bandwidth 3 | 1953 Hz/Px |
| Bandwidth 4 | 1953 Hz/Px |
| Bandwidth 5 | 1953 Hz/Px |
| Bandwidth 6 | 1953 Hz/Px |
| Bandwidth 7 | 1953 Hz/Px |
| Bandwidth 8 | 1953 Hz/Px |
| Bandwidth 9 | 1953 Hz/Px |
| Bandwidth 10 | 1953 Hz/Px |
| Bandwidth 11 | 1953 Hz/Px |
| Bandwidth 12 | 1953 Hz/Px |

#### Sequence - Part 2

| | |
|---|---|
| Define | Segments |
| Segments | 1 |
| RF pulse type | Fast |
| Gradient mode | Fast |
| Excitation | Slice-sel. |
| Flip angle mode | Constant |
| RF spoiling | On |
| Phase Enc. Rewinder | On |
| Cine | Off |

#### Sequence - Assistant

| | |
|---|---|
| Mode | Off |
| Allowed delay | 0 s |

| \\USER\Cardiac Research Protocols\IV IRON\IV IRON4\KLST2Map_TrueFISP |
|---|
| TA: 5.3 s PM: REF Voxel size: 2.4×2.4×8.0 mmPAT: 2 Rel. SNR: 1.00 : tfi |

#### Properties

| | |
|---|---|
| Prio recon | Off |
| Load images to viewer | On |
| Inline movie | Off |
| Auto store images | On |
| Load images to stamp segments | Off |
| Load images to graphic segments | On |
| Auto open inline display | Off |
| Auto close inline display | Off |
| Start measurement without further preparation | Off |
| Wait for user to start | On |
| Start measurements | Single measurement |

#### Routine

| | |
|---|---|
| Slice group | 1 |
| Slices | 1 |
| Dist. factor | 20 % |
| Position | Isocenter |
| Orientation | Transversal |
| Phase enc. dir. | A >> P |
| AutoAlign | --- |
| Phase oversampling | 0 % |
| FoV read | 460 mm |
| FoV phase | 75.0 % |
| Slice thickness | 8.0 mm |
| TR | 179.50 ms |
| TE | 1 ms |
| Averages | 1 |
| Concatenations | 1 |
| Filter | Distortion Corr.(2D) |
| Coil elements | BO1-3;SP1-3 |

#### Contrast - Common

| | |
|---|---|
| TR | 179.50 ms |
| TE | 1 ms |
| Magn. preparation | T2 prep. adiab. |
| T2 prep. duration 1 | 0 ms |
| T2 prep. duration 2 | 25 ms |
| T2 prep. duration 3 | 55 ms |
| Flip angle | 70 deg |
| Fat suppr. | None |
| Wrap-up Magn. | None |

#### Contrast - Dynamic

| | |
|---|---|
| Averages | 1 |
| Averaging mode | Short term |
| Reconstruction | Magnitude |
| Measurements | 1 |
| Multiple series | Off |

#### Resolution - Common

| | |
|---|---|
| FoV read | 460 mm |
| FoV phase | 75.0 % |
| Slice thickness | 8.0 mm |
| Base resolution | 192 |
| Phase resolution | 75 % |
| Phase partial Fourier | 6/8 |
| Trajectory | Cartesian |
| Interpolation | Off |

#### Resolution - iPAT

| | |
|---|---|
| PAT mode | GRAPPA |
| Accel. factor PE | 2 |
| Ref. lines PE | 36 |
| Reference scan mode | GRE/separate |

#### Resolution - Filter Image

| | |
|---|---|
| Image Filter | Off |
| Distortion Corr. | On |
| Mode | 2D |
| Unfiltered images | Off |
| Prescan Normalize | Off |
| Normalize | Off |
| B1 filter | Off |

#### Resolution - Filter Rawdata

| | |
|---|---|
| Raw filter | Off |
| Elliptical filter | Off |
| POCS | Off |

#### Geometry - Common

| | |
|---|---|
| Slice group | 1 |
| Slices | 1 |
| Dist. factor | 20 % |
| Position | Isocenter |
| Orientation | Transversal |
| Phase enc. dir. | A >> P |
| FoV read | 460 mm |
| FoV phase | 75.0 % |
| Slice thickness | 8.0 mm |
| TR | 179.50 ms |
| Multi-slice mode | Sequential |
| Series | Base To Apex |
| Concatenations | 1 |

#### Geometry - AutoAlign

| | |
|---|---|
| Slice group | 1 |
| Position | Isocenter |
| Orientation | Transversal |
| Phase enc. dir. | A >> P |
| AutoAlign | --- |
| Initial Position | Isocenter |
| Phase | 0.0 mm |
| Read | 0.0 mm |
| Shift | 0.0 mm |
| Initial Rotation | 0.00 deg |
| Initial Orientation | Transversal |

#### Geometry - Saturation

| | |
|---|---|
| Fat suppr. | None |
| Wrap-up Magn. | None |
| Special sat. | None |

#### Geometry - Navigator

#### System - Miscellaneous

| | |
|---|---|
| Positioning mode | REF |
| Table position | H |
| Table position | 0 mm |
| MSMA | S - C - T |
| Sagittal | R >> L |

#### System - Miscellaneous

| | |
|---|---|
| Coronal | A >> P |
| Transversal | F >> H |
| Coil Combine Mode | Adaptive Combine |
| Save uncombined | Off |
| Matrix Optimization | Off |
| Coil Focus | Flat |
| AutoAlign | --- |
| Coil Select Mode | Default |

#### System - Adjustments

| | |
|---|---|
| B0 Shim mode | Tune up |
| Adjust with body coil | Off |
| Confirm freq. adjustment | Off |
| Assume Dominant Fat | Off |
| Assume Silicone | Off |
| Adjustment Tolerance | Auto |

#### System - Adjust Volume

| | |
|---|---|
| Position | Isocenter |
| Orientation | Transversal |
| Rotation | 0.00 deg |
| A >> P | 263 mm |
| R >> L | 350 mm |
| F >> H | 350 mm |
| Reset | Off |

#### System - Tx/Rx

| | |
|---|---|
| Frequency 1H | 63.678323 MHz |
| Correction factor | 1 |
| Gain | High |
| Img. Scale Cor. | 1.000 |
| Reset | Off |
| ? Ref. amplitude 1H | 0.000 V |

#### Physio - Signal1

| | |
|---|---|
| 1st Signal/Mode | ECG/Trigger |
| Average cycle | No Signal ms |
| Average cycle | No Signal ms |
| Captured cycle | -not set- |
| Acquisition window | 591 ms |
| Trigger pulse | 1 |
| Trigger delay | 411 ms |
| TR | 179.50 ms |
| Concatenations | 1 |
| Segments | 54 |
| Phases | 1 |
| Adaptive Triggering | Off |

#### Physio - Cardiac

| | |
|---|---|
| Tagging | None |
| Magn. preparation | T2 prep. adiab. |
| T2 prep. duration 1 | 0 ms |
| T2 prep. duration 2 | 25 ms |
| T2 prep. duration 3 | 55 ms |
| Fat suppr. | None |
| Dark blood | Off |
| FoV read | 460 mm |
| FoV phase | 75.0 % |
| Phase resolution | 75 % |
| Cine | Off |
| Trajectory | Cartesian |
| Dummy heartbeats | 0 |
| Motion Correction | Standard |

#### Physio - PACE

| | |
|---|---|
| Resp. control | Breath-hold |
| Concatenations | 1 |

#### Sequence - Part 1

| | |
|---|---|
| Introduction | Off |
| Dimension | 2D |
| Reordering | Linear |
| Asymmetric echo | Weak |
| Contrasts | 1 |
| Optimization | Min. TE TR |
| Multi-slice mode | Sequential |
| Sequence type | Trufi |
| Bandwidth | 1184 Hz/Px |

#### Sequence - Part 2

| | |
|---|---|
| Define | Shots |
| Shots per slice | 1 |
| Segments | 54 |
| Trufi delta freq. | 0 Hz |
| RF pulse type | Fast |
| Gradient mode | Fast |
| Excitation | Slice-sel. |
| Flip angle mode | Constant |
| Cine | Off |

#### Sequence - Assistant

| | |
|---|---|
| Mode | Off |
| Allowed delay | 0 s |

| \\USER\Cardiac Research Protocols\IV IRON\IV IRON4\KLSShMOLLI_C2P_m |
|---|
| TA: 5.3 s PM: FIX Voxel size: 1.2×1.2×8.0 mmPAT: 2 Rel. SNR: 1.00 : tfi |

#### Properties

| | |
|---|---|
| Prio recon | Off |
| Load images to viewer | On |
| Inline movie | Off |
| Auto store images | On |
| Load images to stamp segments | Off |
| Load images to graphic segments | On |
| Auto open inline display | Off |
| Auto close inline display | Off |
| Start measurement without further preparation | On |
| Wait for user to start | Off |
| Start measurements | Single measurement |

#### Routine

| | |
|---|---|
| Slice group | 1 |
| Slices | 1 |
| Dist. factor | 25 % |
| Position | Isocenter |
| Orientation | Transversal |
| Phase enc. dir. | A >> P |
| AutoAlign | --- |
| Phase oversampling | 0 % |
| FoV read | 460 mm |
| FoV phase | 75.0 % |
| Slice thickness | 8.0 mm |
| TR | 375.62 ms |
| TE | 1.07 ms |
| Averages | 1 |
| Concatenations | 1 |
| Filter | Raw filter, Distortion Corr.(2D) |
| Coil elements | BO1-3;SP1-3 |

#### Contrast - Common

| | |
|---|---|
| TR | 375.62 ms |
| TE | 1.07 ms |
| Magn. preparation | Non-sel. IR |
| TI | 260 ms |
| Flip angle | 35 deg |
| Fat suppr. | None |
| Wrap-up Magn. | None |

#### Contrast - Dynamic

| | |
|---|---|
| Averages | 1 |
| Averaging mode | Short term |
| Reconstruction | Magn./Phase |
| Measurements | 1 |
| Multiple series | Off |

#### Resolution - Common

| | |
|---|---|
| FoV read | 460 mm |
| FoV phase | 75.0 % |
| Slice thickness | 8.0 mm |
| Base resolution | 192 |
| Phase resolution | 100 % |
| Phase partial Fourier | 6/8 |
| Trajectory | Cartesian |
| Interpolation | On |

#### Resolution - iPAT

| | |
|---|---|
| PAT mode | GRAPPA |
| Accel. factor PE | 2 |
| Ref. lines PE | 24 |
| Reference scan mode | Integrated |

#### Resolution - Filter Image

| | |
|---|---|
| Image Filter | Off |
| Distortion Corr. | On |
| Mode | 2D |
| Unfiltered images | Off |
| Prescan Normalize | Off |
| Normalize | Off |
| B1 filter | Off |

#### Resolution - Filter Rawdata

| | |
|---|---|
| Raw filter | On |
| Elliptical filter | Off |
| POCS | Off |

#### Geometry - Common

| | |
|---|---|
| Slice group | 1 |
| Slices | 1 |
| Dist. factor | 25 % |
| Position | Isocenter |
| Orientation | Transversal |
| Phase enc. dir. | A >> P |
| FoV read | 460 mm |
| FoV phase | 75.0 % |
| Slice thickness | 8.0 mm |
| TR | 375.62 ms |
| Multi-slice mode | Sequential |
| Series | Interleaved |
| Concatenations | 1 |

#### Geometry - AutoAlign

| | |
|---|---|
| Slice group | 1 |
| Position | Isocenter |
| Orientation | Transversal |
| Phase enc. dir. | A >> P |
| AutoAlign | --- |
| Initial Position | Isocenter |
| Phase | 0.0 mm |
| Read | 0.0 mm |
| Shift | 0.0 mm |
| Initial Rotation | 0.00 deg |
| Initial Orientation | Transversal |

#### Geometry - Saturation

| | |
|---|---|
| Fat suppr. | None |
| Wrap-up Magn. | None |
| Special sat. | None |

#### Geometry - Navigator

#### System - Miscellaneous

| | |
|---|---|
| Positioning mode | FIX |
| Table position | H |
| Table position | 0 mm |
| MSMA | S - C - T |
| Sagittal | R >> L |

#### System - Miscellaneous

| | |
|---|---|
| Coronal | A >> P |
| Transversal | F >> H |
| Coil Combine Mode | Adaptive Combine |
| Save uncombined | Off |
| Matrix Optimization | Off |
| Coil Focus | Flat |
| AutoAlign | --- |
| Coil Select Mode | Default |

#### System - Adjustments

| | |
|---|---|
| B0 Shim mode | Tune up |
| Adjust with body coil | On |
| Confirm freq. adjustment | Off |
| Assume Dominant Fat | Off |
| Assume Silicone | Off |
| Adjustment Tolerance | Auto |

#### System - Adjust Volume

| | |
|---|---|
| ! Position | Isocenter |
| ! Orientation | Transversal |
| ! Rotation | 0.00 deg |
| ! A >> P | 263 mm |
| ! R >> L | 350 mm |
| ! F >> H | 350 mm |
| Reset | Off |

#### System - Tx/Rx

| | |
|---|---|
| Frequency 1H | 63.678323 MHz |
| Correction factor | 1 |
| Gain | High |
| Img. Scale Cor. | 1.000 |
| Reset | Off |
| ? Ref. amplitude 1H | 0.000 V |

#### Physio - Signal1

| | |
|---|---|
| 1st Signal/Mode | ECG/Trigger |
| Average cycle | No Signal ms |
| Average cycle | No Signal ms |
| Captured cycle | -not set- |
| Acquisition window | 591 ms |
| Trigger pulse | 1 |
| Trigger delay | 215 ms |
| TR | 375.62 ms |
| Concatenations | 1 |
| Segments | 84 |
| Phases | 1 |
| Adaptive Triggering | Off |

#### Physio - Cardiac

| | |
|---|---|
| Tagging | None |
| Magn. preparation | Non-sel. IR |
| TI | 260 ms |
| Fat suppr. | None |
| Dark blood | Off |
| FoV read | 460 mm |
| FoV phase | 75.0 % |
| Phase resolution | 100 % |
| Cine | Off |
| Trajectory | Cartesian |
| Dummy heartbeats | 0 |
| Motion Correction | Standard |

#### Physio - PACE

| | |
|---|---|
| Resp. control | Off |

#### Physio - PACE

| | |
|---|---|
| Concatenations | 1 |

#### Sequence - Part 1

| | |
|---|---|
| Introduction | Off |
| Dimension | 2D |
| Reordering | Linear |
| Asymmetric echo | Weak |
| Contrasts | 1 |
| Optimization | Min. TE TR |
| Multi-slice mode | Sequential |
| Sequence type | Trufi |
| Bandwidth | 898 Hz/Px |

#### Sequence - Part 2

| | |
|---|---|
| Define | Shots |
| Shots per slice | 1 |
| Segments | 84 |
| Trufi delta freq. | 0 Hz |
| RF pulse type | Fast |
| Gradient mode | Fast |
| Excitation | Slice-sel. |
| Flip angle mode | Constant |
| Cine | Off |

#### Sequence - Assistant

| | |
|---|---|
| Mode | Off |
| Allowed delay | 0 s |

| \\USER\Cardiac Research Protocols\IV IRON\IV IRON4\KLST2StarMap_8echo_heart |
|---|
| TA: 9.4 s PM: FIX Voxel size: 1.8×1.8×8.0 mmPAT: 2 Rel. SNR: 1.00 : fl_r |

#### Properties

| | |
|---|---|
| Prio recon | Off |
| Load images to viewer | On |
| Inline movie | Off |
| Auto store images | On |
| Load images to stamp segments | Off |
| Load images to graphic segments | On |
| Auto open inline display | Off |
| Auto close inline display | Off |
| Start measurement without further preparation | On |
| Wait for user to start | Off |
| Start measurements | Single measurement |

#### Routine

| | |
|---|---|
| Slice group | 1 |
| Slices | 1 |
| Dist. factor | 20 % |
| Position | Isocenter |
| Orientation | Transversal |
| Phase enc. dir. | A >> P |
| AutoAlign | --- |
| Phase oversampling | 0 % |
| FoV read | 460 mm |
| FoV phase | 75.0 % |
| Slice thickness | 8.0 mm |
| TR | 590.00 ms |
| TE 1 | 1.91 ms |
| TE 2 | 3.85 ms |
| TE 3 | 5.79 ms |
| TE 4 | 7.73 ms |
| TE 5 | 9.67 ms |
| TE 6 | 11.61 ms |
| TE 7 | 13.55 ms |
| TE 8 | 15.49 ms |
| Averages | 1 |
| Concatenations | 1 |
| Filter | Distortion Corr.(2D) |
| Coil elements | BO1-3;SP1-3 |

#### Contrast - Common

| | |
|---|---|
| TR | 590.00 ms |
| TE 1 | 1.91 ms |
| TE 2 | 3.85 ms |
| TE 3 | 5.79 ms |
| TE 4 | 7.73 ms |
| TE 5 | 9.67 ms |
| TE 6 | 11.61 ms |
| TE 7 | 13.55 ms |
| TE 8 | 15.49 ms |
| Flip angle | 20 deg |
| Fat suppr. | Fat sat. |
| Wrap-up Magn. | None |

#### Contrast - Dynamic

| | |
|---|---|
| Averages | 1 |
| Averaging mode | Short term |
| Reconstruction | Magnitude |
| Measurements | 1 |
| Multiple series | Off |

#### Resolution - Common

| | |
|---|---|
| FoV read | 460 mm |
| FoV phase | 75.0 % |
| Slice thickness | 8.0 mm |
| Base resolution | 256 |
| Phase resolution | 60 % |
| Phase partial Fourier | Off |
| Trajectory | Cartesian |
| Interpolation | Off |

#### Resolution - iPAT

| | |
|---|---|
| PAT mode | GRAPPA |
| Accel. factor PE | 2 |
| Ref. lines PE | 24 |
| Reference scan mode | Integrated |

#### Resolution - Filter Image

| | |
|---|---|
| Image Filter | Off |
| Distortion Corr. | On |
| Mode | 2D |
| Unfiltered images | Off |
| Prescan Normalize | Off |
| Normalize | Off |
| B1 filter | Off |

#### Resolution - Filter Rawdata

| | |
|---|---|
| Raw filter | Off |
| Elliptical filter | Off |
| POCS | Off |

#### Geometry - Common

| | |
|---|---|
| Slice group | 1 |
| Slices | 1 |
| Dist. factor | 20 % |
| Position | Isocenter |
| Orientation | Transversal |
| Phase enc. dir. | A >> P |
| FoV read | 460 mm |
| FoV phase | 75.0 % |
| Slice thickness | 8.0 mm |
| TR | 590.00 ms |
| Multi-slice mode | Sequential |
| Series | Base To Apex |
| Concatenations | 1 |

#### Geometry - AutoAlign

| | |
|---|---|
| Slice group | 1 |
| Position | Isocenter |
| Orientation | Transversal |
| Phase enc. dir. | A >> P |
| AutoAlign | --- |
| Initial Position | Isocenter |
| Phase | 0.0 mm |
| Read | 0.0 mm |
| Shift | 0.0 mm |
| Initial Rotation | 0.00 deg |
| Initial Orientation | Transversal |

#### Geometry - Saturation

| | |
|---|---|
| Fat suppr. | Fat sat. |
| Wrap-up Magn. | None |

#### Geometry - Saturation

| | |
|---|---|
| Special sat. | None |

#### Geometry - Navigator

#### System - Miscellaneous

| | |
|---|---|
| Positioning mode | FIX |
| Table position | H |
| Table position | 0 mm |
| MSMA | S - C - T |
| Sagittal | R >> L |
| Coronal | A >> P |
| Transversal | F >> H |
| Coil Combine Mode | Adaptive Combine |
| Save uncombined | Off |
| Matrix Optimization | Off |
| Coil Focus | Flat |
| AutoAlign | --- |
| Coil Select Mode | Default |

#### System - Adjustments

| | |
|---|---|
| B0 Shim mode | Tune up |
| Adjust with body coil | Off |
| Confirm freq. adjustment | Off |
| Assume Dominant Fat | Off |
| Assume Silicone | Off |
| Adjustment Tolerance | Auto |

#### System - Adjust Volume

| | |
|---|---|
| ! Position | Isocenter |
| ! Orientation | Transversal |
| ! Rotation | 0.00 deg |
| ! A >> P | 263 mm |
| ! R >> L | 350 mm |
| ! F >> H | 350 mm |
| Reset | Off |

#### System - Tx/Rx

| | |
|---|---|
| Frequency 1H | 63.678323 MHz |
| Correction factor | 1 |
| Gain | High |
| Img. Scale Cor. | 1.000 |
| Reset | Off |
| ? Ref. amplitude 1H | 0.000 V |

#### Physio - Signal1

| | |
|---|---|
| 1st Signal/Mode | ECG/Trigger |
| Average cycle | No Signal ms |
| Average cycle | No Signal ms |
| Captured cycle | -not set- |
| Acquisition window | 590 ms |
| Trigger pulse | 2 |
| Trigger delay | 0 ms |
| TR | 590.00 ms |
| Concatenations | 1 |
| Segments | 9 |
| Phases | 1 |
| Adaptive Triggering | Off |

#### Physio - Cardiac

| | |
|---|---|
| Tagging | None |
| Fat suppr. | Fat sat. |
| Dark blood | On |
| Dark blood thickness | 200 % |

#### Physio - Cardiac

| | |
|---|---|
| FoV read | 460 mm |
| FoV phase | 75.0 % |
| Phase resolution | 60 % |
| Cine | Off |
| Trajectory | Cartesian |
| Dummy heartbeats | 0 |

#### Physio - PACE

| | |
|---|---|
| Resp. control | Breath-hold |
| Concatenations | 1 |

#### Inline - Common

| | |
|---|---|
| Subtract | Off |
| Measurements | 1 |
| StdDev | Off |
| Save original images | On |

#### Inline - Cardiac

| | |
|---|---|
| Inline Evaluation | T2* map |
| Contrasts | 8 |
| TE 1 | 1.91 ms |
| TE 2 | 3.85 ms |
| TE 3 | 5.79 ms |
| TE 4 | 7.73 ms |
| TE 5 | 9.67 ms |
| TE 6 | 11.61 ms |
| TE 7 | 13.55 ms |
| TE 8 | 15.49 ms |
| TR | 590.00 ms |
| Save original images | On |

#### Inline - MIP

| | |
|---|---|
| MIP-Sag | Off |
| MIP-Cor | Off |
| MIP-Tra | Off |
| MIP-Time | Off |
| Save original images | On |

#### Inline - Composing

| | |
|---|---|
| Inline Composing | Off |
| Distortion Corr. | On |
| Mode | 2D |
| Unfiltered images | Off |

#### Sequence - Part 1

| | |
|---|---|
| Introduction | Off |
| Dimension | 2D |
| Reordering | Linear |
| Asymmetric echo | Weak |
| Contrasts | 8 |
| Flow comp. 1 | Yes |
| Readout mode | Monopolar |
| Optimization | Min. TE |
| Multi-slice mode | Sequential |
| Echo spacing | 17.4 ms |
| Sequence type | Gre |
| Bandwidth 1 | 814 Hz/Px |
| Bandwidth 2 | 814 Hz/Px |
| Bandwidth 3 | 814 Hz/Px |
| Bandwidth 4 | 814 Hz/Px |
| Bandwidth 5 | 814 Hz/Px |
| Bandwidth 6 | 814 Hz/Px |
| Bandwidth 7 | 814 Hz/Px |
| Bandwidth 8 | 814 Hz/Px |

#### Sequence - Part 2

| | |
|---|---|
| Define | Segments |
| Segments | 9 |
| RF pulse type | Fast |
| Gradient mode | Fast |
| Excitation | Slice-sel. |
| Flip angle mode | Constant |
| RF spoiling | On |
| Phase Enc. Rewinder | On |
| Cine | Off |

#### Sequence - Assistant

| | |
|---|---|
| Mode | Off |
| Allowed delay | 0 s |

#### \\USER\Cardiac Research Protocols\IV IRON\IV IRON4\KLSFB_MOCO_gt_T2star_DB_8e_128 FS_HiFE

TA: 9.4 s PM: FIX Voxel size: 3.6×3.6×8.0 mmPAT: 4 Rel. SNR: 1.00 : tfl

##### Properties

| | |
|---|---|
| Prio recon | Off |
| Load images to viewer | On |
| Inline movie | Off |
| Auto store images | On |
| Load images to stamp segments | Off |
| Load images to graphic segments | On |
| Auto open inline display | Off |
| Auto close inline display | Off |
| Start measurement without further preparation | On |
| Wait for user to start | Off |
| Start measurements | Single measurement |

##### Routine

| | |
|---|---|
| Slice group | 1 |
| Slices | 1 |
| Dist. factor | 20 % |
| Position | Isocenter |
| Orientation | Transversal |
| Phase enc. dir. | A >> P |
| AutoAlign | --- |
| Phase oversampling | 0 % |
| FoV read | 460 mm |
| FoV phase | 75.0 % |
| Slice thickness | 8.0 mm |
| TR | 590.00 ms |
| TE 1 | 1.07 ms |
| TE 2 | 2.42 ms |
| TE 3 | 3.77 ms |
| TE 4 | 5.12 ms |
| TE 5 | 6.47 ms |
| TE 6 | 7.82 ms |
| TE 7 | 9.17 ms |
| TE 8 | 10.52 ms |
| Averages | 1 |
| Concatenations | 1 |
| Filter | Distortion Corr.(2D) |
| Coil elements | BO1-3;SP1-3 |

##### Contrast - Common

| | |
|---|---|
| TR | 590.00 ms |
| TE 1 | 1.07 ms |
| TE 2 | 2.42 ms |
| TE 3 | 3.77 ms |
| TE 4 | 5.12 ms |
| TE 5 | 6.47 ms |
| TE 6 | 7.82 ms |
| TE 7 | 9.17 ms |
| TE 8 | 10.52 ms |
| Magn. preparation | None |
| Flip angle | 18 deg |
| Fat suppr. | Fat sat. |
| Wrap-up Magn. | None |

##### Contrast - Dynamic

| | |
|---|---|
| Averages | 1 |
| Averaging mode | Long term |
| Reconstruction | Magnitude |
| Measurements | 16 |

##### Contrast - Dynamic

| | |
|---|---|
| Pause after meas. 1 | 0.0 s |
| Pause after meas. 2 | 0.0 s |
| Pause after meas. 3 | 0.0 s |
| Pause after meas. 4 | 0.0 s |
| Pause after meas. 5 | 0.0 s |
| Pause after meas. 6 | 0.0 s |
| Pause after meas. 7 | 0.0 s |
| Pause after meas. 8 | 0.0 s |
| Pause after meas. 9 | 0.0 s |
| Pause after meas. 10 | 0.0 s |
| Pause after meas. 11 | 0.0 s |
| Pause after meas. 12 | 0.0 s |
| Pause after meas. 13 | 0.0 s |
| Pause after meas. 14 | 0.0 s |
| Pause after meas. 15 | 0.0 s |
| Multiple series | Off |

##### Resolution - Common

| | |
|---|---|
| FoV read | 460 mm |
| FoV phase | 75.0 % |
| Slice thickness | 8.0 mm |
| Base resolution | 128 |
| Phase resolution | 100 % |
| Phase partial Fourier | Off |
| Trajectory | Cartesian |
| Interpolation | Off |

##### Resolution - iPAT

| | |
|---|---|
| PAT mode | GRAPPA |
| Accel. factor PE | 4 |
| Reference scan mode | T-PAT |

##### Resolution - Filter Image

| | |
|---|---|
| Image Filter | Off |
| Distortion Corr. | On |
| Mode | 2D |
| Unfiltered images | Off |
| Prescan Normalize | Off |
| Normalize | Off |
| B1 filter | Off |

##### Resolution - Filter Rawdata

| | |
|---|---|
| Raw filter | Off |
| Elliptical filter | Off |
| POCS | Off |

##### Geometry - Common

| | |
|---|---|
| Slice group | 1 |
| Slices | 1 |
| Dist. factor | 20 % |
| Position | Isocenter |
| Orientation | Transversal |
| Phase enc. dir. | A >> P |
| FoV read | 460 mm |
| FoV phase | 75.0 % |
| Slice thickness | 8.0 mm |
| TR | 590.00 ms |
| Multi-slice mode | Single shot |
| Series | Interleaved |

#### Geometry - Common

| | |
|---|---|
| Concatenations | 1 |

#### Geometry - AutoAlign

| | |
|---|---|
| Slice group | 1 |
| Position | Isocenter |
| Orientation | Transversal |
| Phase enc. dir. | A >> P |
| AutoAlign | --- |
| Initial Position | Isocenter |
| Phase | 0.0 mm |
| Read | 0.0 mm |
| Shift | 0.0 mm |
| Initial Rotation | 0.00 deg |
| Initial Orientation | Transversal |

#### Geometry - Saturation

| | |
|---|---|
| Fat suppr. | Fat sat. |
| Wrap-up Magn. | None |
| Special sat. | None |

#### Geometry - Navigator

#### System - Miscellaneous

| | |
|---|---|
| Positioning mode | FIX |
| Table position | H |
| Table position | 0 mm |
| MSMA | S - C - T |
| Sagittal | R >> L |
| Coronal | A >> P |
| Transversal | F >> H |
| Coil Combine Mode | Adaptive Combine |
| Save uncombined | Off |
| Matrix Optimization | Off |
| Coil Focus | Flat |
| AutoAlign | --- |
| Coil Select Mode | Default |

#### System - Adjustments

| | |
|---|---|
| B0 Shim mode | Tune up |
| Adjust with body coil | On |
| Confirm freq. adjustment | Off |
| Assume Dominant Fat | Off |
| Assume Silicone | Off |
| Adjustment Tolerance | Auto |

#### System - Adjust Volume

| | |
|---|---|
| ! Position | Isocenter |
| ! Orientation | Transversal |
| ! Rotation | 0.00 deg |
| ! A >> P | 263 mm |
| ! R >> L | 350 mm |
| ! F >> H | 350 mm |
| Reset | Off |

#### System - Tx/Rx

| | |
|---|---|
| Frequency 1H | 63.678323 MHz |
| Correction factor | 1 |
| Gain | High |
| Img. Scale Cor. | 1.000 |
| Reset | Off |
| ? Ref. amplitude 1H | 0.000 V |

#### Physio - Signal1

| | |
|---|---|
| 1st Signal/Mode | ECG/Trigger |
| Average cycle | No Signal ms |
| Average cycle | No Signal ms |
| Captured cycle | -not set- |
| Acquisition window | 590 ms |
| Trigger pulse | 1 |
| Trigger delay | 0 ms |
| TR | 590.00 ms |
| Concatenations | 1 |
| Segments | 24 |
| Phases | 1 |
| Adaptive Triggering | Off |

#### Physio - Cardiac

| | |
|---|---|
| Tagging | None |
| Magn. preparation | None |
| Fat suppr. | Fat sat. |
| Dark blood | On |
| Dark blood thickness | 300 % |
| FoV read | 460 mm |
| FoV phase | 75.0 % |
| Phase resolution | 100 % |
| Cine | Off |
| Trajectory | Cartesian |
| Dummy heartbeats | 0 |

#### Physio - PACE

| | |
|---|---|
| Resp. control | Off |
| Concatenations | 1 |

#### Inline - Common

| | |
|---|---|
| Subtract | Off |
| Measurements | 16 |
| StdDev | Off |
| Save original images | On |

#### Inline - Cardiac

| | |
|---|---|
| Inline Evaluation | Off |
| Magn. preparation | None |
| Contrasts | 8 |
| TE 1 | 1.07 ms |
| TE 2 | 2.42 ms |
| TE 3 | 3.77 ms |
| TE 4 | 5.12 ms |
| TE 5 | 6.47 ms |
| TE 6 | 7.82 ms |
| TE 7 | 9.17 ms |
| TE 8 | 10.52 ms |
| TR | 590.00 ms |
| Save original images | On |

#### Inline - MIP

| | |
|---|---|
| MIP-Sag | Off |
| MIP-Cor | Off |
| MIP-Tra | Off |
| MIP-Time | Off |
| Save original images | On |

#### Inline - Composing

| | |
|---|---|
| Inline Composing | Off |
| Distortion Corr. | On |
| Mode | 2D |
| Unfiltered images | Off |

#### Sequence - Part 1

| | |
|---|---|
| Introduction | Off |
| Dimension | 2D |
| Reordering | Linear |
| Asymmetric echo | Off |
| Contrasts | 8 |
| Flow comp. 1 | No |
| Readout mode | Monopolar |
| Optimization | Min. TE |
| Multi-slice mode | Single shot |
| Echo spacing | 11.4 ms |
| Sequence type | Gre |
| Bandwidth 1 | 1502 Hz/Px |
| Bandwidth 2 | 1502 Hz/Px |
| Bandwidth 3 | 1502 Hz/Px |
| Bandwidth 4 | 1502 Hz/Px |
| Bandwidth 5 | 1502 Hz/Px |
| Bandwidth 6 | 1502 Hz/Px |
| Bandwidth 7 | 1502 Hz/Px |
| Bandwidth 8 | 1502 Hz/Px |

#### Sequence - Part 2

| | |
|---|---|
| Define | Shots |
| Shots per slice | 1 |
| Segments | 24 |
| RF pulse type | Fast |
| Gradient mode | Fast |
| Excitation | Slice-sel. |
| Flip angle mode | Constant |
| RF spoiling | On |
| Phase Enc. Rewinder | On |
| Cine | Off |

#### Sequence - Special

| | |
|---|---|
| FatWater Separation | On |
| Multi-echo Images | Off |
| In-Opp Phase Images | Off |
| Frequency Map | Off |
| T2* Map | Off |
| Motion Correction | Off |
| MoCo Averaging Mode | Complex MoCo |
| MoCo Images Only? | Off |
| No. of Interleaves | 0 |

#### Sequence - Assistant

| | |
|---|---|
| Mode | Off |
| Allowed delay | 0 s |

**\\USER\Cardiac Research Protocols\IV IRON\IV IRON4\KLSFB_MOCO_gt_T2star_DB_8e_160 FS_LowNFE**

TA: 9.4 s PM: FIX Voxel size: 2.9×2.9×8.0 mmPAT: 4 Rel. SNR: 1.00 : tfl

#### Properties

| | |
|---|---|
| Prio recon | Off |
| Load images to viewer | On |
| Inline movie | Off |
| Auto store images | On |
| Load images to stamp segments | Off |
| Load images to graphic segments | On |
| Auto open inline display | Off |
| Auto close inline display | Off |
| Start measurement without further preparation | On |
| Wait for user to start | Off |
| Start measurements | Single measurement |

#### Routine

| | |
|---|---|
| Slice group | 1 |
| Slices | 1 |
| Dist. factor | 20 % |
| Position | Isocenter |
| Orientation | Transversal |
| Phase enc. dir. | A >> P |
| AutoAlign | --- |
| Phase oversampling | 0 % |
| FoV read | 460 mm |
| FoV phase | 75.0 % |
| Slice thickness | 8.0 mm |
| TR | 590.00 ms |
| TE 1 | 1.2 ms |
| TE 2 | 2.87 ms |
| TE 3 | 4.54 ms |
| TE 4 | 6.21 ms |
| TE 5 | 7.88 ms |
| TE 6 | 9.55 ms |
| TE 7 | 11.22 ms |
| TE 8 | 12.89 ms |
| Averages | 1 |
| Concatenations | 1 |
| Filter | Distortion Corr.(2D) |
| Coil elements | BO1-3;SP1-3 |

#### Contrast - Common

| | |
|---|---|
| TR | 590.00 ms |
| TE 1 | 1.2 ms |
| TE 2 | 2.87 ms |
| TE 3 | 4.54 ms |
| TE 4 | 6.21 ms |
| TE 5 | 7.88 ms |
| TE 6 | 9.55 ms |
| TE 7 | 11.22 ms |
| TE 8 | 12.89 ms |
| Magn. preparation | None |
| Flip angle | 18 deg |
| Fat suppr. | Fat sat. |
| Wrap-up Magn. | None |

#### Contrast - Dynamic

| | |
|---|---|
| Averages | 1 |
| Averaging mode | Long term |
| Reconstruction | Magnitude |
| Measurements | 16 |

#### Contrast - Dynamic

| | |
|---|---|
| Pause after meas. 1 | 0.0 s |
| Pause after meas. 2 | 0.0 s |
| Pause after meas. 3 | 0.0 s |
| Pause after meas. 4 | 0.0 s |
| Pause after meas. 5 | 0.0 s |
| Pause after meas. 6 | 0.0 s |
| Pause after meas. 7 | 0.0 s |
| Pause after meas. 8 | 0.0 s |
| Pause after meas. 9 | 0.0 s |
| Pause after meas. 10 | 0.0 s |
| Pause after meas. 11 | 0.0 s |
| Pause after meas. 12 | 0.0 s |
| Pause after meas. 13 | 0.0 s |
| Pause after meas. 14 | 0.0 s |
| Pause after meas. 15 | 0.0 s |
| Multiple series | Off |

#### Resolution - Common

| | |
|---|---|
| FoV read | 460 mm |
| FoV phase | 75.0 % |
| Slice thickness | 8.0 mm |
| Base resolution | 160 |
| Phase resolution | 87 % |
| Phase partial Fourier | Off |
| Trajectory | Cartesian |
| Interpolation | Off |

#### Resolution - iPAT

| | |
|---|---|
| PAT mode | GRAPPA |
| Accel. factor PE | 4 |
| Reference scan mode | T-PAT |

#### Resolution - Filter Image

| | |
|---|---|
| Image Filter | Off |
| Distortion Corr. | On |
| Mode | 2D |
| Unfiltered images | Off |
| Prescan Normalize | Off |
| Normalize | Off |
| B1 filter | Off |

#### Resolution - Filter Rawdata

| | |
|---|---|
| Raw filter | Off |
| Elliptical filter | Off |
| POCS | Off |

#### Geometry - Common

| | |
|---|---|
| Slice group | 1 |
| Slices | 1 |
| Dist. factor | 20 % |
| Position | Isocenter |
| Orientation | Transversal |
| Phase enc. dir. | A >> P |
| FoV read | 460 mm |
| FoV phase | 75.0 % |
| Slice thickness | 8.0 mm |
| TR | 590.00 ms |
| Multi-slice mode | Single shot |
| Series | Interleaved |

#### Geometry - Common

| | |
|---|---|
| Concatenations | 1 |

#### Geometry - AutoAlign

| | |
|---|---|
| Slice group | 1 |
| Position | Isocenter |
| Orientation | Transversal |
| Phase enc. dir. | A >> P |
| AutoAlign | --- |
| Initial Position | Isocenter |
| Phase | 0.0 mm |
| Read | 0.0 mm |
| Shift | 0.0 mm |
| Initial Rotation | 0.00 deg |
| Initial Orientation | Transversal |

#### Geometry - Saturation

| | |
|---|---|
| Fat suppr. | Fat sat. |
| Wrap-up Magn. | None |
| Special sat. | None |

#### Geometry - Navigator

#### System - Miscellaneous

| | |
|---|---|
| Positioning mode | FIX |
| Table position | H |
| Table position | 0 mm |
| MSMA | S - C - T |
| Sagittal | R >> L |
| Coronal | A >> P |
| Transversal | F >> H |
| Coil Combine Mode | Adaptive Combine |
| Save uncombined | Off |
| Matrix Optimization | Off |
| Coil Focus | Flat |
| AutoAlign | --- |
| Coil Select Mode | Default |

#### System - Adjustments

| | |
|---|---|
| B0 Shim mode | Tune up |
| Adjust with body coil | On |
| Confirm freq. adjustment | Off |
| Assume Dominant Fat | Off |
| Assume Silicone | Off |
| Adjustment Tolerance | Auto |

#### System - Adjust Volume

| | |
|---|---|
| ! Position | Isocenter |
| ! Orientation | Transversal |
| ! Rotation | 0.00 deg |
| ! A >> P | 263 mm |
| ! R >> L | 350 mm |
| ! F >> H | 350 mm |
| Reset | Off |

#### System - Tx/Rx

| | |
|---|---|
| Frequency 1H | 63.678323 MHz |
| Correction factor | 1 |
| Gain | High |
| Img. Scale Cor. | 1.000 |
| Reset | Off |
| ? Ref. amplitude 1H | 0.000 V |

#### Physio - Signal1

| | |
|---|---|
| 1st Signal/Mode | ECG/Trigger |
| Average cycle | No Signal ms |
| Average cycle | No Signal ms |
| Captured cycle | -not set- |
| Acquisition window | 590 ms |
| Trigger pulse | 1 |
| Trigger delay | 0 ms |
| TR | 590.00 ms |
| Concatenations | 1 |
| Segments | 26 |
| Phases | 1 |
| Adaptive Triggering | Off |

#### Physio - Cardiac

| | |
|---|---|
| Tagging | None |
| Magn. preparation | None |
| Fat suppr. | Fat sat. |
| Dark blood | On |
| Dark blood thickness | 300 % |
| FoV read | 460 mm |
| FoV phase | 75.0 % |
| Phase resolution | 87 % |
| Cine | Off |
| Trajectory | Cartesian |
| Dummy heartbeats | 0 |

#### Physio - PACE

| | |
|---|---|
| Resp. control | Off |
| Concatenations | 1 |

#### Inline - Common

| | |
|---|---|
| Subtract | Off |
| Measurements | 16 |
| StdDev | Off |
| Save original images | On |

#### Inline - Cardiac

| | |
|---|---|
| Inline Evaluation | Off |
| Magn. preparation | None |
| Contrasts | 8 |
| TE 1 | 1.2 ms |
| TE 2 | 2.87 ms |
| TE 3 | 4.54 ms |
| TE 4 | 6.21 ms |
| TE 5 | 7.88 ms |
| TE 6 | 9.55 ms |
| TE 7 | 11.22 ms |
| TE 8 | 12.89 ms |
| TR | 590.00 ms |
| Save original images | On |

#### Inline - MIP

| | |
|---|---|
| MIP-Sag | Off |
| MIP-Cor | Off |
| MIP-Tra | Off |
| MIP-Time | Off |
| Save original images | On |

#### Inline - Composing

| | |
|---|---|
| Inline Composing | Off |
| Distortion Corr. | On |
| Mode | 2D |
| Unfiltered images | Off |

#### Sequence - Part 1

| | |
|---|---|
| Introduction | Off |
| Dimension | 2D |
| Reordering | Linear |
| Asymmetric echo | Off |
| Contrasts | 8 |
| Flow comp. 1 | No |
| Readout mode | Monopolar |
| Optimization | Min. TE |
| Multi-slice mode | Single shot |
| Echo spacing | 13.9 ms |
| Sequence type | Gre |
| Bandwidth 1 | 1078 Hz/Px |
| Bandwidth 2 | 1078 Hz/Px |
| Bandwidth 3 | 1078 Hz/Px |
| Bandwidth 4 | 1078 Hz/Px |
| Bandwidth 5 | 1078 Hz/Px |
| Bandwidth 6 | 1078 Hz/Px |
| Bandwidth 7 | 1078 Hz/Px |
| Bandwidth 8 | 1078 Hz/Px |

#### Sequence - Part 2

| | |
|---|---|
| Define | Shots |
| Shots per slice | 1 |
| Segments | 26 |
| RF pulse type | Fast |
| Gradient mode | Fast |
| Excitation | Slice-sel. |
| Flip angle mode | Constant |
| RF spoiling | On |
| Phase Enc. Rewinder | On |
| Cine | Off |

#### Sequence - Special

| | |
|---|---|
| FatWater Separation | On |
| Multi-echo Images | Off |
| In-Opp Phase Images | Off |
| Frequency Map | Off |
| T2* Map | Off |
| Motion Correction | Off |
| MoCo Averaging Mode | Complex MoCo |
| MoCo Images Only? | Off |
| No. of Interleaves | 0 |

#### Sequence - Assistant

| | |
|---|---|
| Mode | Off |
| Allowed delay | 0 s |

| \\USER\Cardiac Research Protocols\IV IRON\IV IRON4\KLSnonBH_T2StarMap_12echo_liver |
|---|
| TA: 0:23 PM: FIX Voxel size: 3.6×3.6×8.0 mmPAT: Off Rel. SNR: 1.00 : fl |

#### Properties

| | |
|---|---|
| Prio recon | Off |
| Load images to viewer | On |
| Inline movie | Off |
| Auto store images | On |
| Load images to stamp segments | Off |
| Load images to graphic segments | On |
| Auto open inline display | Off |
| Auto close inline display | Off |
| Start measurement without further preparation | On |
| Wait for user to start | Off |
| Start measurements | Single measurement |

#### Routine

| | |
|---|---|
| Slice group | 1 |
| Slices | 1 |
| Dist. factor | 20 % |
| Position | Isocenter |
| Orientation | Transversal |
| Phase enc. dir. | A >> P |
| AutoAlign | --- |
| Phase oversampling | 20 % |
| FoV read | 460 mm |
| FoV phase | 75.0 % |
| Slice thickness | 8.0 mm |
| TR | 200.00 ms |
| TE 1 | 0.92 ms |
| TE 2 | 2.07 ms |
| TE 3 | 3.22 ms |
| TE 4 | 4.37 ms |
| TE 5 | 5.52 ms |
| TE 6 | 6.67 ms |
| TE 7 | 7.82 ms |
| TE 8 | 8.97 ms |
| TE 9 | 10.12 ms |
| TE 10 | 11.27 ms |
| TE 11 | 12.42 ms |
| TE 12 | 13.57 ms |
| Averages | 1 |
| Concatenations | 1 |
| Filter | Distortion Corr.(2D) |
| Coil elements | BO1-3;SP1-3 |

#### Contrast - Common

| | |
|---|---|
| TR | 200.00 ms |
| TE 1 | 0.92 ms |
| TE 2 | 2.07 ms |
| TE 3 | 3.22 ms |
| TE 4 | 4.37 ms |
| TE 5 | 5.52 ms |
| TE 6 | 6.67 ms |
| TE 7 | 7.82 ms |
| TE 8 | 8.97 ms |
| TE 9 | 10.12 ms |
| TE 10 | 11.27 ms |
| TE 11 | 12.42 ms |
| TE 12 | 13.57 ms |
| Flip angle | 20 deg |
| Fat suppr. | Fat sat. |
| Wrap-up Magn. | None |

#### Contrast - Dynamic

| | |
|---|---|
| Averages | 1 |
| Averaging mode | Short term |
| Reconstruction | Magnitude |
| Measurements | 1 |
| Multiple series | Each measurement |

#### Resolution - Common

| | |
|---|---|
| FoV read | 460 mm |
| FoV phase | 75.0 % |
| Slice thickness | 8.0 mm |
| Base resolution | 128 |
| Phase resolution | 100 % |
| Phase partial Fourier | Off |
| Trajectory | Cartesian |
| Interpolation | Off |

#### Resolution - iPAT

| | |
|---|---|
| PAT mode | None |

#### Resolution - Filter Image

| | |
|---|---|
| Image Filter | Off |
| Distortion Corr. | On |
| Mode | 2D |
| Unfiltered images | Off |
| Prescan Normalize | Off |
| Normalize | Off |
| B1 filter | Off |

#### Resolution - Filter Rawdata

| | |
|---|---|
| Raw filter | Off |
| Elliptical filter | Off |
| POCS | Off |

#### Geometry - Common

| | |
|---|---|
| Slice group | 1 |
| Slices | 1 |
| Dist. factor | 20 % |
| Position | Isocenter |
| Orientation | Transversal |
| Phase enc. dir. | A >> P |
| FoV read | 460 mm |
| FoV phase | 75.0 % |
| Slice thickness | 8.0 mm |
| TR | 200.00 ms |
| Multi-slice mode | Sequential |
| Series | Ascending |
| Concatenations | 1 |

#### Geometry - AutoAlign

| | |
|---|---|
| Slice group | 1 |
| Position | Isocenter |
| Orientation | Transversal |
| Phase enc. dir. | A >> P |
| AutoAlign | --- |
| Initial Position | Isocenter |
| Phase | 0.0 mm |
| Read | 0.0 mm |
| Shift | 0.0 mm |
| Initial Rotation | 0.00 deg |
| Initial Orientation | Transversal |

##### Geometry - Saturation

| | |
|---|---|
| Fat suppr. | Fat sat. |
| Wrap-up Magn. | None |
| Special sat. | None |

##### Geometry - Navigator

##### System - Miscellaneous

| | |
|---|---|
| Positioning mode | FIX |
| Table position | H |
| Table position | 0 mm |
| MSMA | S - C - T |
| Sagittal | R >> L |
| Coronal | A >> P |
| Transversal | F >> H |
| Coil Combine Mode | Adaptive Combine |
| Save uncombined | Off |
| Matrix Optimization | Off |
| Coil Focus | Flat |
| AutoAlign | --- |
| Coil Select Mode | Default |

##### System - Adjustments

| | |
|---|---|
| B0 Shim mode | Tune up |
| Adjust with body coil | Off |
| Confirm freq. adjustment | Off |
| Assume Dominant Fat | Off |
| Assume Silicone | Off |
| Adjustment Tolerance | Auto |

##### System - Adjust Volume

| | |
|---|---|
| ! Position | Isocenter |
| ! Orientation | Transversal |
| ! Rotation | 0.00 deg |
| ! A >> P | 263 mm |
| ! R >> L | 350 mm |
| ! F >> H | 350 mm |
| Reset | Off |

##### System - Tx/Rx

| | |
|---|---|
| Frequency 1H | 63.678323 MHz |
| Correction factor | 1 |
| Gain | High |
| Img. Scale Cor. | 1.000 |
| Reset | Off |
| ? Ref. amplitude 1H | 0.000 V |

##### Physio - Signal1

| | |
|---|---|
| 1st Signal/Mode | None |
| TR | 200.00 ms |
| Concatenations | 1 |
| Segments | 1 |

##### Physio - Cardiac

| | |
|---|---|
| Tagging | None |
| Fat suppr. | Fat sat. |
| Dark blood | Off |
| FoV read | 460 mm |
| FoV phase | 75.0 % |
| Phase resolution | 100 % |
| Cine | Off |
| Trajectory | Cartesian |
| Dummy heartbeats | 0 |

##### Physio - PACE

| | |
|---|---|
| Resp. control | Off |
| Concatenations | 1 |

##### Inline - Common

| | |
|---|---|
| Subtract | Off |
| Measurements | 1 |
| StdDev | Off |
| Save original images | On |

##### Inline - Cardiac

| | |
|---|---|
| Inline Evaluation | T2* map |
| Contrasts | 12 |
| TE 1 | 0.92 ms |
| TE 2 | 2.07 ms |
| TE 3 | 3.22 ms |
| TE 4 | 4.37 ms |
| TE 5 | 5.52 ms |
| TE 6 | 6.67 ms |
| TE 7 | 7.82 ms |
| TE 8 | 8.97 ms |
| TE 9 | 10.12 ms |
| TE 10 | 11.27 ms |
| TE 11 | 12.42 ms |
| TE 12 | 13.57 ms |
| TR | 200.00 ms |
| Save original images | On |

##### Inline - MIP

| | |
|---|---|
| MIP-Sag | Off |
| MIP-Cor | Off |
| MIP-Tra | Off |
| MIP-Time | Off |
| Save original images | On |

##### Inline - Composing

| | |
|---|---|
| Inline Composing | Off |
| Distortion Corr. | On |
| Mode | 2D |
| Unfiltered images | Off |

##### Sequence - Part 1

| | |
|---|---|
| Introduction | Off |
| Dimension | 2D |
| Reordering | Linear |
| Asymmetric echo | Weak |
| Contrasts | 12 |
| Flow comp. 1 | No |
| Readout mode | Monopolar |
| Optimization | Min. TE |
| Multi-slice mode | Sequential |
| Echo spacing | 14.8 ms |
| Sequence type | Gre |
| Bandwidth 1 | 1953 Hz/Px |
| Bandwidth 2 | 1953 Hz/Px |
| Bandwidth 3 | 1953 Hz/Px |
| Bandwidth 4 | 1953 Hz/Px |
| Bandwidth 5 | 1953 Hz/Px |
| Bandwidth 6 | 1953 Hz/Px |
| Bandwidth 7 | 1953 Hz/Px |
| Bandwidth 8 | 1953 Hz/Px |
| Bandwidth 9 | 1953 Hz/Px |
| Bandwidth 10 | 1953 Hz/Px |
| Bandwidth 11 | 1953 Hz/Px |
| Bandwidth 12 | 1953 Hz/Px |

#### Sequence - Part 2

| | |
|---|---|
| Define | Segments |
| Segments | 1 |
| RF pulse type | Fast |
| Gradient mode | Fast |
| Excitation | Slice-sel. |
| Flip angle mode | Constant |
| RF spoiling | On |
| Phase Enc. Rewinder | On |
| Cine | Off |

#### Sequence - Assistant

| | |
|---|---|
| Mode | Off |
| Allowed delay | 0 s |
